# Population-Level Associations in the Spread of Co-Circulating Respiratory Viruses: A Multi-Method Statistical Investigation Using Incidence Data

**DOI:** 10.1101/2025.11.20.25340550

**Authors:** Nadine Barth, Gesa Carstens, Eva Kozanli, Wanda Han, Lisa Hermans, Daniela Paolotti, Steven Abrams, Geert Molenberghs, Niel Hens, Christel Faes, Dirk Eggink, Albert Jan Van Hoek, Andrea Torneri

**Affiliations:** Interuniversity Institute for Biostatistics and statistical Bioinformatics, Data Science Institute, Hasselt University, Hasselt, Belgium; Dutch National Institute for Public Health and the Environment, Centre for Infectious Disease Control, Bilthoven, The Netherlands; Global Health Institute, Department of Family Medicine and Population Health, University of Antwerp, Antwerp, Belgium; Interuniversity Institute for Biostatistics and statistical Bioinformatics, Leuven Biostatistics and Statistical Bioinformatics Centre, KU Leuven, Leuven, Belgium; University of Rome La Sapienza, Rome, Italy; Centre for Health Economics Research and Modelling Infectious Diseases, Vaccine and Infectious Disease Institute, University of Antwerp, Antwerp, Belgium; ISI Foundation, Torino, Italy

## Abstract

Respiratory infections remain a major global health burden, causing substantial morbidity and mortality worldwide. The responsible viruses circulate concurrently, potentially affecting each other’s dynamics, yet the extent and direction of such interactions remain poorly understood. Characterising these cross-pathogen effects at the population level is essential for elucidating transmission dynamics and guiding mitigation strategies. Using incidence data from a participatory syndromic surveillance system with multiplex PCR confirmation of specific pathogens, we applied complementary statistical approaches, including multivariate regression, endemic–epidemic, and distributed-lag models, to characterise immediate and delayed associations among seven major respiratory diseases. We show that these pathogens form a connected system in which some, such as SARS-CoV-2 and human seasonal coronaviruses, enhance each other’s transmission, whereas others, notably influenza, inhibit the concurrent circulation of competitors such as rhinovirus or parainfluenza virus. Effects were often directional rather than reciprocal: for instance, rhinovirus inhibited human seasonal coronaviruses but not vice versa, while mutual enhancement between human metapneumovirus and parainfluenza virus appeared across several models. Interaction patterns were time-dependent yet largely consistent, indicating persistent ecological interference among co-circulating respiratory viruses. By integrating multiple analytic frameworks, our study provides a comprehensive, data-driven view of how respiratory viruses coexist and compete, offering crucial insights for improved epidemic forecasting and mitigation strategies.

## 1 Introduction

Respiratory pathogens pose a persistent and substantial public health threat, being a leading cause of mortality and morbidity worldwide [1, 2]. While often mild, infections caused by these pathogens can lead to severe outcomes, including hospitalisation, disability, or even death, particularly in vulnerable populations [3]. This was especially evident during the recent COVID-19 pandemic, which vividly demonstrated the immense societal, economic, and health-care impact that major respiratory viral outbreaks can have [4]. Beyond a pandemic context, the burden of acute respiratory infections (ARIs) is substantial even during regular seasonal epidemics. For instance, seasonal influenza alone causes an estimated 3-5 million severe cases and up to 650,000 deaths worldwide each year [5], while respiratory syncytial virus leads to around 33 million infections, 3-4 million hospitalisations, and more than 100,000 deaths among children under five annually [6, 7]. More generally, ARIs increase disability-adjusted life-years, accounting for life-years lost due to premature death and years living with disability, while quality-adjusted life-years are reduced as a consequence of ARIs, thereby demonstrating their impact on human health [8, 9].

Understanding and controlling the spread of ARIs is challenging because they represent a group of pathogens that are difficult to distinguish and disentangle, given their similar symptomatology. The complex interplay between these different pathogens further complicates their transmission dynamics at both the host and population levels. At the host level, interactions are driven by immunological phenomena that occur either during co-infection or as a consequence of prior exposures [10]. These mechanisms include cross-immunity within pathogen families (e.g. between influenza strains or among human coronaviruses) and, more broadly, heterologous immunity across unrelated pathogens, which can lead to both protective and harmful effects [11]. Host-level variability further reflects differences in the degree of infectiousness among infected individuals, susceptibility to infection, and immune responses triggered by prior exposures. A notable harmful effect is immune misjudgement of a novel virus, potentially triggering dysregulated responses such as cytokine storms, as documented for the 1918 influenza pandemic [12]. Another critical example is antigenic imprinting, where a host’s initial immunological memory shapes subsequent, potentially suboptimal, responses to new variants. Such mechanisms can influence transmission dynamics by altering host susceptibility to subsequent infections and thereby modifying population-level patterns of spread. Beyond these host-level effects, pathogen interactions at the population level can manifest as changes in epidemic curves, such as viral competition causing pathogens to peak at different times or viral cooperation leading to simultaneous peaks. Population-level dynamics are further shaped by heterogeneity in social behaviour, such as self-isolation during illness, which reduces contact rates and in turn affects the pool of susceptible individuals that can be reached by co-circulating pathogens [13].

Theoretical and simulation-based mathematical modelling studies have shown that cross-pathogen interactions can profoundly alter epidemic dynamics [14, 15]. For example, these models predict that cross-immunity between pathogens can lead to a reduced attack rate and an increased probability of extinction for one or both viruses, effectively suppressing their spread [16, 17]. Conversely, enhancing interactions, such as increased severity or susceptibility to secondary infections, can lead to larger or prolonged outbreaks and greater healthcare burden [18]. As a consequence, a thorough understanding of these complex inter-pathogen dynamics is potentially critical for designing effective and informed public health interventions. When such associations exist, they have the potential to reshape infection risks at the individual level and the timing and magnitude of epidemics at the population level, underscoring the need to establish their presence and character.

Given the profound implications of potential interactions, several studies have empirically investigated these relationships using both mathematical and phenomenological approaches, though often limited by available data For instance, Chiavenna et al. [19] used a time series model to estimate age-stratified influenza-associated invasive pneumococcal disease, investigating enhancing relationships between a virus and a bacterium. Similarly, Shrestha et al. [20] employed mathematical models to characterise the enhanced risk of pneumococcal pneumonia following influenza infection. More recently, Madewell et al. [21] applied multivariate Bayesian hierarchical modelling to identify population-level virus–virus interactions in Puerto Rico, based on correlations between residual odds of infection. While these studies provide valuable insights into specific pathogen pairs or co-occurrence patterns in particular climates, they highlight the difficulty of analysing interactions across many respiratory pathogens, largely due to limited pathogen-specific data [22]. Crucially, a major gap remains in understanding inter-pathogen influences across shortand long-term lags and in characterising spatial co-occurrence patterns.

To investigate the associations among co-circulating respiratory pathogens, we employed several statistical frameworks to quantify diverse forms of cross-pathogen interactions and their temporal dynamics. Specifically, we used latent Gaussian models (LGMs) to identify co-occurrence patterns, generalised regression frameworks to examine short-term lagged effects, the endemic-epidemic (hereafter referred to as E&E) model to separate endemic and epidemic contributions, and distributed lag nonlinear models (DLNMs) to characterise long-term lagged interactions. We relied on surveillance data collected by the National Institute for Public Health and the Environment (RIVM), the Netherlands, through *infectieradar*, a participatory online surveillance platform [23]. These data are unique as they consist of high-resolution, laboratory-confirmed weekly incidence of common ARIs for the period between 2022 and 2025. By applying the aforementioned statistical approaches, we assessed the directionality and persistence of associations among co-circulating viral respiratory pathogens and evaluated how spatial and temporal scales influence them.

## 2 Methodology

We define *cross-pathogen interactions* as any direct or indirect influence the spread of one pathogen has on the transmission dynamics or clinical outcome of another co-circulating pathogen within a host population. These interactions may be considered *cooperative* or *enhancing* if the presence or activity of one pathogen leads to an increased incidence, transmission, or severity of illness of another. Conversely, they are considered *competitive* or *competing* if they lead to a decreased incidence, transmission, or severity of illness of another. While this provides general definitions, in this manuscript we focus on the epidemiological co-occurrence of pathogens and temporal associations in disease incidence that may reflect interactions between pathogens.

We conducted our analysis considering different levels of complexity. First, we performed exploratory analyses, including Granger causality [24] and Principal Component Analysis (PCA) [25], to gain initial insights into predictive relationships and spatial components. Next, we used LGMs [26] to assess co-occurrence patterns and afterwards three related generalised regression frameworks to investigate short-term lagged effects: generalised linear mixed models (GLMMs) [27], vector generalised linear models (VGLMs) [28], and generalised additive models for location, scale, and shape (GAMLSS) [29]. To ensure comparability across approaches, we maintained model assumptions and outcome distributions as similar as possible, aligning structural components such as covariate effects and temporal resolution. The E&E model [30, 31] was also employed for short-term effects. Last, the distributed lag nonlinear models (DLNMs) [32] were used for investigating long-term interactions. We considered seven respiratory pathogens for our main analyses: severe acute respiratory syndrome coronavirus 2 (SARS-CoV-2), respiratory syncytial virus (RSV), influenza viruses (IV), rhino-/enterovirus (RV/EV) complex, human metapneumovirus (hMPV), parainfluenza virus (PIV), and human seasonal coronaviruses (hCoV). These represent the most prevalent viral respiratory agents. Subtypes were aggregated for IV, PIV and hCoV primarily to increase case counts and ensure sufficient statistical power, while maintaining epidemiological coherence within each virus group. Bacterial and less common pathogens were excluded due to their low detection frequency and differing transmission routes. Disease counts were aggregated weekly (Monday—Sunday) at either the national level or at the NUTS-2 level, following the Nomenclature of Territorial Units for Statistics (NUTS) of Eurostat [33], corresponding to the 12 provinces of the Netherlands.

### 2.1 Disease Surveillance

This study relied on data collected by *Infectieradar* (infection radar), a participatory online surveillance platform operated by the Dutch National Institute for Public Health and the Environment (RIVM) [34]. The platform used a longitudinal, opt-in cohort design where participants voluntarily reported symptoms weekly. In addition to symptom reports, the platform collected information on testing behaviour, basic health characteristics, healthcare seeking behaviour, and social contact patterns, and it included a laboratory testing component for a subset of participants [35, 23]. Our analysis used data spanning three distinct epidemiological seasons. The specific data for our analysis were collected between 4 October 2022 and 12 May 2025. The analyses covered the entire study period, including inter-seasonal months, to capture continuous circulation patterns. Data were analysed across all seasons to capture overall associations across multiple epidemic periods and to ensure sufficient statistical power for less prevalent pathogens. For the sake of clarity, we define an epidemiological season as the period from 1 September of one year to 30 April of the subsequent year. Unless stated otherwise, all mentions of ‘season’ refer to this interval. The dataset included respiratory disease cases confirmed via multiplex PCR assays, collecting a total of 7,642 confirmed infections for the selected diseases. We excluded cases with missing regional information (*n* = 167) and those from participants under 20 years (*n* = 43) due to insufficient representation. Participants were subsequently aggregated into weekly counts per pathogen, stratified by geographical region (NUTS2) and age group (for two age groups: 20-65 years and > 65 years).

### 2.2 Exploratory Analysis

#### Graphical Exploration

First, a graphical inspection of the time series of weekly incidence for each pathogen, both overall and by region was performed, to visually identify broad patterns of co-circulation and spatial variations. This descriptive step served to generate hypotheses and guided the subsequent modelling, but did not provide statistical inference or control for potential confounding factors.

#### Investigating Directional Predictive Relationships

We examined whether past incidence of one pathogen statistically predicted the future incidence of another using Granger causality within a multivariate vector autoregressive (VAR) framework. Stationarity was enforced through variance-stabilising transformations and differencing, and lag order was selected by information criteria. The multivariate formulation accounted for simultaneous co-circulation of pathogens, reducing artefactual associations from purely pairwise analyses. This approach identifies temporal predictability rather than mechanistic causation. Full mathematical details, preprocessing steps, assumption checks, and regional results are provided in Appendix B.

#### Investigating Spatial Clustering and Disease Associations

We applied PCA to explore dominant patterns of temporal co-variation among respiratory pathogens. Weekly case counts were aggregated over age groups, and each pathogen was standardised within region to remove differences in absolute incidence and highlight relative temporal dynamics. First, we performed a national analysis with counts aggregated across regions. Next, we applied PCA separately by region to assess spatial heterogeneity. Components were retained based on the Kaiser criterion (eigenvalue *>* 1) and visual inspection of scree plots. Loadings were interpreted to evaluate each pathogen’s contribution to the principal components (PCs), and cosine similarity between loading vectors was computed to quantify temporal association patterns across pathogen pairs. To formally evaluate global spatial clustering, we assess systematic differences in multivariate temporal patterns across regions by applying permutational multivariate analysis of variance (PERMANOVA) and multivariate analysis of variance (MANOVA).

PCA provides a reduced-dimension representation of shared temporal variance, but it does not identify causal or directional interactions between pathogens. Because standardisation removes differences in absolute incidence, the analysis reflected relative co-variation only, and the variance explained by the retained components accounted for part, but not all, of the epidemic complexity. Full methodological details and extended results can be found in Appendix C.

### 2.3 Residual Inter-Pathogen Correlations

To quantify contemporaneous dependencies among the selected respiratory pathogens, we developed a LGM with correlated residuals. This hierarchical Bayesian formulation jointly models weekly incidence across pathogens, allowing estimation of instantaneous cross-pathogen correlations after adjustment for demographic, regional, and temporal effects. Weekly counts *Y*_*d,r,a,t*_ for pathogen *d*, region *r*, age group *a*, and week *t* followed a negative binomial distribution

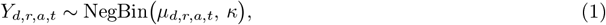

where *µ*_*d,r,a,t*_ is the expected mean and *κ* a shared over-dispersion parameter. The linear predictor combined pathogen-, age-, region-, and time-specific components:

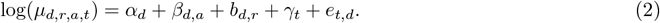

Here *α*_*d*_ is a disease-specific intercept, *β*_*d,a*_ an age effect structured by the contact matrix *C, b*_*d,r*_ a region-bydisease random effect, *γ*_*t*_ a shared temporal component represented by a fixed spline basis of dimension 5, and *e*_*t,d*_ a residual term capturing cross-pathogen correlation at time *t*. Social contact data were obtained from the POLYMOD study of Mossong et al. [36], accessed via the publicly available SOCRATES data tool [37].

At each week, residuals formed a *K*-dimensional Gaussian vector **e**_*t*_ ~ 𝒩_*K*_(**0**, Σ_*e*_), with 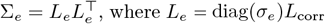, where *L*_*e*_ = diag(*σ*_*e*_)*L*_corr_. We used a Lewandowski-Kurowicka-Joe (LKJ) prior to have a weakly informative correlation structure among pathogens *L*_corr_, and exponential penalised-complexity priors on all standard deviations to ensure shrinkage toward zero.

Posterior correlations derived from Σ_*e*_ quantify the remaining synchronous variation among pathogens. Positive values indicate cooperative contemporaneous behaviour, whereas negative values imply residual competition. The full model specification and estimation details are given in Supplementary Section C.5.

### 2.4 Short-Term Lagged Effects

We investigated one-week lagged effects of each pathogen on all others using regression frameworks that included directly lagged counts as predictors. To test the robustness of short-term associations, we compared several frameworks differing in how they represent spatial and distributional heterogeneity.

Weekly counts *Y*_*d,r,a,t*_ were assumed to follow a negative binomial distribution, *Y*_*d,r,a,t*_ ~ NegBin(*µ*_*d,r,a,t*_, *κ*), where *µ*_*d,r,a,t*_ = *E*(*Y*_*d,r,a,t*_) denotes the mean count and is modeled using a log link function in relation to the predictors. These predictors included one-week lags of disease incidence for all pathogens, age group and region effects, a seasonal effect and a smooth temporal trend. The seasonal term is defined as

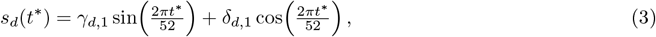

and temporal smooths used a natural cubic spline *f*_*d*_(*t*\*) with five degrees of freedom. Details on model selection are provided in Appendix D. Model coefficients were exponentiated and expressed as incidence rate ratios (IRRs), representing the multiplicative change in mean weekly counts per one-unit increase in a specific predictor value, with other covariates being fixed.

#### Generalised Linear Mixed Model (GLMM)

The GLMM incorporated a random intercept for region to quantify cross-pathogen associations while accounting for unobserved spatial heterogeneity, thereby avoiding a large number of nuisance parameters since inference was not targeted at region-specific effects and assuming a common dispersion across regions. The random intercept *b*_*r*_ was assumed normally distributed, 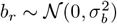, and the mean count *µ*_*d,r,a,t*_ followed

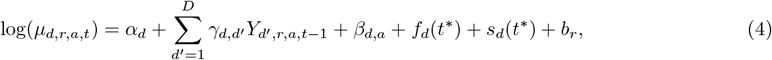

where 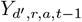 denotes the one-week lagged count of pathogen *d*′ in region *r* and age group 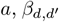 quantifies the cross-pathogen lag effect, 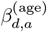 the age effect, *s*_*d*_(*t*\*) and *f*_*d*_(*t*\*) the seasonal and smooth temporal components, and *b*_*r*_ the regional random effect. Dispersion was controlled by a single parameter *κ* common to all pathogens, regions, age groups and time points.

#### Vector Generalised Linear Model (VGLM)

The VGLM excluded random effects and instead modelled region as fixed terms, while introducing pathogen-specific dispersion parameters (*κ*_*d*_) to capture differences in overdispersion between viruses. This structure allowed pathogen-specific variability and ensured comparability of lag effects across pathogens, though residual regional heterogeneity could not be explicitly accounted for. The mean count *µ*_*d,r,a,t*_ was modelled as

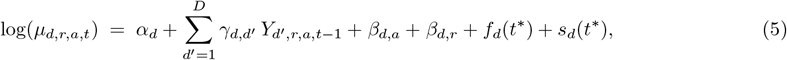

retaining the same lag, age, and temporal components as specified before.

#### Generalised Additive Models for Location, Scale and Shape (GAMLSS)

The GAMLSS extends the previous formulations by jointly modelling the mean and dispersion components of the negative binomial response, providing additional flexibility to accommodate pathogen-specific differences in overdispersion. It retained the same covariate structure and fixed region effects as the VGLM, thereby preserving comparability of mean effects but without accounting for residual regional heterogeneity. The mean sub-model was

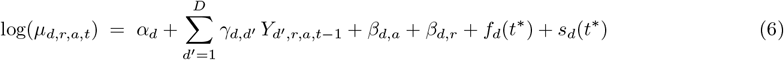

and the dispersion parameter followed log(*κ*_*d*_) = *δ*_*d*_, constant within each pathogen.

This formulation thus allows pathogen-specific variance structures while maintaining a common mean specification. More complex timeor covariate-dependent dispersion models were evaluated but not supported by model selection.

### 2.5 Disentangling Baseline and Transmission Effects

In addition to the one-week lagged regression models, we applied the E&E framework, which decomposes incidence into background and transmission-driven components. Although lagged predictors were defined on the same short-term scale, this approach distinguishes how cross-pathogen effects are distributed between longterm shared drivers, such as seasonality or susceptibility, and short-term dependencies linked to concurrent transmission. Model covariates, temporal smooths, and seasonal terms followed the same specifications as in the short-term frameworks for comparability. The model assumed a fixed spatial and contact structure across the study period and did not include random effects or pathogen-specific dispersion. The mean structure is

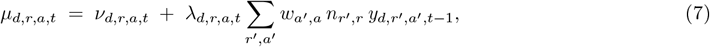

where *y*_*d,r,a,t*_ is a realisation of the weekly count for pathogen *d* in age group *a* and region *r* at time point 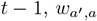 age-specific contact weights, and 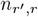 regional neighbourhood weights (restricted to within-region transmission). Non-negativity of the mean *µ*_*d,r,a,t*_ is ensured by the log-link formulations of *ν*_*d,r,a,t*_ and *λ*_*d,r,a,t*_ and by the non-negative contact and neighbourhood weights.

The epidemic component was parameterised as

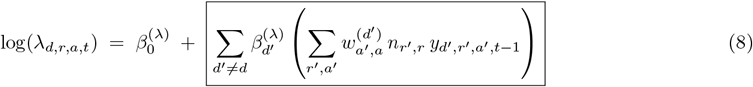

and the endemic component as

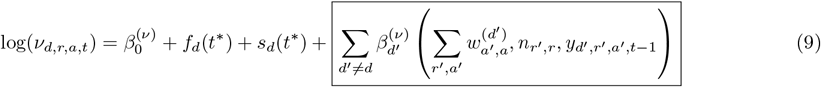

To investigate how lagged pathogen counts affect background and transmission dynamics, we explored three configurations: including lagged counts only in the endemic component, only in the epidemic component, or in both. The boxed terms in (8) and (9) represent these cross-pathogen contributions (*d*′ = *d*) through their lagged, contact- and region-weighted counts. Each component also contains a pathogen-specific intercept (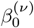 or 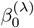), capturing baseline intensity when cross-pathogen terms are absent. The epidemic component already includes an autoregressive term for the same pathogen (*d*), so additional lagged predictors were restricted to other pathogens (*d*′ ≠ *d*) to avoid parameter redundancy and to ensure identifiability.

### 2.6 Long-Term Lagged Effects

Finally, we extended the short-term lagged models to investigate delayed cross-pathogen effects over multiple weeks by embedding a DLNM within the VGLM framework. This model captures potentially nonlinear associations while assuming that lag structures are constant across regions and age groups and does not incorporate random effects or shared latent components. As before, the outcomes were weekly counts *Y*_*d,r,a,t*_ of pathogen *d* in region *r*, age group *a*, and week *t*. Mean counts *µ*_*d,r,a,t*_ were modelled on the log scale with categorical effects for age group and region, a smooth temporal trend with annual seasonality, and lagged pathogen counts were represented via cross-basis functions.

The mean is specified as

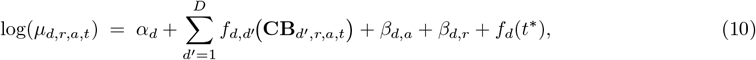

where *f*_*d,d*_*′* (*·*) denotes the effect of past incidence of pathogen *d*′ on pathogen *d*, represented through a cross-basis transformation **CB**_*d*_*′*_,*r,a,t*_. Each cross-basis maps the history of pathogen *d*′ in region *r*, age group *a*, and week *t* onto a two-dimensional basis over exposure (past counts) and lag (weeks since exposure):

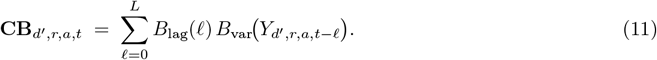

Here, *B*_var_(*·*) is a linear basis for the exposure dimension, and *B*_lag_(*ℓ*) represents B-splines of degree one across lags *ℓ* = 0, …, *L*.

We considered a maximum lag of *L* = 8 weeks, with quadratic and cubic B-spline variants evaluated in sensitivity analyses (see Appendix G). This construction allows estimation of the full lag-response surface, i.e., relative risks (RRs) as a function of both exposure level and lag. The function *f*_*d,d*_*′* (*x, ℓ*) denotes the smooth bivariate lag–response surface estimated from the cross-basis, describing how past incidence of pathogen *d*′ at exposure level *x* and lag *l* affects pathogen *d*. The corresponding relative risk is *RR*(*x, ℓ*) = exp *f*_*d,d*_*′* (*x, ℓ*). Full basis specification and parameterisation are provided in Appendix G.1.

### 2.7 Subtype-Level Interaction Analysis

Furthermore, we investigated interactions within specific viral families for which data were available. More precisely, we looked at short- and long-term effects for influenza subtypes (A-H1N1, A-H3, and B) and coronaviruses (SARS-CoV-2 and its co-circulating human seasonal coronaviruses subtypes: 229E, OC43, and NL63/HKU1, already combined as one target in the multiplex PCR). To do so, we used the DLNM model described above.

### 2.8 Software and Implementation

All statistical analyses were performed using `R`(version 4.4.1) [38]. Specific `R`packages used for modelling and inference include `glmmTMB`, `VGAM`, `gamlss`, `MASS`, `dlnm`, `surveillance`, `hhh4contacts`, `nimble`, and `vars`, with supporting packages for diagnostics and dimension reduction such as `lmtest`, `car`, `DHARMa`, `FactoMineR`, `factoextra`, `vegan`, `tseries`, `spdep`, `ISOweek`, and `splines`. Model comparison and selection for frequentist models relied on Akaike Information Criterion (AIC) and Bayesian Information Criterion (BIC). Comprehensive residual diagnostics were performed to assess model adequacy, with the `DHARMa`package specifically used for scaled residual diagnostics in GLMMs. For frequentist models, inference was based on point estimates accompanied by asymptotic 95% confidence intervals, supplemented by two-sided *p*-values at a 5% significance level. For Bayesian models, posterior means were generally reported as point estimates and uncertainty expressed through 95% CrIs, with effects considered notable when CrIs did not include zero. No adjustments for multiplicity were applied, and the resulting analyses should therefore be regarded as exploratory rather than confirmatory assessments of cross-pathogen associations.

## 3 Results

### 3.1 Exploratory Data Analysis

This exploratory analysis summarises temporal and spatial patterns in respiratory pathogen circulation to contextualise subsequent modelling of inter-pathogen associations. Weekly confirmed cases aggregated across all pathogens showed a strong seasonal pattern (Figure A.1). Seasonal interpretation follows the epidemiological definition given above. In addition, the time series demonstrated clear spatial heterogeneity in case counts (Figure A.2), while peak timing remained broadly consistent across regions for most pathogens (Figure A.3). Regional curves for each pathogen are provided in Appendix A.

Pathogen-specific smoothed series revealed distinct dynamics for each pathogen (Figure 1). IV, RSV, and hMPV exhibited pronounced winter waves, with RSV and hMPV dropping to near-zero in between epidemiological seasons. SARS-CoV-2 and hCoV remained detectable year-round but peaked in winter. PIV and RV/EV were detectable throughout the year and often peaked from late summer to autumn.

**Figure 1:**
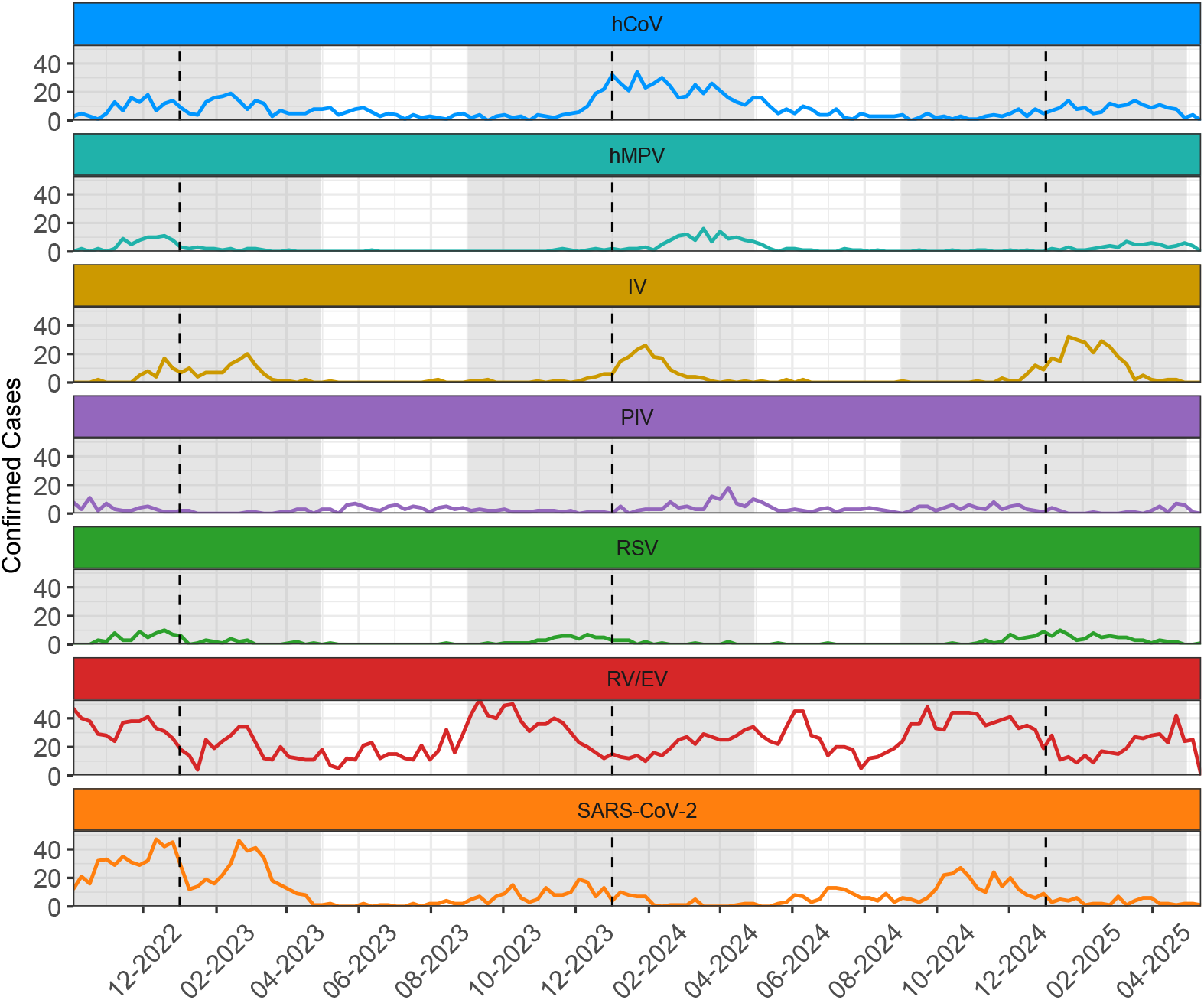
Weekly confirmed cases by respiratory pathogen over time, aggregated across all regions, from 4 October 2022 to 12 May 2025. Vertical dashed lines indicate the start of each calendar year. Shaded areas indicate epidemiological seasons.

Case counts were sparse at the week–region level, consistent with concentrated seasonal peaks; zero-week proportions ranged roughly from ~ 50% for RV/EV to ~ 93% for RSV (Table A.1). Over the full period, RV/EV contributed the largest share of confirmed cases (45% of all cases), followed by SARS-CoV-2 (18%), hCoV (12%), IV (12%), PIV (5%), hMPV (4%) and RSV (3%).

#### Investigating Directional Predictive Relationships

Building on these descriptive patterns, this analysis quantifies potential directional relationships among respiratory pathogens, assessing whether fluctuations in one pathogen’s incidence systematically precede changes in another. The national-level analysis (Table B.2 in Appendix B) revealed that most pathogen pairs did not exhibit significant temporally ordered relationships. However, three pairs stood out: hMPV predicted IV (*p* = 0.0074), and vice versa (*p* = 0.0114). In addition, SARS-CoV-2 showed a borderline significant predictive effect on PIV (*p* = 0.0429).

To further investigate spatial variations, separate Granger causality tests were performed for each region. Not all pathogen pairs could be assessed in every region because some regional time series were too sparse to fit a valid VAR model. Compared to the national-level analysis, which identified only three significant associations, the regional analyses revealed a more heterogeneous picture (Figure 2). Several pathogen pairs showed significant associations in multiple regions, although no pair was consistently significant across all regions. For instance, hMPV predicting PIV was significant in 38% (3*/*8) of regions, RSV predicting IV in 50% (5*/*10), IV predicting RSV in 38% (3*/*8), and SARS-CoV-2 predicting RSV in 38% (3*/*8) (Table B.3). These findings highlight that predictive links between pathogens are highly dependent on the region.

**Figure 2:**
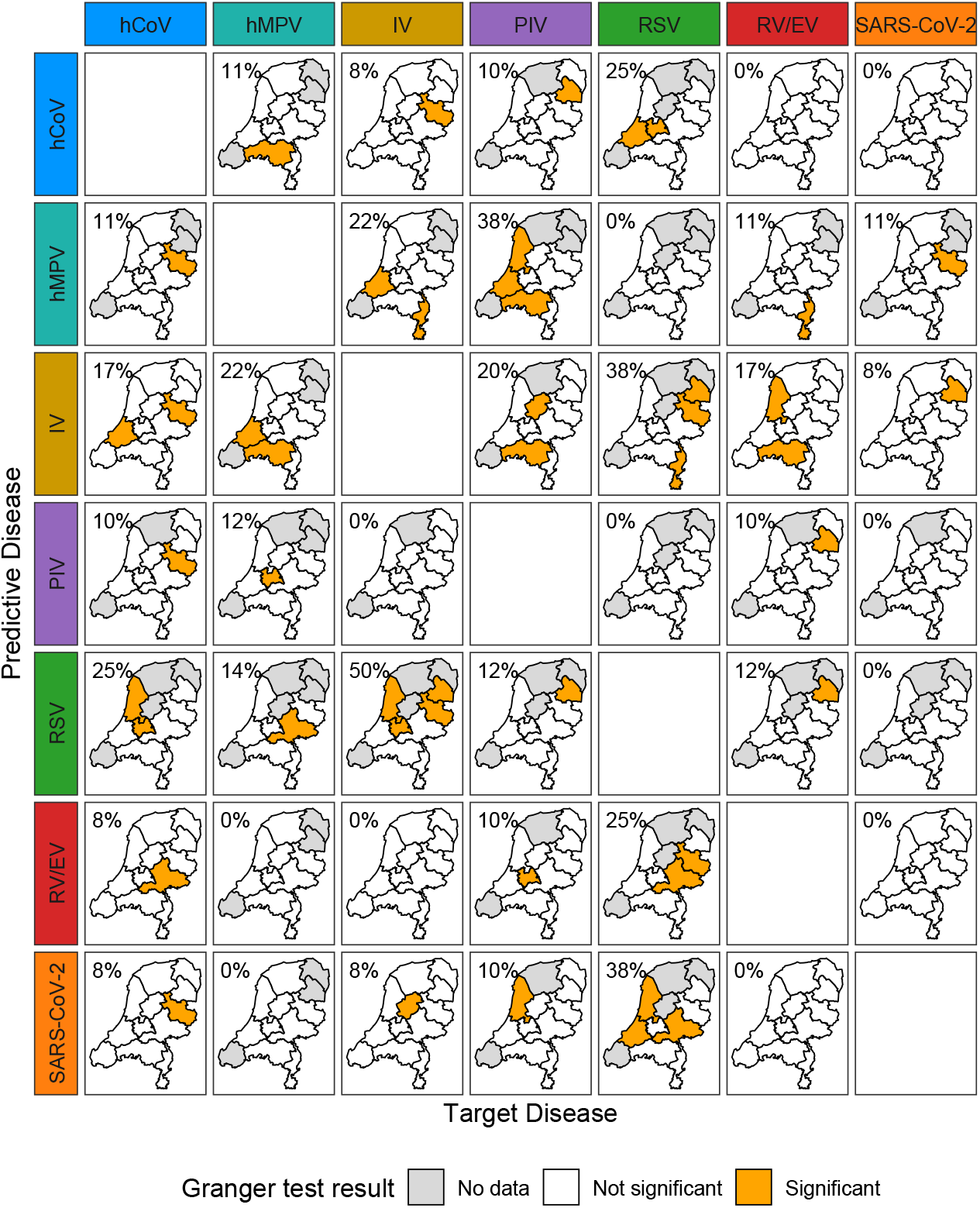
Regional Granger causality analysis. Each panel shows the Netherlands map for a given predictive (row) and target (column) pathogen pair. Provinces are colored according to Granger test results: orange for statistically significant associations (*p <* 0.05), white for non-significant results, and gray where no or too little data were available. The proportion of regions in which the result was significant is indicated in the top left corner of each panel.

To evaluate the robustness of the VAR findings, we performed a sensitivity analysis based on purely pairwise Granger causality tests. While the multivariate VAR accounts for the influence of all other pathogens simultaneously, the pairwise approach examines two series in isolation. The comparison of regional significance proportions indicates broadly consistent patterns across both specifications. At the national level, several direct associations reached statistical significance in the pairwise tests (Table B.4). Examples include hMPV→IV, IV→hMPV, and SARS-CoV-2→PIV, which were detected only in the pairwise setting but not in the multivariate framework. Together, these results underline the value of modelling all pathogens together, rather than in a purely pairwise manner.

#### Investigating Spatial Clustering and Disease Associations

Extending beyond pairwise temporal relationships, this analysis examines the principal modes of shared temporal variation among respiratory pathogens and evaluates how consistently these patterns manifest across regions. The pooled PCA showed that the first three PCs explained 19.5%, 16.7%, and 15.7% of the total variance, respectively (cumulative 51.9%). Scatter plots of PC scores showed no systematic regional clustering (Figure C.2), and both PERMANOVA and MANOVA tests (*p* = 1.0000) indicated that, although temporal patterns varied between regions, these differences were irregular rather than forming coherent spatial groups.

The PCA identifies dominant modes of temporal variation across pathogens. Pathogens with loading vectors pointing in the same direction on a given PC show similar timing and shape in their temporal dynamics, whereas those pointing in opposite or orthogonal directions represent asynchronous or distinct seasonal patterns (Figure C.4). The arrows indicated both the direction and magnitude of each pathogen’s contribution, so that angular closeness reflected synchrony and opposition reflected anti-phase relationships. This visualisation showed, for example, that IV and hCoV pointed broadly in the same direction on PC1, whereas PIV and RV/EV were rotated towards PC2, capturing a partially distinct seasonal component. Cosine similarity (Table 1) between loading vectors formalised these patterns. Strong alignment was found between hCoV and hMPV (0.921) and between RSV and SARS-CoV (0.761), while clear opposition emerged between IV and PIV (−0.625) and RSV and PIV (−0.655).

**Table 1:**
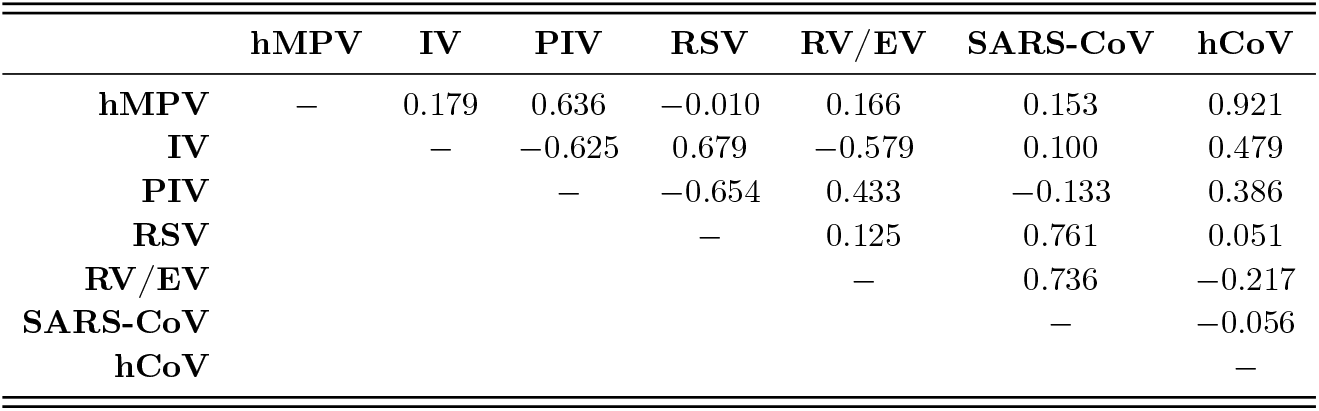
Cosine similarity between pathogen loading vectors. Values represent the cosine similarity between loading vectors of each pathogen pair across the first three PCs.

|  | **hMPV** | **IV** | **PIV** | **RSV** | **RV/EV** | **SARS-CoV** | **hCoV** |
| --- | --- | --- | --- | --- | --- | --- | --- |
| **hMPV** | — | 0.179 | 0.636 | $-0.010$ | 0.166 | 0.153 | 0.921 |
| **IV** | | — | $-0.625$ | 0.679 | $-0.579$ | 0.100 | 0.479 |
| **PIV** | | | — | $-0.654$ | 0.433 | $-0.133$ | 0.386 |
| **RSV** |  |  |  | — | 0.125 | 0.761 | 0.051 |
| **RV/EV** | | | | | — | 0.736 | $-0.217$ |
| **SARS-CoV** | | | | | | — | $-0.056$ |
| **hCoV** |  |  |  |  |  |  | — |

At the regional scale, PCA explained a similar proportion of variance (~ 56%, Figure C.6) but showed substantial heterogeneity in loadings across regions (Figure C.7). Mean cosine similarities are shown with 95% confidence intervals quantifying between-region variability in Figure C.8. They confirmed stable cooperative associations such as between hCoV and hMPV (mean: 0.44, 95% CI: [0.13; 0.75]) and RSV and SARS-CoV-2 (mean: 0.56, 95% CI: [0.28; 0.84]), and identified a consistent competing association of IV and PIV (mean: −0.40, 95% CI: [−0.64; −0.17]). However, wide confidence intervals for several pathogen pairs reflected strong differences in cosine similarity estimates across regions.

### 3.2 Residual Inter-Pathogen Correlations

Following the exploratory analyses, this section introduces a modelling approach incorporating demographic, spatial, and temporal covariates to estimate residual contemporaneous correlations between pathogens, thereby assessing whether any co-fluctuation remains unexplained by shared factors. Posterior estimates from the LGM revealed clear residual dependencies among several pathogens after adjustment for demographic, regional, and temporal effects (Figure 3). Significant positive residual correlations were observed between for example IV and RSV (*ρ* : 0.51, 95% CrI: [0.36; 0.64]) and between SARS-CoV-2 and RSV (*ρ* : 0.45, 95% CrI: [0.29; 0.59]), indicating coordinated increases in their circulation. Several pairs exhibited significant negative residual correlations, for example IV and PIV (*ρ* : −0.46, 95% CrI: [−0.62; −0.27]). Overall, this framework identified both cooperative and competitive residual relationships. While these correlations identify synchronous or opposing fluctuations, the model does not estimate the directionality of these associations. Overall, this framework identified both cooperative and competitive residual relationships among the analysed pathogens.

**Figure 3:**
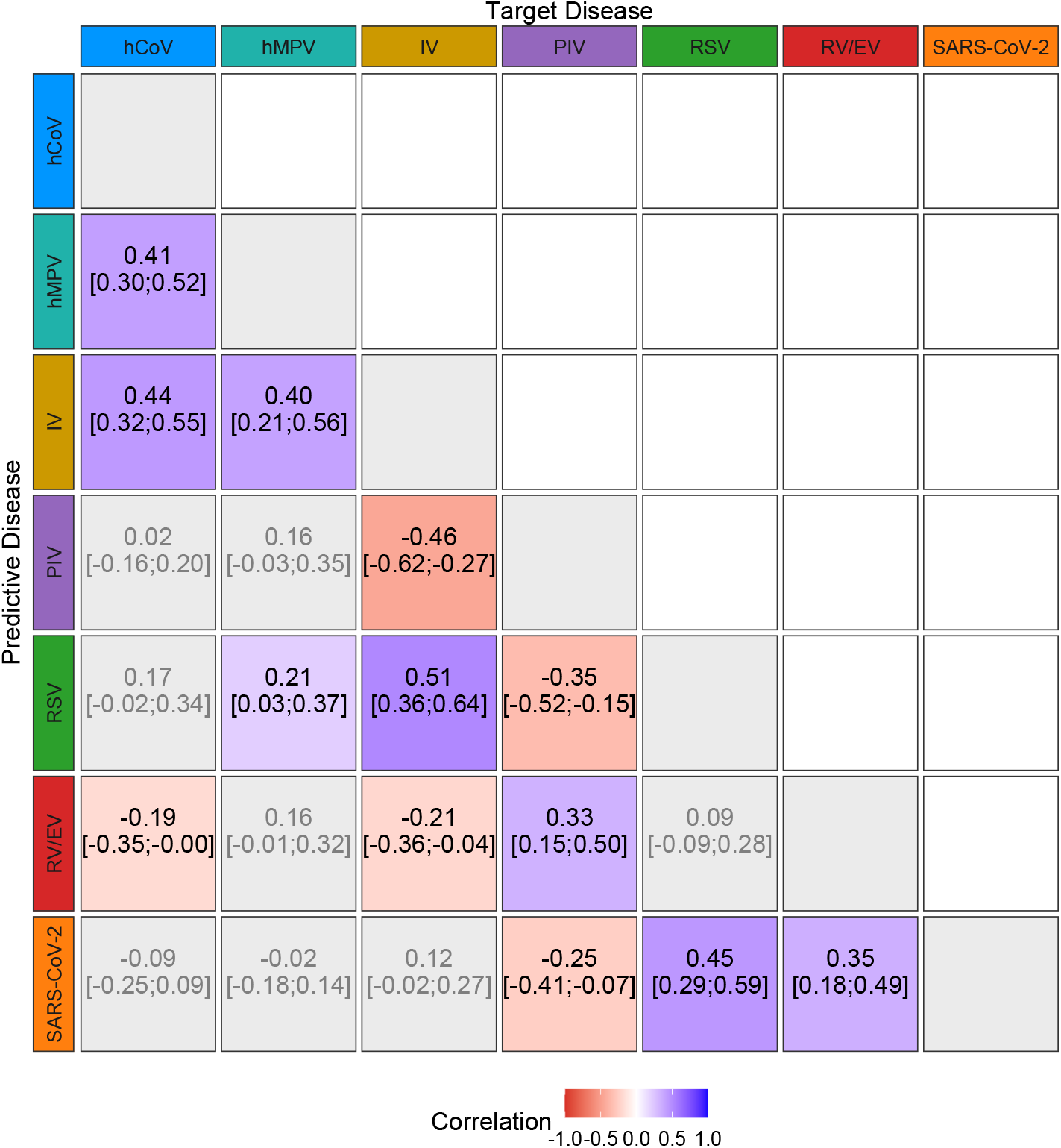
Posterior mean residual correlations between respiratory pathogens estimated from the LGM. Values indicate mean posterior correlations with corresponding 95% credible intervals. Color intensity reflects the magnitude and sign of correlation (blue for positive, red for negative). Correlations represent contemporaneous associations and do not imply directionality.

**Figure 4:**
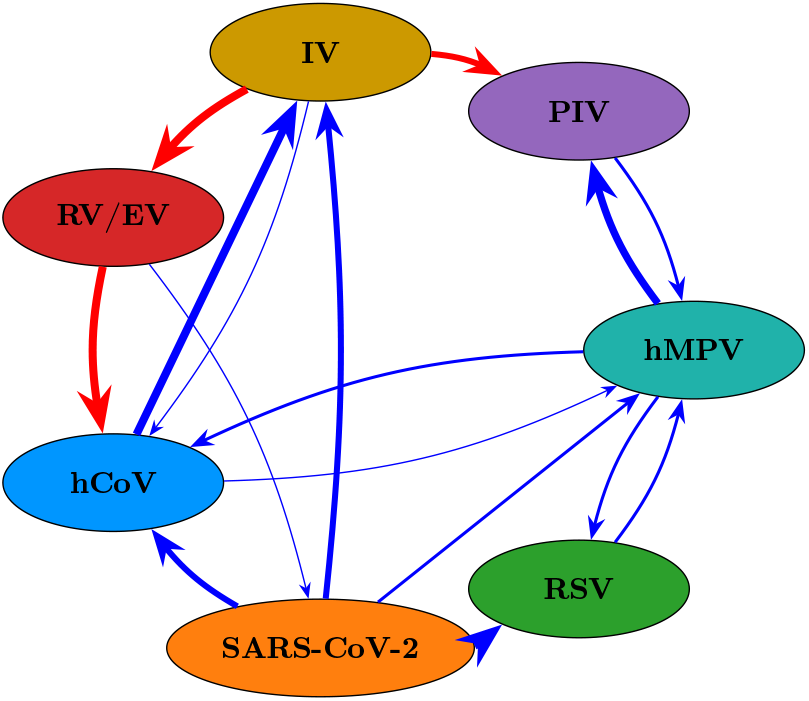
Disease interaction network based on one-week lagged regression models (GLMM, VGLM and GAMLSS). Arrows represent significant temporal associations between respiratory viruses. Blue arrows indicate enhancing (cooperative) and red arrows competing (inhibitory) effects. Arrow thickness reflects the number of models in which the association was statistically significant.

### 3.3 Short-Term Lagged Effects

Extending beyond contemporaneous correlations, this analysis quantifies short-term lagged associations between pathogens, assessing whether recent increases in one pathogen’s incidence predict changes in another after adjusting for demographic and temporal covariates. Across the three frameworks we identified a small, reproducible set of one-week cross-pathogen associations. Robust cooperative interactions across all modelling approaches emerged between several pathogen pairs: hMPV showed a consistent enhancing effect on PIV (GLMM mean IRR: 1.3259, 95% CI: [1.1272; 1.5597], VGLM mean IRR: 1.6629, 95% CI: [1.3634; 2.0282], GAMLSS mean IRR: 1.2998, 95% CI: [1.1001; 1.5358]). Similarly, SARS-CoV-2 was positively associated with RSV, while hCoV also showed a consistent enhancing effect on IV. Competitive effects included IV reducing subsequent RV/EV (GLMM mean IRR: 0.8831, 95% CI: [0.8333; 0.9358], VGLM mean IRR: 0.8790, 95% CI: [0.8204; 0.9418], GAMLSS mean IRR: 0.8784, 95% CI: [0.8320; 0.9274]), while RV/EV was negatively associated with hCoV. Further associations detected in two or only a single framework are listed in Table D.4.

Although the magnitude of these effects varied slightly between models, the direction of point estimates for cross-pathogen interactions remained remarkably consistent, indicating robust findings across different statistical frameworks. As expected, autoregressive terms were uniformly positive across pathogens, indicating temporal persistence.

### 3.4 Disentangling Baseline and Transmission Effects

When decomposing incidence into endemic and epidemic components, we can separate long-term shared drivers of circulation from short-term cross-pathogen transmission effects. We focus on a combined specification in which lagged counts enter in both components, which identifies dependencies that persist after accounting for shared background drivers (typically reflected in the endemic component) while still capturing short-term, transmission-related interactions represented by the epidemic term.

Across pathogens, the endemic component consistently accounted for the majority of fitted incidence (Table F.3). RSV, hMPV, and PIV were almost entirely endemic-driven (*>* 90%), while IV, SARS-CoV-2, and hCoV showed epidemic contributions of 15–20% on average. RV/EV displayed the largest epidemic share, with the autoregressive component explaining nearly 40% of fitted incidence. This indicated that while baseline temporal trends and exogenous factors dominated the dynamics for most pathogens, persistence played a larger role for high-burden and highly transmissible agents such as RV/EV.

The E&E models identified several significant enhancing effects between pathogens when lagged counts were included in both the endemic and epidemic components. Two associations were consistently detected across all model specifications: an enhancing effect of SARS-CoV-2 on RV/EV (mean effects estimated in epidemic component: 1.0819, 95% CI: [1.0201; 1.1474] and in endemic component: 1.0953, 95% CI: [1.0090; 1.1890]) and of hCoV on IV (mean effects estimated in epidemic component: 1.1360, 95% CI: [1.0535; 1.5868] and in endemic component: 1.5251, 95% CI: [1.3231; 1.7579]). Additional effects, such as RV/EV on IV SARS-CoV-2, PIV or hCoV on hMPV, also emerged when the same lagged covariates were entered in both components (Figure 5).

**Figure 5:**
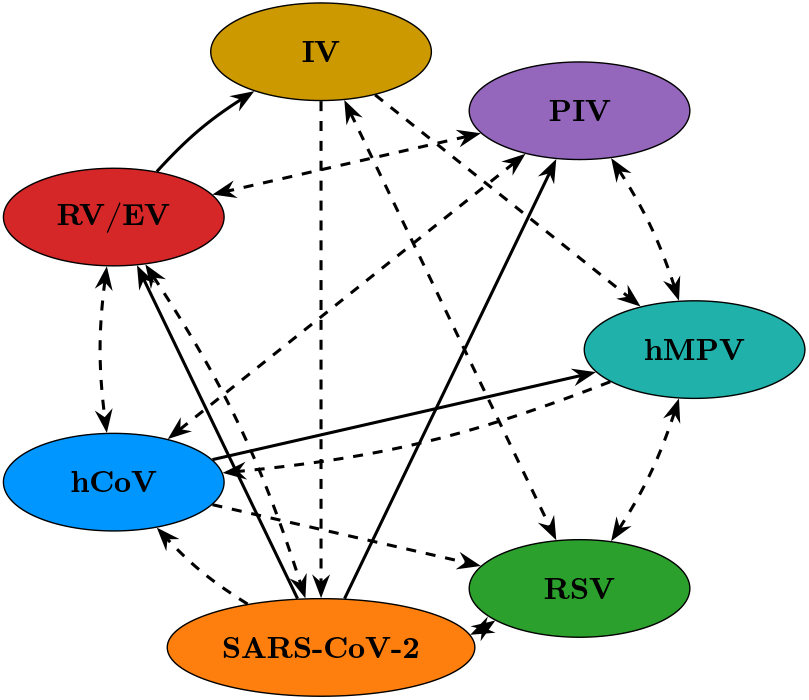
Disease interaction network based on the E&E model. Arrows represent significant temporal associations between respiratory viruses. Solid arrows indicate an effect in the epidemic component and dashed arrows in the endemic component.

When compared to the regression-based models, the E&E framework reproduced the positive association between hCoV on IV (in both components) was consistent across GLMM, VGLM, and GAMLSS. Effects of SARS-CoV-2 on hCoV ans RSV as well as hMPV on PIV could be found in the endemic component only. An enhancing effect of SARS-CoV-2 on IV was not found in the E&E model.

### 3.5 Long-term Lagged Effects

This analysis quantifies delayed and nonlinear cross-pathogen associations, capturing how one pathogen’s influence evolves over multiple weeks rather than a single lag. We used the DLNM to map cross-pathogen effects over exposure and lag. Figure 6 shows the RR over lags for a fixed predictor value of 1.

**Figure 6:**
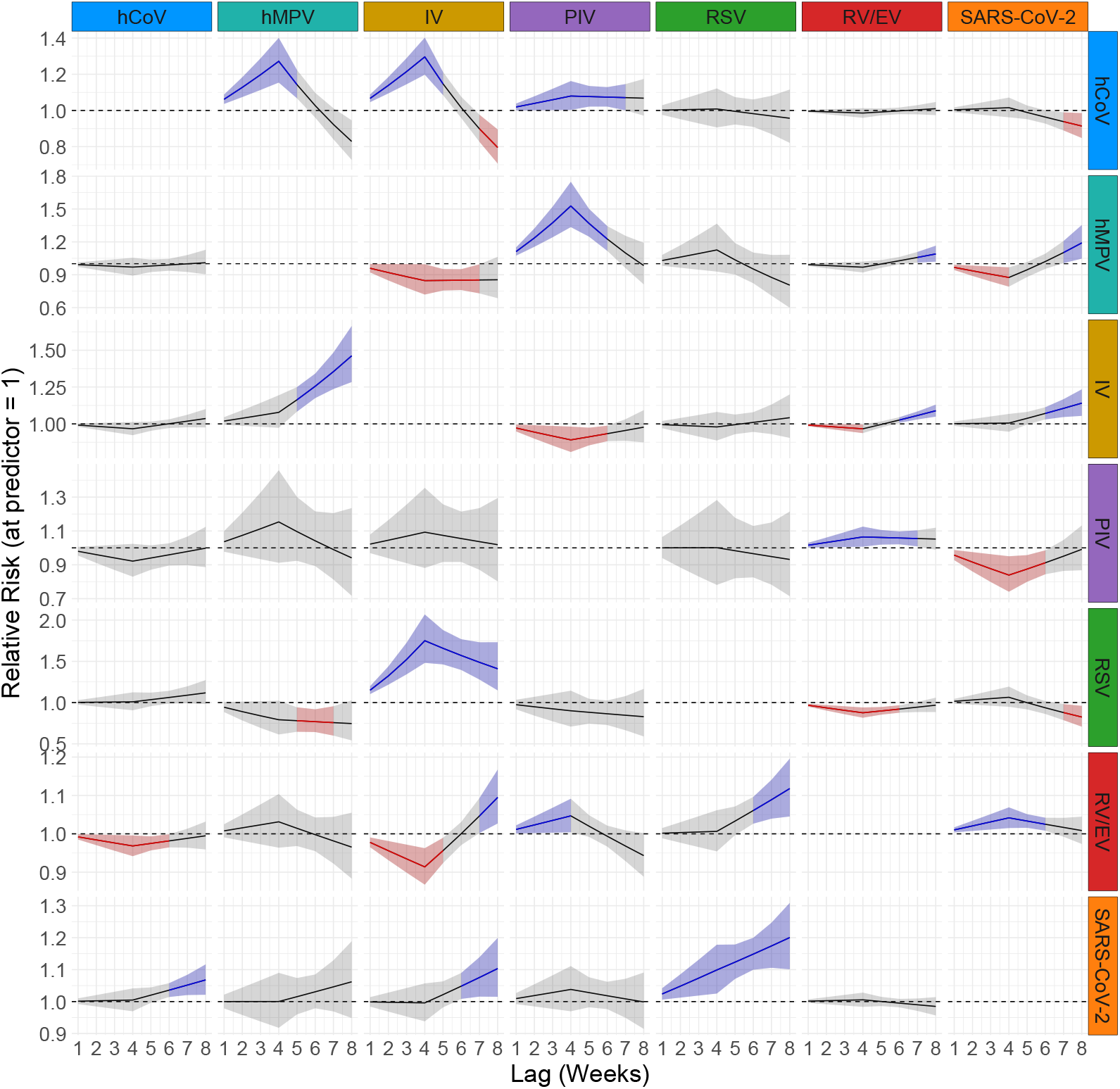
Lag-Response curves from the DLNM. Each panel shows the RR and 95% CI (shaded area) for a target disease (rows) in response to a lagged predictor disease (columns) at a fixed predictor value of 1. The dashed horizontal line indicates RR = 1 (no effect).

Overall, the lag–response curves indicated that some cross-pathogen effects were short-lived and already captured by one-week regressions (Section 3.3), whereas others unfolded gradually across longer lag periods and became detectable only when distributed effects were modelled. Several clear patterns emerged from the DLNM analysis. Largely consistent with the short-term lagged models, cooperative effects were evident for hCoV on IV (peak at lag 4, RR: 1.2962, 95% CI: [1.1979; 1.4025]), hMPV on PIV (peak at lag 4, RR: 1.5276, 95% CI: [1.3350; 1.7479]), and SARS-CoV-2 on RSV (peak at lag 8, RR: 1.2003, 95% CI: [1.1010; 1.3086]). Competitive effects were observed for RV/EV on hCoV (peak at lag 4, RR: 0.9684, 95% CI: [0.9418; 0.9957]) and IV on RV/EV (peak at lag 4, RR: 0.9663, 95% CI: [0.9370; 0.9965]). Additional associations became apparent only in the distributed-lag framework, including competitive effects of hMPV on IV and PIV on SARS-CoV-2. Some effects were detectable only at longer lags, such as a delayed cooperative effect of hMPV on RV/EV. A table with the corresponding estimates can be found in Appendix G.

### 3.6 Subtype-Level Interaction Analysis

While the previous analyses merged subtypes within broader pathogen groups, we further investigated subtype- and lineage-specific interactions for influenza viruses and coronaviruses to assess whether circulation of one strain predicts short- or longer-term changes in another.

For cross-pathogen interactions, the DLNM model revealed cooperative effects of IV A-H1N1 on both other subtypes for lags up to around 5 weeks and of IV A-H3 on subtype B for lags up to 6 weeks. Concerning competition, we found significant effects of IV A-H3 on IV A-H1N1 and of IV B on IV A-H3 for longer lags (Figure 7).

**Figure 7:**
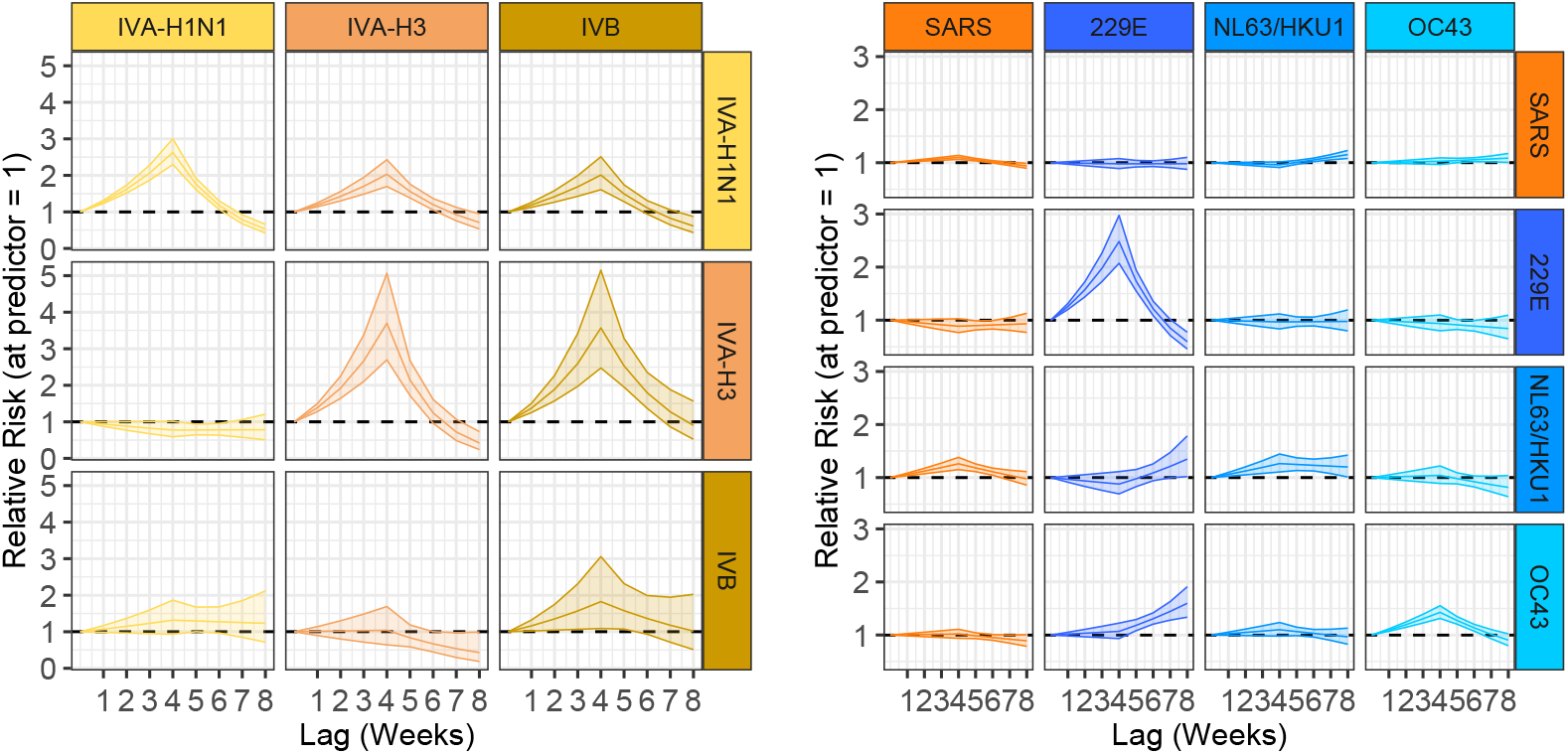
Lag-Response curves from the DLNM. The left plot illustrates interactions among IV subtypes (A-H1N1, A-H3, and B). The right plot displays interactions between SARS-CoV-2 and hCoV subtypes (229E, OC43, NL63/HKU1). Each panel shows the RR and 95% CI (shaded area) for a target disease (rows) in response to a lagged predictor disease (columns) at a fixed predictor value of 1. The dashed horizontal line indicates RR = 1 (no effect).

Regarding coronaviruses, we detected a significant negative effect of hCoV-229E on SARS-CoV-2 for larger lags. Additionally, significant cooperative effects were detected for hCoV-NL63/HKU1 on SARS-CoV-2 for all lags and vice verse for lags exceeding 6 weeks, for hCoV-OC43 on hCoV-229E and SARS-CoV-2 on hCoV-OC43 for lags exceeding 5 weeks (Figure 7).

## 4 Discussion

In this study we explored several statistical modelling approaches to characterise cross-pathogen associations among co-circulating respiratory viruses. We found consistent evidence of both competitive and cooperative interactions at different temporal and spatial scales, with many virus pairs showing significant directional associations (i.e., A→B not necessarily equal to B→A). By combining multiple modelling frameworks, our analysis revealed robust pairwise effects while showing that each framework offers distinct insights into cross-pathogen dynamics, underscoring the value of the ensemble of complementary models for studying cross-pathogen dynamics.

The models used to investigate associations differ in structure, assumptions, and interpretability, thereby providing complementary insights. The results are interpreted along four dimensions capturing distinct aspects of cross-pathogen dynamics: spatial heterogeneity, directionality of associations, context-specific decomposition of baseline and transmission-driven processes, and temporal lag structure. Together, our work provides a comprehensive view of how respiratory viruses coexist and influence one another.

Exploratory analyses, including raw time series, Granger causality tests, and PCA, revealed associations and regional heterogeneity. Granger analysis and PCA captured local temporal dependencies that varied by region, underscoring the need to incorporate spatial aspects into the analyses. Spatial variation was modelled explicitly in the LGMs and GLMMs through region-level random effects. Including regional effects markedly improved fit highlighting the importance of spatial heterogeneity in pathogen incidence. Although our analyses were limited to the NUTS2 level, future work with higher-resolution data could extend the LGM and E&E frameworks to model spatial dependence explicitly through adjacency or distance structures.

Regression frameworks estimated direct short-term lagged effects of disease incidence while adjusting for covariates. The inferred directions of association were consistent across models, though effect sizes and significance occasionally differed. Differences in significance likely reflect the models’ varying sensitivity and structural assumptions, so effects reproduced across multiple frameworks were interpreted as more robust. The VGLM identified the most significant associations and tended to yield larger effect estimates, indicating higher sensitivity but also possible inflation of magnitudes. Associations unique to the VGLM should therefore be interpreted cautiously.

The E&E model indicated that cross-pathogen effects were context-specific, often confined to either the (endemic) or the transmission-related (epidemic) component. Overall, enhancement dominated the detected associations, with most effects expressed through the endemic baseline and fewer through short-term epidemic carryover. Associations limited to the endemic component likely represent shared background drivers such as seasonality or susceptibility, whereas those in the epidemic component capture short-term dependencies linked to concurrent transmission. Although the E&E framework has rarely been used for pathogen interactions, its extension here to include lagged counts of multiple pathogens enabled detection of both cooperative and competitive effects. However, only positive associations were significant, suggesting that suppressive interactions were weak or absorbed by the flexible baseline.

The DLNM showed that associations varied over time, with some emerging only at longer lags. Although results were sensitive to the choice of lag structure, sensitivity analyses yielded largely consistent estimates. Several pathogen pairs displayed asymmetric effects (positive in one direction but negative or absent in the other) reflecting differences in transmission dynamics, immunity, or epidemic timing, though sparse data or residual seasonality may also contribute. These findings highlight that cross-pathogen associations are temporally complex, context-dependent, and not necessarily reciprocal: a pathogen peaking earlier or inducing longer-lasting immunity may exert a stronger apparent influence on another.

Despite differences in model specification, consistent results have been overall obtained across frameworks, supporting the robustness of the inferred temporal patterns. No single framework, however, can capture the full range of mechanisms underlying these associations, which may arise from immunological or behavioural processes. We therefore advocate a tiered strategy: using VGLMs to screen for potential associations and complementing them with DLNMs and E&E models to characterise temporal complexity and distinguish baseline from transmission-driven effects.

Concerning the estimated significant associations, we found, in line with previous work, a recurrent competitive association between IV and RV/EV, with negative effects in all temporal scales (lags up to four weeks), consistent with clinical and experimental evidence for RV/EV-mediated interference of IV [39, 40, 41]. Our results refine these findings by demonstrating that this inhibitory effect persists for several weeks when explicitly modelling temporal lags across multiple seasons. We also observed recurring enhancing effects from SARS-CoV-2 toward hCoV across regression models and in the endemic component of the E&E model as well as in the DLNM for lags over 6 weeks. The DLNM further indicated a subtype-specific pattern: prior hCoV-NL63 incidence showed an enhancing association on SARS-CoV-2 at lags up to seven weeks, whereas prior hCoV-229E incidence showed a competing effect at lags five to seven weeks. These observations sit alongside animal and modelling evidence for cross-protection from hCoV against SARS-CoV-2, including partial protection after 229E/NL63 priming in mice [42] and population-level scenarios where hCoV immunity can reduce SARS-CoV-2 attack rates [14]. Human serology also indicates that pre-existing hCoV antibodies can be back-boosted without protection and may correlate with susceptibility or severity [43, 44]. Taken together, the direction and magnitude of SARS-CoV-2/hCoV interactions likely depend on the hCoV subtype and immune context. For RSV, we found a recurring enhancing effect from SARS-CoV-2 at all time scales and in the E&E endemic component, while the opposite direction appeared as a long-term protective effect in the DLNM at lags 7–8. This pattern is compatible with sequential-infection experiments where RSV→SARS-CoV-2 is protective but SARS-CoV-2→RSV worsens SARS-CoV-2 outcomes [45], and extends these observations to population-level incidence data, suggesting that similar directional asymmetry may operate in natural circulation. For RV/EV with hCoV, our regression and DLNM results suggested competition, whereas the E&E endemic component indicated enhancement. Surveillance from Nepal documented frequent detections of EV/RV and hCoV but did not resolve temporal precedence, so comparisons remain cautious [46]. Finally, we found a consistent enhancing effect of hMPV on PIV across all regression-based models, in the endemic component of the E&E framework, and in the DLNM for lags up to six weeks, further supported by a cosine similarity of 0.636 in the PCA. By contrast, Nickbakhsh et al. [41] reported a negative association between hMPV and PIV in Scottish surveillance data, while Madewell et al. [21] identified a positive association between hMPV and PIV-1 in Puerto Rico. Beyond these reports, the literature provides little direct evidence on hMPV–PIV interactions. Our results therefore contribute population-level, temporally resolved evidence of a consistent enhancing effect that warrants further investigation.

Our models identified the type, direction, and relative strength of cross-pathogen associations but cannot disentangle underlying mechanisms. Short-lag effects may reflect immunological processes (interference, facilitation, trained immunity) or behavioural changes such as reduced contact during illness. Clarifying these mechanisms will require the analysis of individual-level data combining behavioural and immunological information.

The data used in these analyses have several limitations that may affect the strength and generalisability of the observed associations. Infectieradar is a participatory surveillance system and therefore not representative of the general population. Underrepresentation of certain age groups and uneven regional participation may influence estimated associations, particularly for pathogens concentrated in specific subpopulations or areas. Additional data would help assess potential biases from this non-representative sampling. Testing intensity also varied across weeks and regions, and preferential inclusion of participants with positive SARS-CoV-2 self-tests in earlier seasons may have inflated synchrony in some areas. Because the number of weekly laboratory samples was capped, low-incidence periods may be overrepresented, dampening seasonal amplitudes and reducing contrasts across pathogens. Differences in test sensitivity may further contribute to pathogen-specific underdetection. These limitations likely introduce random measurement error that attenuates rather than generates associations. To limit such effects, autoregressive terms and regional random effects were included in the models, and seasonal sensitivity analyses were performed. As data were analysed across three epidemiological seasons, the estimated associations reflect average effects over time. Season-specific analyses yielded broadly consistent results, though the strength and significance of some associations varied, suggesting that interaction patterns depend on the prevailing epidemiological context. However, properly quantifying potential biases introduced by the study design will require additional data and dedicated analyses.

With this study, we provided an exploration of cross-pathogen interactions among a selection of respiratory viruses through diverse statistical methodologies applied to complex epidemiological data. Our findings reveal short- and long-term predictive effects, providing a richer picture of the patterns of pathogen interaction. The observed spatiotemporal and seasonal variability in these interactions highlights the potential need for dynamic and context-specific public health interventions and underscores the value of combining multiple modelling approaches for robust epidemiological insights. Beyond the specific context of respiratory pathogens, the integrated statistical methodology employed in this study, offers a powerful and adaptable framework that could be broadly applied to other biological processes involving dynamic interactions between different entities, such as in ecological population dynamics or inter-species competition.

## Data Availibility

The full data supporting the findings of this study are retained at the Dutch National Institute for Public Health and the Environment (RIVM), Bilthoven, and will not be made openly accessible due to ethical and privacy concerns. Data can, however, be made available after approval of a motivated and written request to the RIVM at.

## Acknowledgement/Funding

We thank Prof. Andrew B. Lawson for constructive feedback on the model specification of the Latent Gaussian model. We thank the participants of Infectieradar for their weekly contributions and submission of samples. Without their contribution this project won’t have been a success. N.H. was supported by the European Union’s Horizon Europe programme under the VERDI project (grant agreement No. 101045989). Views and opinions expressed are those of the authors only and do not necessarily reflect those of the European Union or the Health and Digital Executive Agency (HADEA). Neither the European Union nor the granting authority can be held responsible for them.

## Authors’ contributions

AT, NB, DE and AJVH contributed to conceptualization of the study. GC, EK, WH, DE and AJVH contributed to acquisition of the data. LH handled project administration. AT, NB, DP, SA, GM, NH and CF contributed to the methodology and interpretation of the results. NB performed the analysis. NB and AT drafted the manuscript. All co-authors critically reviewed the manuscript.

