## Supplementary Material for "Population-Level Associations in the Spread of Co-Circulating Respiratory Viruses: A Multi-Method Statistical Investigation Using Incidence Data"

### A Exploratory Data Analysis

This section provides supplementary figures and tables supporting the exploratory graphical inspection described in the main text.

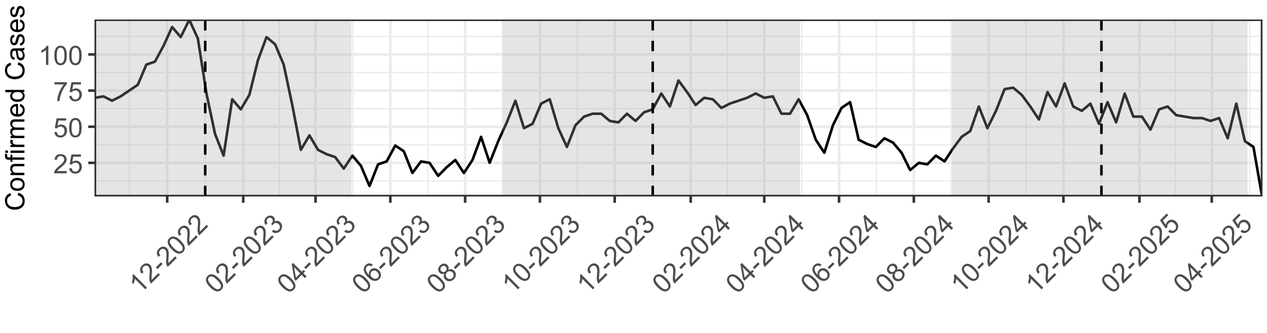


Figure A.1: Weekly confirmed cases aggregated across all pathogens and regions per week, from 4 October 2022 to 12 May 2025. Vertical dashed lines indicate the start of each calendar year. Shaded areas indicate epidemiological seasons.

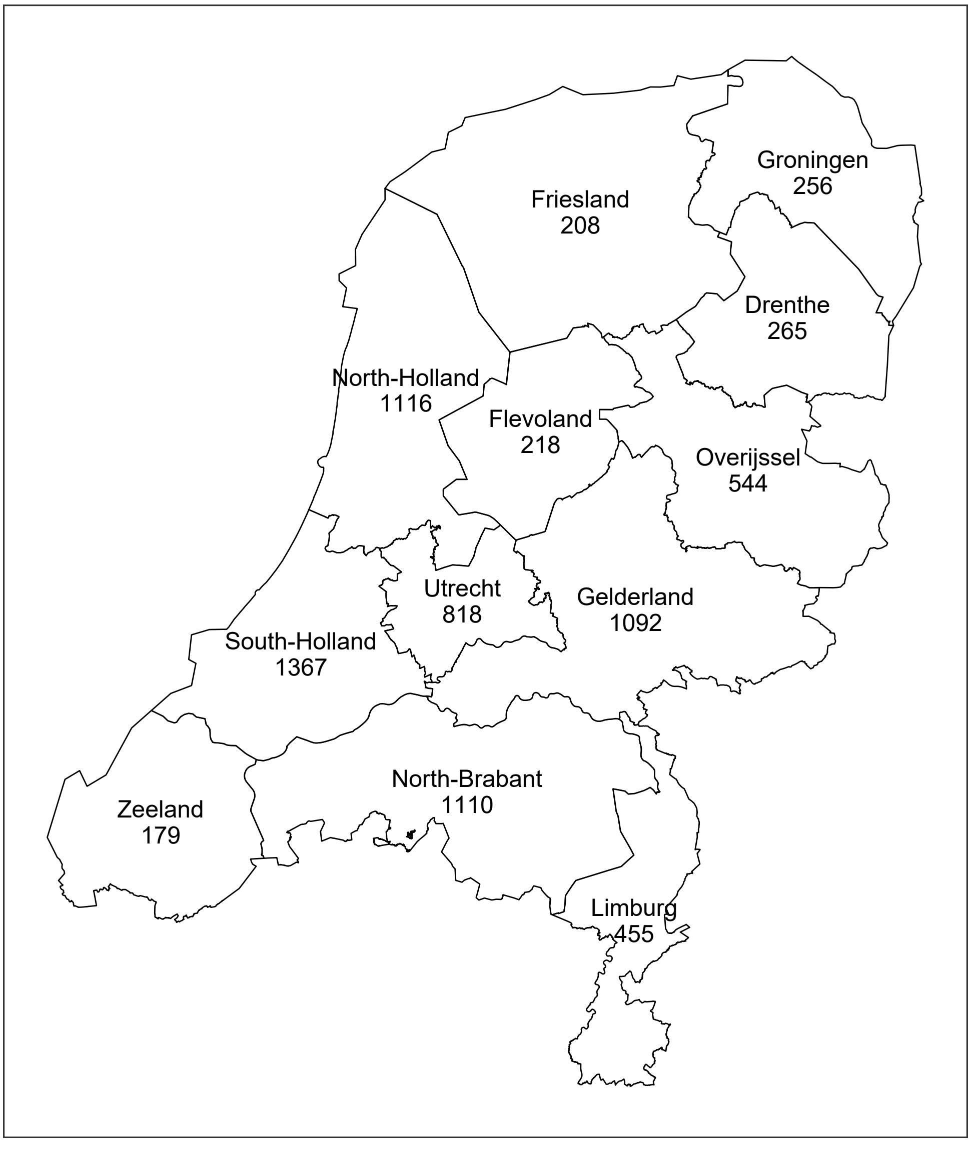


Figure A.2: Aggregated incidence across all pathogens and the entire study period, stratified by region.

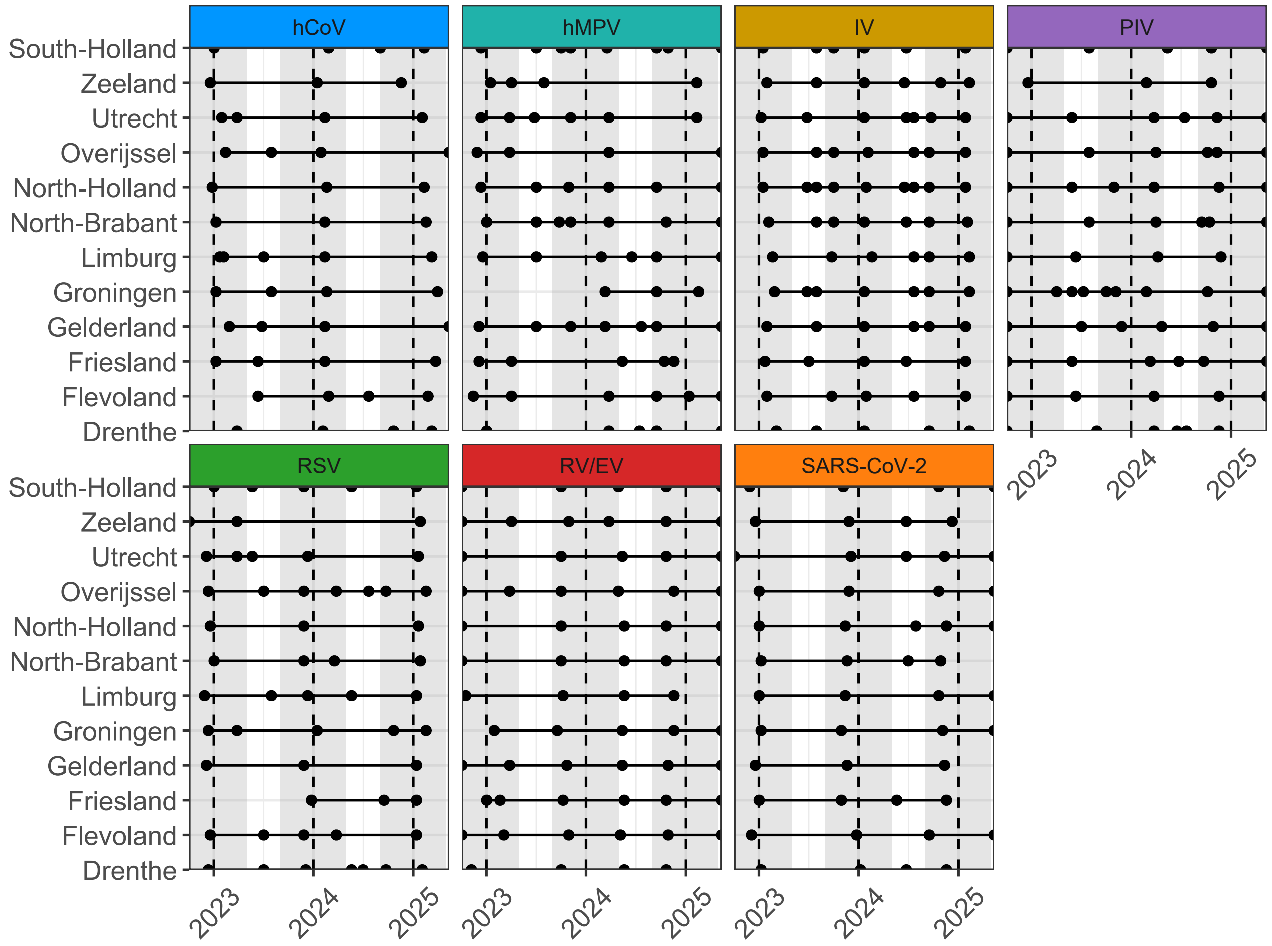


Figure A.3: Temporal and spatial distribution of the timing of local maxima (peaks) in the smoothed weekly incidence curves by disease.

Table A.1: Descriptive statistics for weekly confirmed cases by week–region combination (minimum, mean, maximum, and proportion of zero-weeks) and total confirmed cases for each of the seven diseases.

| | **By Week-Region Combination** | | | | |
|---|---|---|---|---|---|
| **Disease** | **Minimum** | **Mean** | **Maximum** | **Zero-weeks** | **Total** |
| RV/EV | 0 | 1.1 | 11 | 50.7% | 3453 |
| SARS-CoV-2 | 0 | 0.42 | 20 | 74.9% | 1395 |
| hCoV | 0 | 0.36 | 8 | 87.3% | 1172 |
| IV | 0 | 0.19 | 9 | 88.3% | 619 |
| PIV | 0 | 0.13 | 5 | 89.4% | 412 |
| hMPV | 0 | 0.95 | 5 | 92.7% | 312 |
| RSV | 0 | 0.08 | 4 | 93% | 265 |

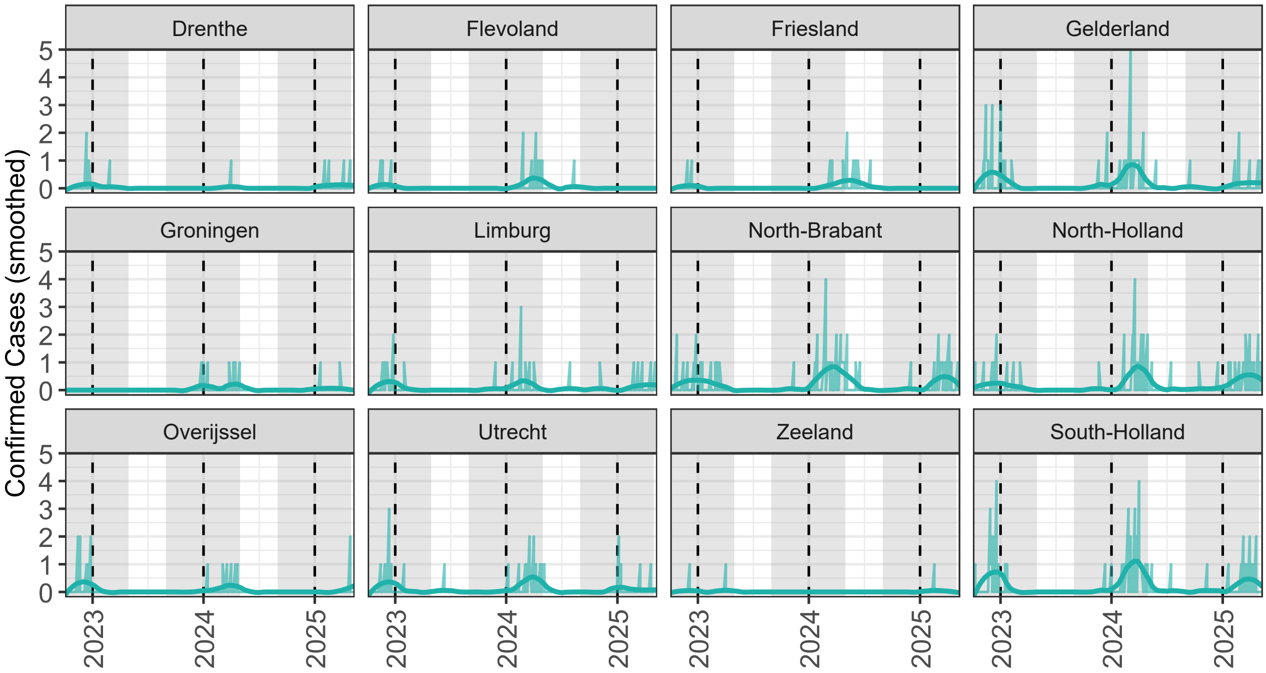


Figure A.4: Weekly hMPV incidence by region from 4 October 2022 to 12 May 2025. The light bars represent the raw weekly case counts, while the darker line shows the smoothed incidence trend (estimated using a LOESS smoother with span = 0.2). Vertical dashed lines indicate the start of each calendar year. Shaded areas indicate epidemiological seasons.

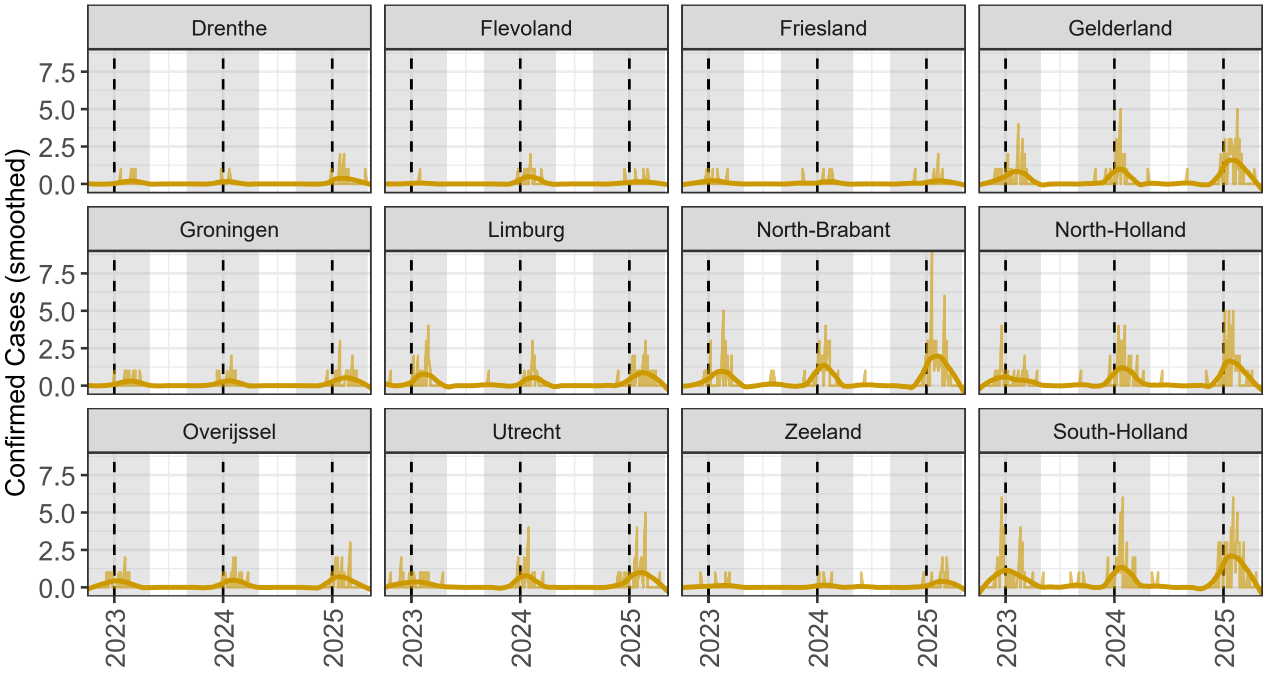


Figure A.5: Weekly IV incidence by region from 4 October 2022 to 12 May 2025. The light bars represent the raw weekly case counts, while the darker line shows the smoothed incidence trend (estimated using a LOESS smoother with span = 0.2). Vertical dashed lines indicate the start of each calendar year. Shaded areas indicate epidemiological seasons.

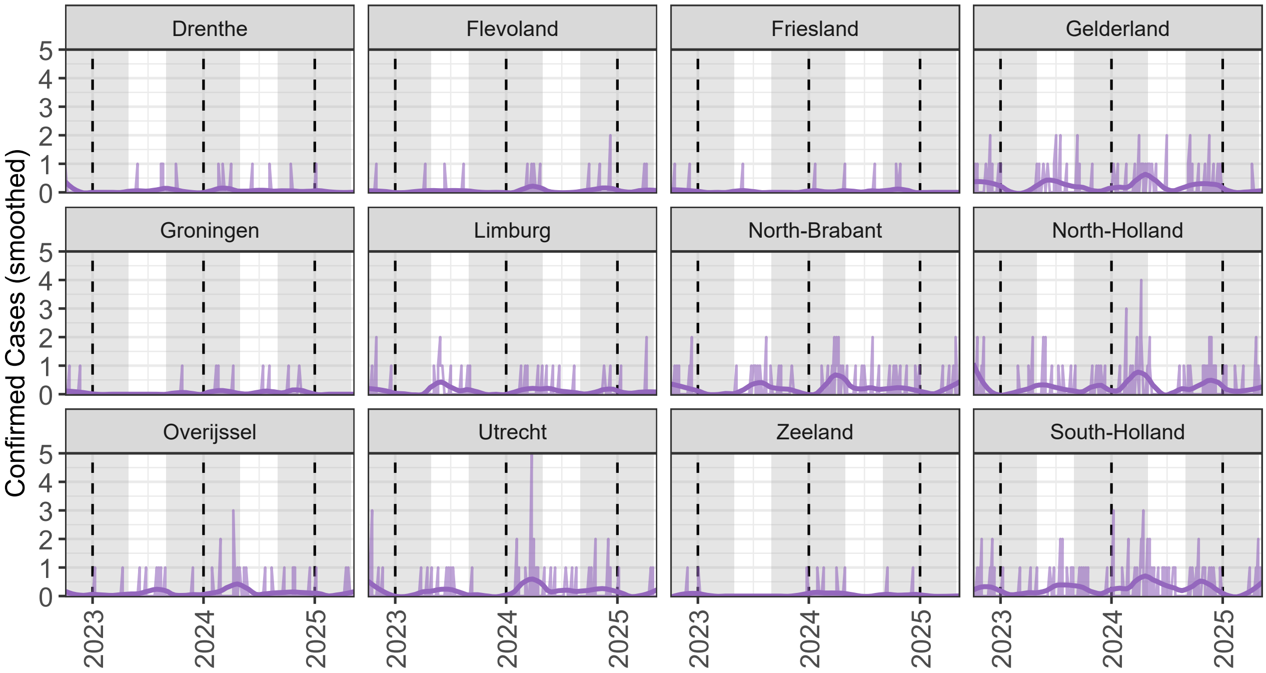


Figure A.6: Weekly PIV incidence by region from 4 October 2022 to 12 May 2025. The light bars represent the raw weekly case counts, while the darker line shows the smoothed incidence trend (estimated using a LOESS smoother with span = 0.2). Vertical dashed lines indicate the start of each calendar year. Shaded areas indicate epidemiological seasons.

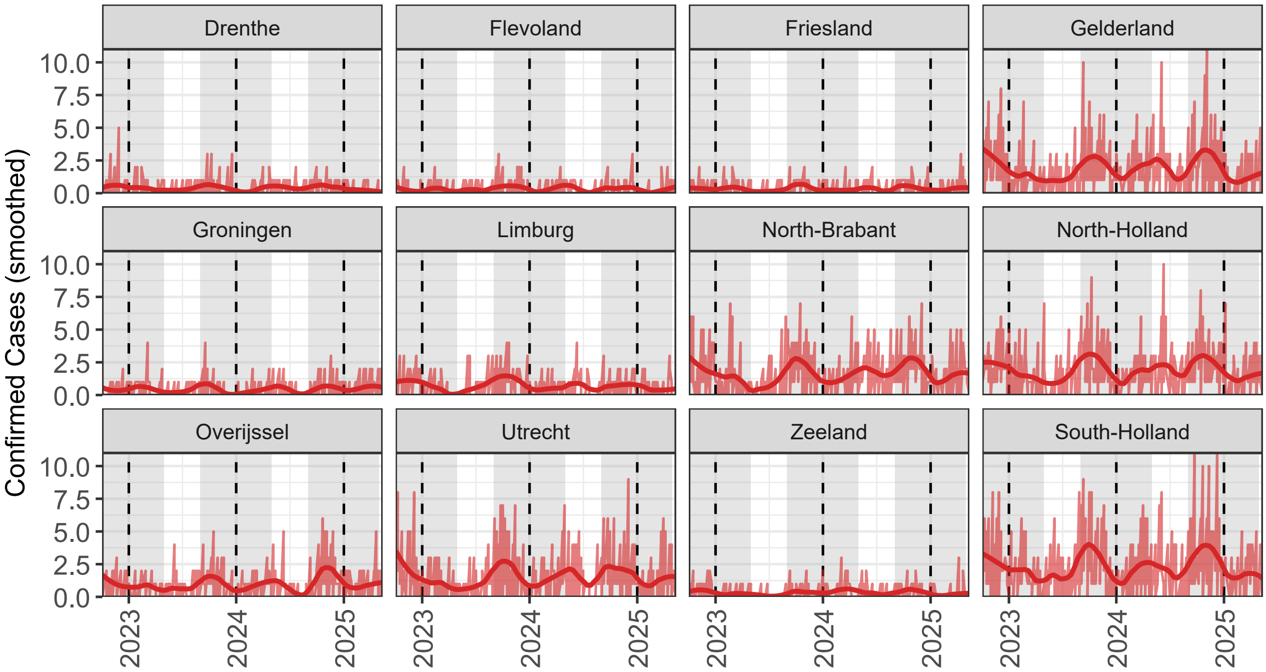


Figure A.7: Weekly RV/EV incidence by region from 4 October 2022 to 12 May 2025. The light bars represent the raw weekly case counts, while the darker line shows the smoothed incidence trend (estimated using a LOESS smoother with span = 0.2). Vertical dashed lines indicate the start of each calendar year. Shaded areas indicate epidemiological seasons.

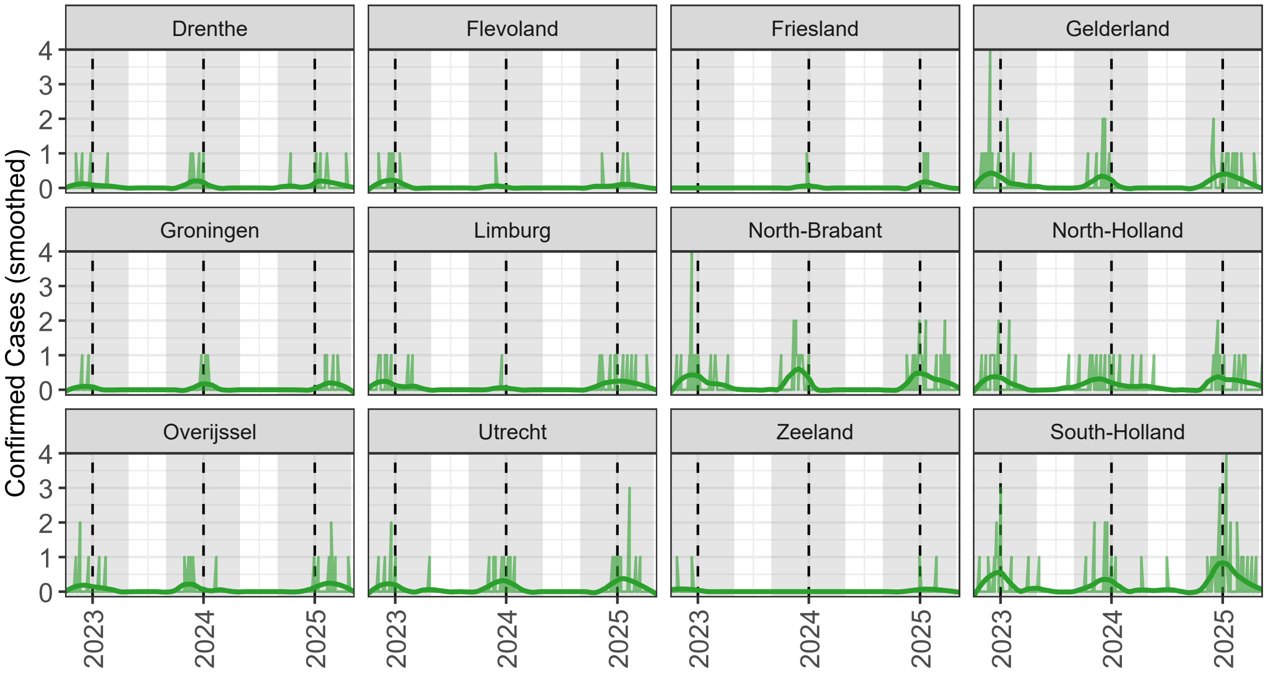


Figure A.8: Weekly RSV incidence by region from 4 October 2022 to 12 May 2025. The light bars represent the raw weekly case counts, while the darker line shows the smoothed incidence trend (estimated using a LOESS smoother with span = 0.2). Vertical dashed lines indicate the start of each calendar year. Shaded areas indicate epidemiological seasons.

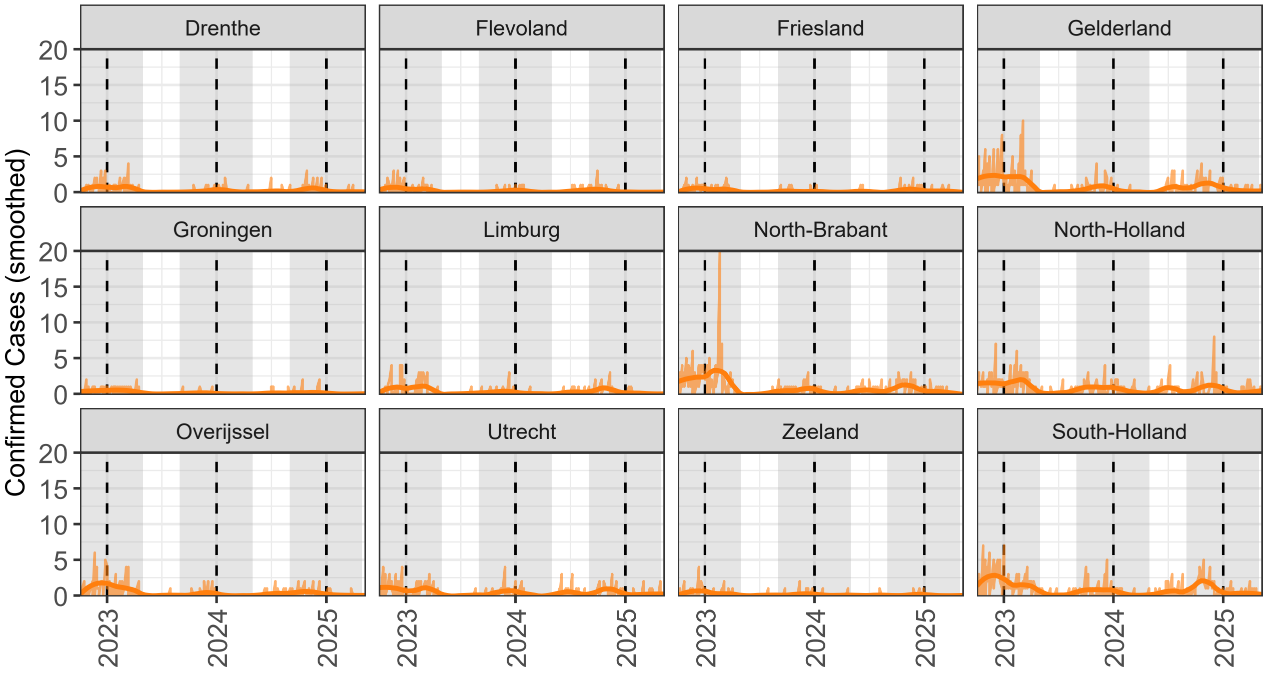


Figure A.9: Weekly SARS-CoV-2 incidence by region from 4 October 2022 to 12 May 2025. The light bars represent the raw weekly case counts, while the darker line shows the smoothed incidence trend (estimated using a LOESS smoother with span = 0.2). Vertical dashed lines indicate the start of each calendar year. Shaded areas indicate epidemiological seasons.

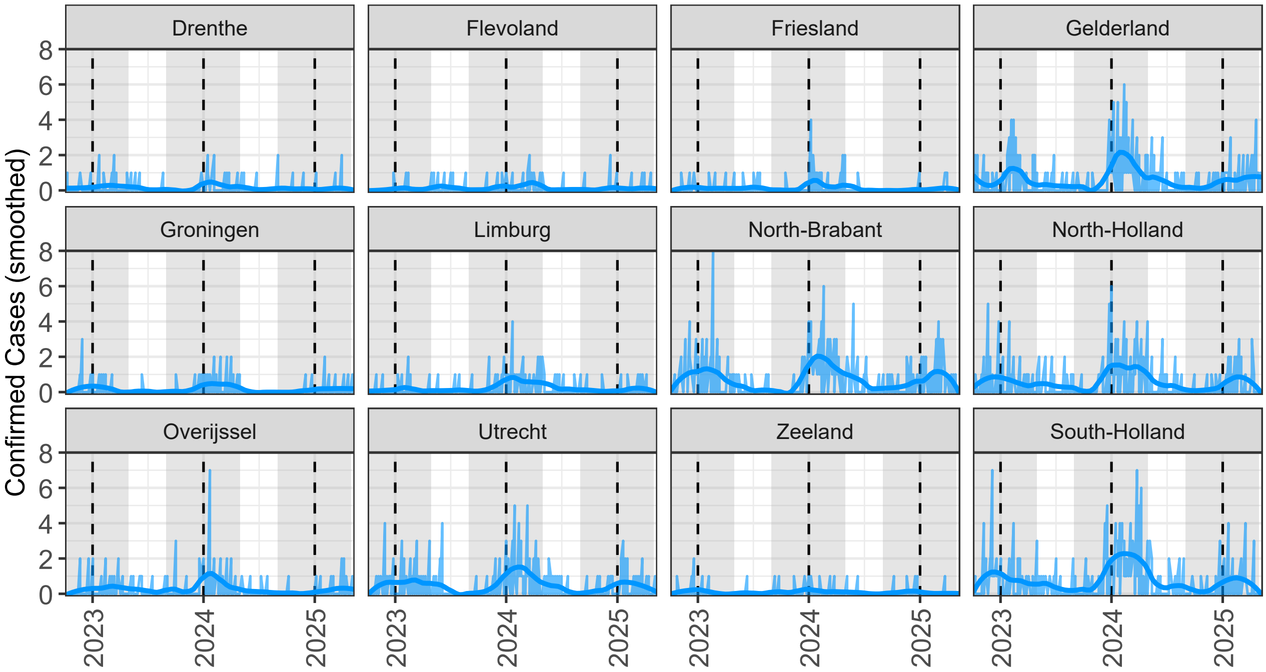


Figure A.10: Weekly hCoV incidence by region from 4 October 2022 to 12 May 2025. The light bars represent the raw weekly case counts, while the darker line shows the smoothed incidence trend (estimated using a LOESS smoother with span = 0.2). Vertical dashed lines indicate the start of each calendar year. Shaded areas indicate epidemiological seasons.

### B Granger Causality

Data for each geographical region were processed independently for region-specific Granger analyses, while for the national-level analysis, data were aggregated across all regions.

#### B.1 Preprocessing of Time Series

To ensure comparability between national and regional Granger analyses, identical preprocessing was applied to all pathogen time series.

**Aggregation level** For the national analysis, weekly case counts were aggregated across all regions to obtain one series per pathogen. For the regional analysis, the same steps were applied separately within each region.

**Variance stabilisation** To reduce skewness and stabilise variance, we applied a log transformation with an offset of 1 (to avoid issues with zero counts) to the weekly count $y_{d,t}$: $z_t = log(1 + y_{d,t})$.

**Seasonal differencing** Respiratory pathogens typically exhibit strong annual cycles. To remove this seasonality, we computed a 52-week lagged difference: $z_t^{(s)} = z_t - z_{t-52}$. This isolates intra-annual variation by cancelling out recurring yearly patterns.

**First-order differencing** o remove remaining low-frequency trends and seasonal shifts in mean level, we applied a first-order difference to the seasonally adjusted series: $z_t^{(sf)} = z_t^{(s)} - z_{t-1}^{(s)}$.

**Final transformation** Combining the above steps, the fully transformed time series used for modelling is:

$$z_t^{(sf)} = \log\left(\frac{(1 + y_{d,t})(1 + y_{d,t-53})}{(1 + y_{d,t-52})(1 + y_{d,t-1})}\right).$$

This transformation yielded approximately stationary, variance-stabilised series for all pathogens.

**Exclusion criterion** Pathogen–region combinations with fewer than 10 non-zero weeks were excluded from the regional analysis.

#### B.2 Lag Order Selection

The optimal lag length for the vector autoregressive (VAR) models was determined using information criteria to balance model fit and parsimony. For both the national and regional analyses, lag order selection compared the AIC and BIC. Across all regions, AIC consistently favoured the maximum tested lag ($p = 8$), whereas BIC uniformly selected the minimum lag ($p = 1$). Because BIC imposes a stronger penalty on over-parametrisation and performs more conservatively in small samples, we adopted a single-lag specification ($p = 1$) for all models. This choice ensured comparability between national and regional VAR systems, controlled overfitting, and simplified interpretation of Granger causality results.

#### B.3 Mathematical Formulation of Granger Causality

To assess potential causal interactions between pathogens, Granger causality was assessed within a multivariate VAR framework, applied either to the aggregated national time series or to region-specific time series.

Let $\boldsymbol{y}_t = (y_{1,t}, y_{2,t}, \ldots, y_{D,t})^\top$ denote the joint observation of the $D$ stationary pathogen time series at time $t$. The system was modelled as a first-order vector autoregressive process:

$$\boldsymbol{y}_t = \mathbf{A}_1 \boldsymbol{y}_{t-1} + \boldsymbol{\varepsilon}_t,$$

where $\mathbf{A}_1 \in \mathbb{R}^{D \times D}$ is the matrix of autoregressive coefficients and $\boldsymbol{\varepsilon}_t$ is a zero-mean white-noise process with covariance matrix $\boldsymbol{\Sigma}$. To test whether pathogen $d'$ Granger-causes pathogen $d$, we evaluate the null hypothesis that the lagged coefficient of $y_{d',t-1}$ in the equation for $y_{d,t}$ equals zero $H_0 : a_{d,d'} = 0$. A small $p$-value indicates rejection of $H_0$, implying that past values of pathogen $d'$ provide predictive information about pathogen $d$, conditional on all other pathogens in the system.

The national VAR system included one transformed time series per pathogen, whereas each regional VAR included one series per pathogen–region combination. All Granger causality tests were conducted pairwise while conditioning on all other series in the respective system, thereby reducing the risk of spurious associations that can arise in bivariate settings.

#### B.4 Model Assumption Checks

Before interpreting Granger causality results, we verified key assumptions of the VAR framework at both the national and regional levels: weak stationarity, linear specification, and absence of strong residual autocorrelation or instability.

Stationarity was assessed using two complementary tests. The Augmented Dickey–Fuller (ADF) test examines the null hypothesis of a unit root (non-stationarity), whereas the Kwiatkowski–Phillips–Schmidt–Shin (KPSS) test evaluates the null hypothesis of stationarity. Using both allows for a more robust assessment: rejecting the ADF null while not rejecting the KPSS null provides stronger evidence of stationarity than either test alone.

Linearity of the VAR equations was evaluated using the Ramsey Regression Equation Specification Error Test (RESET). This test examines whether higher-order powers of the fitted values (e.g., squared or cubic terms) significantly improve model fit, which would indicate model misspecification or unaccounted nonlinear relationships. Failing to reject the null hypothesis implies that the assumed linear structure of the VAR equations is adequate for the data.

Residual independence and model stability were assessed to ensure valid statistical inference from the VAR models. Residual diagnostics tested whether remaining autocorrelation could bias the Granger causality estimates, as correlated residuals may indicate omitted dynamics or an insufficient lag structure. We applied the Portmanteau test for serial correlation up to lag 16, which evaluates the joint null hypothesis that residual autocorrelations across all lags are zero. Failure to reject this null supports the assumption that residuals approximate white noise.

Model stability was examined by inspecting the roots of the characteristic polynomial of each VAR system. Stability requires that all root moduli lie strictly within the unit circle, ensuring that shocks to the system dissipate over time rather than accumulating. This condition guarantees that the VAR process is dynamically stable and that estimated Granger causality relations reflect genuine predictive structure rather than model instability.

##### B.4.1 National-level Checks

**Stationarity** All seven national-level pathogen series satisfied the stationarity requirement. The ADF test yielded $p < 0.05$ for all series except RV/EV ($p = 0.085$), while the KPSS test returned $p > 0.05$ for all series. This concordant pattern indicates that the transformed series were stationary after seasonal and first-order differencing. No further adjustment was required.

**Linearity** All $p$-values from the RESET test exceeded 0.05, indicating that the linear specification of the national VAR model was adequate.

**Residual diagnostics** At the national level, the Portmanteau test (up to lag 16) found no evidence of residual autocorrelation ($p = 0.9916$), indicating that model residuals approximated white noise and that the lag specification adequately captured short-term dynamics.

**Model stability** All root moduli were well below 1 (range: $0.18 - 0.55$), confirming that the national VAR model was dynamically stable and satisfied the stationarity assumption.

##### B.4.2 Regional-level Checks

**Stationarity** All transformed pathogen time series satisfied the stationarity requirements after applying seasonal and first-order differencing, according to both the ADF test ($p < 0.05$) and the KPSS test ($p > 0.05$)

for the majority of series. A single exception was observed for RV/EV ($p_{ADF} = 0.0852$). However, as the KPSS test did not reject the null hypothesis of stationarity and visual inspection showed stable variance and mean, this series was considered sufficiently stationary for analysis. To maintain methodological consistency and comparability across pathogens, no additional differencing was applied.

**Linearity** Most $p$-values from the RESET test exceeded 0.05, indicating adequate linear specification across the regional VAR models. A small subset showed borderline evidence of nonlinearity (Table B.1). In some pathogen–region combinations, no $p$-value could be computed because the pathogen was nearly absent in that region. These cases were excluded from the RESET analysis but did not affect the overall estimation, which proceeded on the available pathogens within each region.

Table B.1: RESET $p$-values for each pathogen in the regional Granger VAR models. Values below 0.05 are shown in bold.

| | **hMPV** | **IV** | **PIV** | **RV/EV** | **RSV** | **SARS-CoV-2** | **hCoV** |
|---|---|---|---|---|---|---|---|
| **Drenthe** | – | 0.101 | 0.357 | 0.865 | 0.351 | 0.794 | 0.425 |
| **Flevoland** | 0.540 | **0.007** | **0.000** | 0.258 | – | 0.191 | 0.431 |
| **Friesland** | 0.112 | **0.000** | – | 0.949 | – | 0.956 | **0.023** |
| **Gelderland** | 0.553 | 0.715 | 0.922 | 0.592 | 0.068 | 0.949 | 0.681 |
| **Groningen** | – | 0.212 | 0.067 | 0.843 | – | **0.047** | 0.748 |
| **Limburg** | 0.369 | 0.864 | 0.902 | 0.229 | **0.043** | 0.787 | 0.391 |
| **North-Brabant** | 0.611 | 0.753 | 0.584 | 0.374 | 0.111 | 0.594 | 0.466 |
| **North-Holland** | **0.050** | 0.063 | 0.527 | 0.270 | 0.224 | 0.327 | 0.677 |
| **Overijssel** | 0.104 | 0.171 | 0.528 | 0.103 | 0.597 | 0.120 | 0.066 |
| **Utrecht** | 0.718 | 0.151 | 0.283 | 0.940 | 0.547 | 0.244 | 0.504 |
| **Zeeland** | – | **0.000** | – | 0.224 | – | **0.013** | 0.578 |
| **South-Holland** | 0.842 | 0.186 | 0.981 | 0.975 | 0.384 | 0.997 | 0.607 |

**Residual Diagnostics** At the regional level, Portmanteau tests (up to lag 16) indicated some residual autocorrelation across models. To evaluate this further, we inspected the autocorrelation functions (ACFs) of the residuals by pathogen and region (Figure B.1). Although individual ACF values occasionally exceeded the 95% confidence bounds at lower lags, the overall magnitude of autocorrelation remained modest. Most bars fell within or only slightly outside the bounds, and no systematic patterns were observed, supporting the adequacy of the chosen lag structure for the regional VAR models.

**Model stability** All root moduli were strictly less than one, with maximum values ranging from 0.39 in Limburg to 0.81 in South-Holland. Most root distributions clustered between 0.50 and 0.70, confirming that all regional VAR models were dynamically stable and met the required condition for reliable inference.

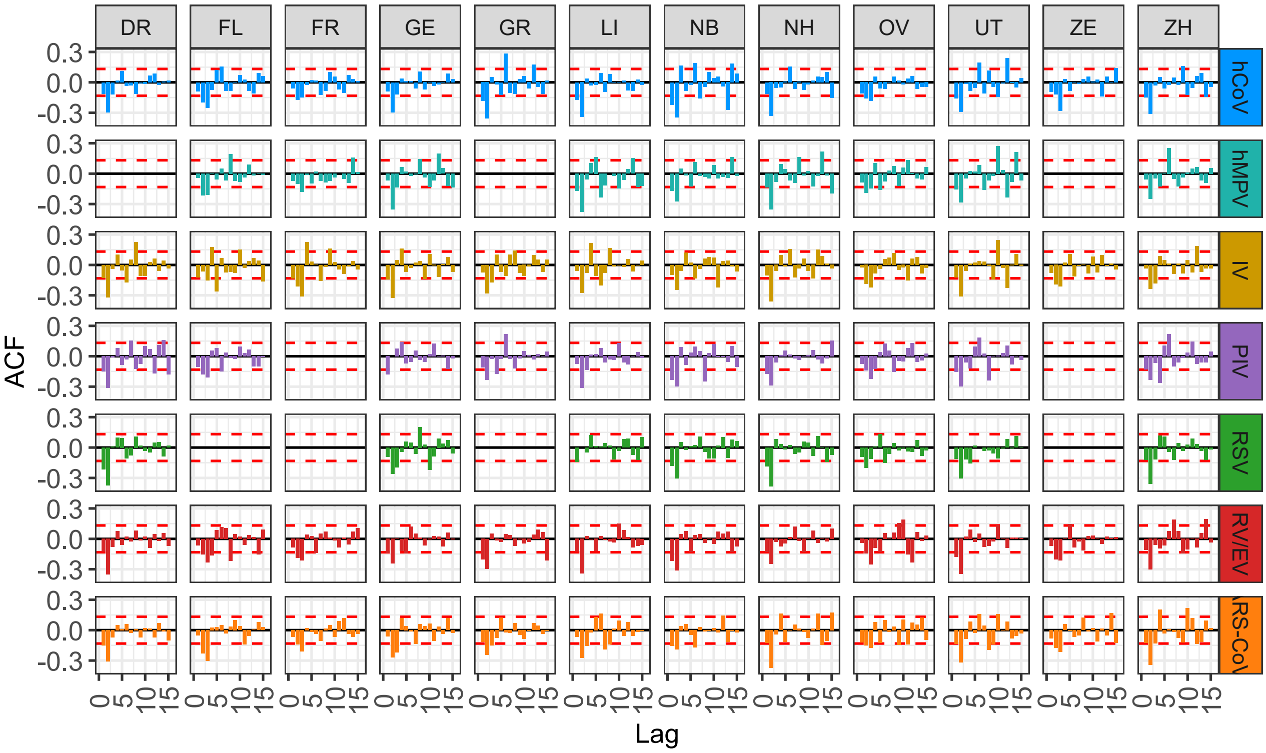


Figure B.1: ACFs of residuals from the regional VAR models, stratified by disease and region. Dashed red lines indicate approximate 95% confidence bounds. Abbreviations of Dutch provinces: DR = Drenthe, FL = Flevoland, FR = Friesland, GE = Gelderland, GR = Groningen, LI = Limburg, NB = North-Brabant, NH = North-Holland, OV = Overijssel, UT = Utrecht, ZE = Zeeland, ZH = South-Holland.

#### B.5 Additional Results

The following supplementary results provide detailed outputs of the Granger causality analyses at the national and regional levels.

##### B.5.1 National-level Analysis

Pairwise $p$-values from the national VAR model are presented in Table B.2. Only a few pathogen pairs show significant predictive relationships, reflecting limited evidence of strong directional effects at the aggregated level.

Table B.2: Pairwise $p$-values from the national (aggregated) Granger causality analysis. Values in bold indicate statistically significant associations ($p < 0.05$).

| | Target Disease | | | | | | |
|---|---|---|---|---|---|---|---|
| **Predictive Disease** | **hMPV** | **IV** | **PIV** | **RSV** | **RV/EV** | **SARS-CoV-2** | **hCoV** |
| **hMPV** | – | **0.0074** | 0.679 | 0.0991 | 0.9076 | 0.1838 | 0.5571 |
| **IV** | **0.0114** | – | 0.1569 | 0.3163 | 0.4749 | 0.2225 | 0.7245 |
| **PIV** | 0.5822 | 0.4081 | – | 0.2359 | 0.2953 | 0.9607 | 0.5672 |
| **RSV** | 0.3501 | 0.6551 | 0.1844 | – | 0.2154 | 0.8691 | 0.2201 |
| **RV/EV** | 0.7330 | 0.1509 | 0.8465 | 0.6965 | – | 0.7466 | 0.4699 |
| **SARS-CoV-2** | 0.9049 | 0.1374 | **0.0429** | 0.2980 | 0.8309 | – | 0.9253 |
| **hCoV** | 0.2236 | 0.4828 | 0.3845 | 0.9991 | 0.8942 | 0.4846 | – |

#### B.6 Regional-level Analysis

Regional analyses reveal greater heterogeneity. Figure 2 shows regional Granger causality matrices, and Figure B.2 summarises how frequently each association was significant across provinces. Together, these plots highlight which pathogen interactions were most consistent spatially.

Table B.3: Proportion of regions in which each Granger causality test was significant ($p < 0.05$) relative to the number of regions for which an assessment was possible.

| | Target Disease | | | | | | |
|---|---|---|---|---|---|---|---|
| **Predictive Disease** | **hMPV** | **IV** | **PIV** | **RSV** | **RV/EV** | **SARS-CoV-2** | **hCoV** |
| **hMPV** | – | 0.22 | 0.38 | 0.000 | 0.11 | 0.11 | 0.11 |
| **IV** | 0.22 | – | 0.20 | 0.38 | 0.17 | 0.08 | 0.17 |
| **PIV** | 0.12 | 0.00 | – | 0.00 | 0.10 | 0.00 | 0.10 |
| **RSV** | 0.14 | 0.50 | 0.12 | – | 0.12 | 0.00 | 0.25 |
| **RV/EV** | 0.00 | 0.00 | 0.10 | 0.25 | – | 0.00 | 0.08 |
| **SARS-CoV-2** | 0.00 | 0.08 | 0.10 | 0.38 | 0.00 | – | 0.08 |
| **hCoV** | 0.11 | 0.08 | 0.10 | 0.25 | 0.00 | 0.00 | – |

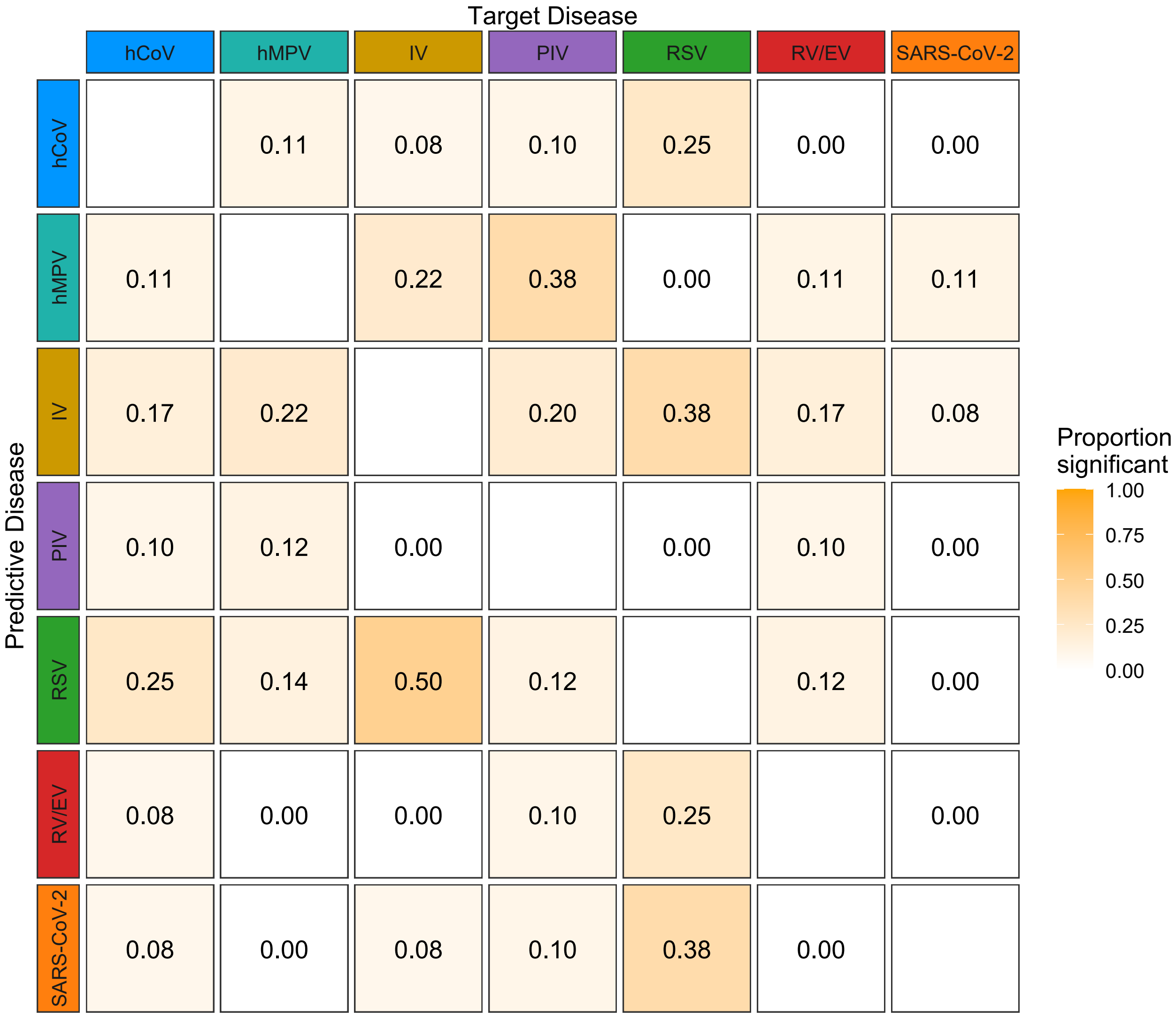


Figure B.2: Proportion of regions with significant Granger causality tests ($p < 0.05$).

#### B.7 Sensitivity Analysis: Pairwise Granger Causality

To assess the robustness of the multivariate VAR results, we repeated the Granger causality tests using a purely pairwise approach. Unlike the main analysis, which controlled for confounding by including all pathogens jointly, this approach evaluated each pathogen pair in isolation without adjustment for the remaining series. It therefore represents a more permissive test, expected to detect additional (but potentially spurious) associations.

All time series were preprocessed identically to the main analysis (log transformation, seasonal differencing, and first-order differencing). Pairwise tests were conducted at lag 1.

##### National-level Results

Pairwise tests at the national level revealed several significant associations (Table B.4). Some overlap was observed with the multivariate results, but additional associations appeared when no adjustment for other pathogens was made, illustrating the effect of omitted-variable confounding.

Table B.4: Pairwise $p$-values from the univariate national Granger causality analysis. Values in bold indicate statistically significant associations ($p < 0.05$).

| | | **Target Disease** | | | | | | |
|---|---|---|---|---|---|---|---|---|
| | | **hMPV** | **IV** | **PIV** | **RSV** | **RV/EV** | **SARS-CoV-2** | **hCoV** |
| **Predictive Disease** | **hMPV** | – | $<$ **0.0001** | **0.0021** | 0.1716 | 0.9886 | 0.2395 | 0.4453 |
| | **IV** | **0.0193** | – | 0.1679 | 0.616 | 0.3725 | 0.3406 | 0.5262 |
| | **PIV** | 0.974 | 0.4541 | – | 0.2903 | 0.3913 | 0.9000 | 0.6768 |
| | **RSV** | 0.7515 | 0.8031 | 0.2242 | – | 0.1234 | 0.8707 | 0.1515 |
| | **RV/EV** | 0.8433 | 0.5897 | 0.2887 | 0.6029 | – | 0.7282 | 0.6221 |
| | **SARS-CoV-2** | 0.9197 | **0.0121** | **0.0010** | 0.759 | 0.5233 | – | 0.9747 |
| | **hCoV** | 0.2294 | 0.3578 | 0.2516 | 0.8675 | 0.8741 | 0.5834 | – |

##### Regional-level Results

Pairwise Granger causality tests were also applied independently within each region to identify direct, bivariate associations between pathogens. Figure B.4 displays the regional matrices of pairwise $p$-values, and Figure **??** summarises the proportion of regions with significant associations. As expected, the pairwise approach yielded more frequent significant links than the multivariate analysis, reflecting the absence of adjustment for indirect dependencies.

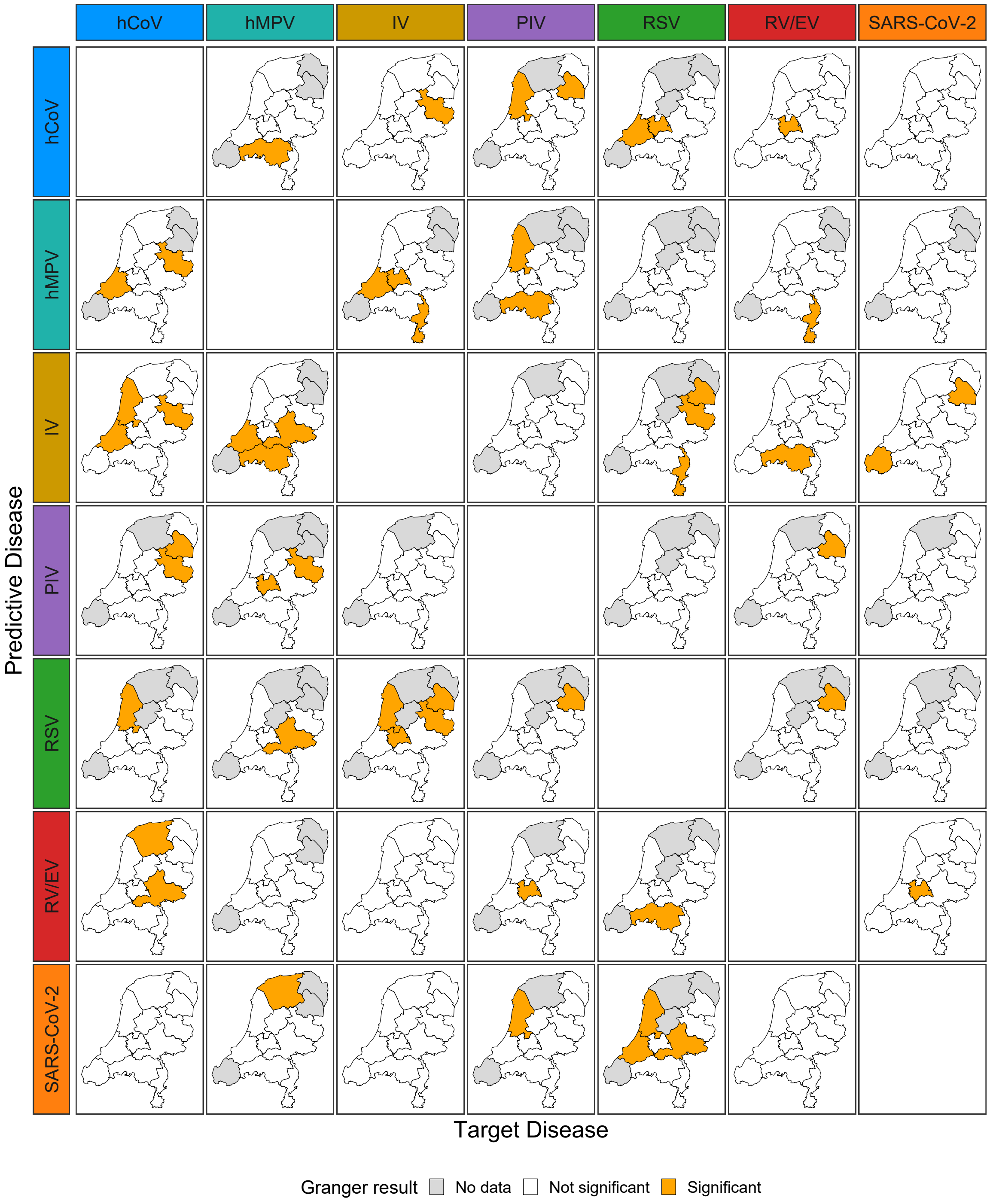


Figure B.3: Regional pairwise Granger causality analysis (sensitivity analysis). Each panel shows the Netherlands map for a given predictive (row) and target (column) pathogen pair. Provinces are colored according to Granger test results: orange for statistically significant associations ($p < 0.05$), white for non-significant results, and gray where no or too little data were available.

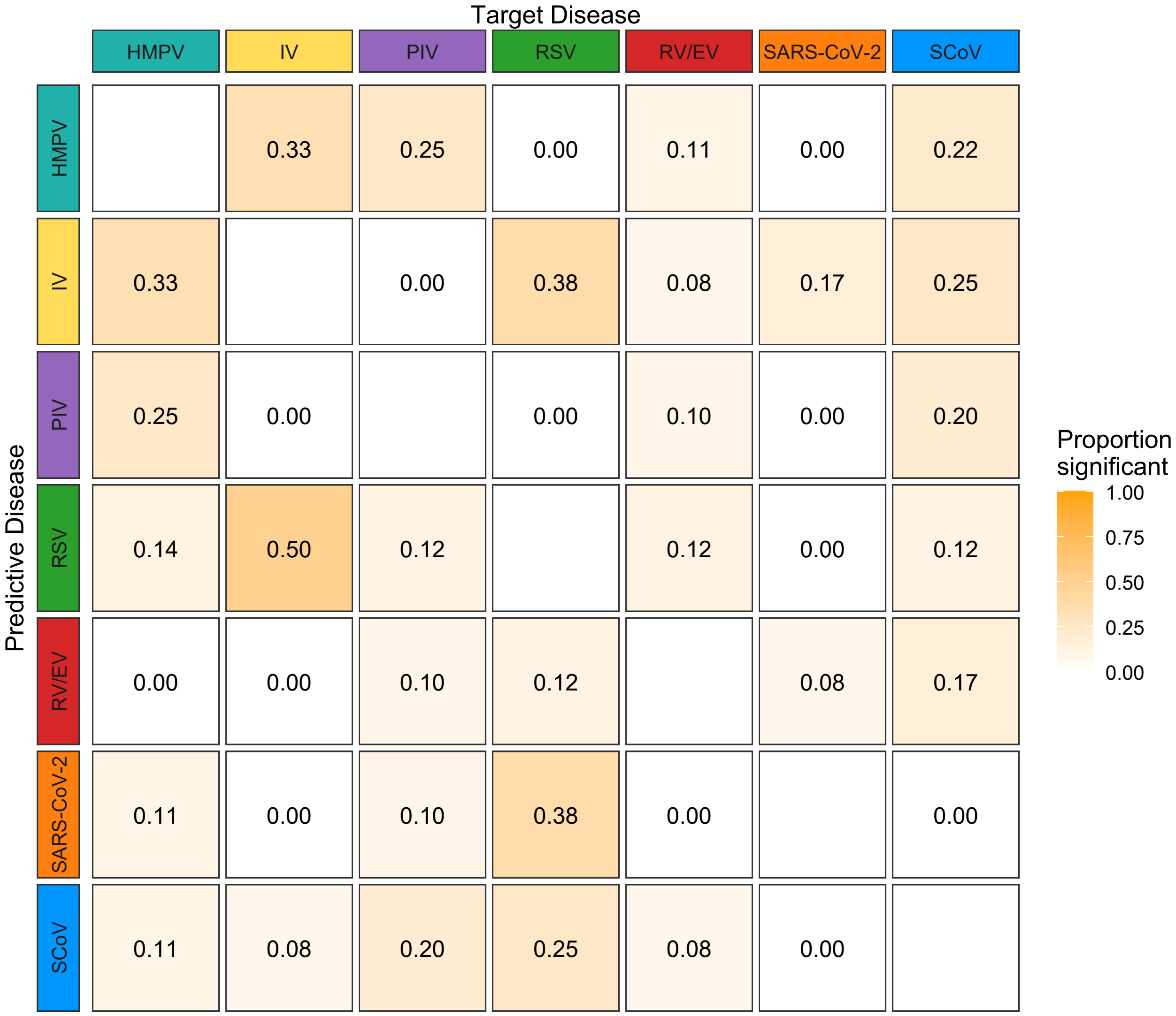


Figure B.4: Proportion of regions with significant pairwise Granger causality tests (sensitivity analysis, $p < 0.05$).

### C Principal Component Analysis

Data for each geographical region were processed independently for region-specific PCA, while for the national-level analysis, data were aggregated across all regions.

#### C.1 Preprocessing of Time Series

To ensure comparability between national and regional PCA, identical preprocessing was applied to all pathogen time series.

##### Preprocessing time series

To ensure comparability between national and regional PCA, identical preprocessing was applied to all pathogen time series.

**Aggregation level** For the national analysis, weekly case counts were aggregated across all regions to obtain one multivariate series per pathogen set. For the regional analysis, the same steps were applied separately within each region.

**Aggregation and standardisation** Weekly case counts for each of the seven respiratory pathogens were first aggregated over age groups. To remove differences in absolute incidence between regions and emphasise relative temporal variation, each pathogen series was standardised within region according to

$$z_{r,t,d} = \frac{y_{r,t,d} - \bar{y}_{r,d}}{s_{r,d}}, \tag{C.1}$$

where $y_{r,t,d}$ denotes the weekly count of pathogen $d$ in region $r$ at week $t$, $\bar{y}_{r,d}$ its regional mean, and $s_{r,d}$ its regional standard deviation. This within-region z-scoring ensured that all pathogens contributed equally to the PCA, independent of baseline level or variance.

#### C.2 Matrix structure and decomposition

For each region $r$, the standardised data were arranged in a matrix $\mathbf{Z}_r$ of dimensions $T_r \times D$, with rows representing weeks and columns representing pathogens. PCA was then applied to $\mathbf{Z}_r$ by eigendecomposition of its covariance matrix $\mathbf{S}_r = \frac{1}{T_r - 1}\mathbf{Z}_r^\top \mathbf{Z}_r$, yielding eigenvectors $\mathbf{P}_r$ (loadings) and corresponding eigenvalues $\boldsymbol{\lambda}_r$ that represent the proportion of total variance explained by each component. Component scores were obtained as $\mathbf{Z}_r\mathbf{P}_r$ and describe the temporal evolution of the PCs within each region.

Component retention and interpretation

For each regional and national PCA, components were ranked by their explained variance. The number of retained components was determined by the Kaiser criterion (eigenvalues $> 1$) and visual inspection of scree plots. Loadings were used to identify groups of pathogens contributing to common temporal patterns, while the corresponding scores traced the temporal trajectories of these components. Comparison of loading patterns across regions provided insight into the spatial consistency of pathogen co-variation structures.

#### C.3 National analysis

We performed a PCA on the standardised weekly incidence data, pooling observations across all regions. Let $\mathbf{X}$ denote the $(R \cdot T) \times D$ data matrix (here, $R \cdot T$ was the number of region–week combinations and $D$ was the number of pathogens), with variables standardised to zero mean and unit variance. The PCA decomposition was defined as in (C.2), where $\mathbf{V}$ contained the eigenvectors (loadings) of the covariance matrix $\mathbf{X}^\top\mathbf{X}$, $\mathbf{D}$ was a diagonal matrix of singular values, and $\mathbf{UD}$ gave the PC scores. The $k$-th principal component ($\text{PC}_k$) was the projection of the data onto the $k$-th loading vector.

$$\mathbf{X} = \mathbf{UDV}^\top \tag{C.2}$$

##### Component retention

The variance explained by each component is defined by (C.3), where $\lambda_k$ is the $k$-th eigenvalue of the covariance matrix $\mathbf{X}^\top \mathbf{X}/(n-1)$.

$$\text{Var explained}(k) = \frac{\lambda_k}{\sum_{j=1}^{p} \lambda_j} \tag{C.3}$$

Component retention was guided by the Kaiser criterion (eigenvalues $> 1$) and the scree "elbow" method, both of which indicated that three components should be retained.

The proportion of total variance explained by the first three PCs was relatively low. This may have been due to within-region standardisation, which removed large between-region differences in disease magnitude and left only within-region temporal fluctuations, to multiple pathogens with partially asynchronous epidemic peaks, which reduced the strength of shared variance, or to inclusion of all weeks, not just epidemic periods, which added noise relative to peak-focused datasets.

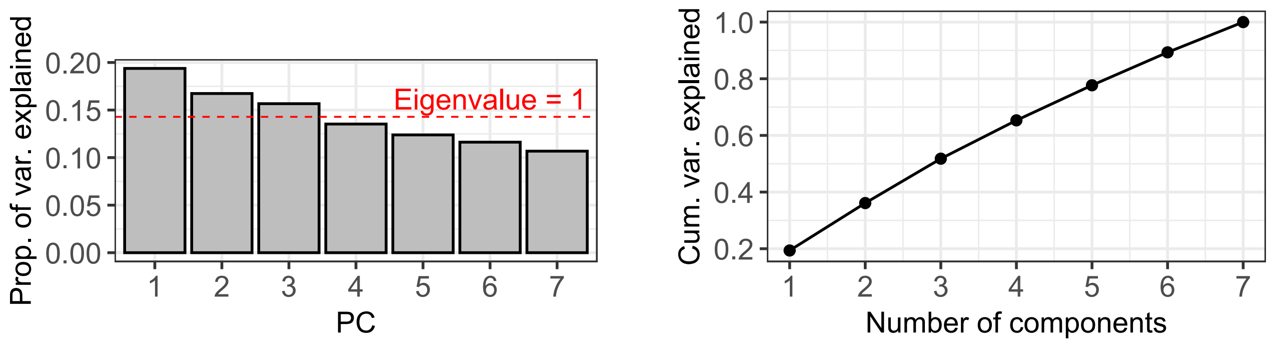


(a) (b)

Figure C.1: Variance explained in the pooled PCA analysis. The left panel shows the scree plot with the proportion of variance explained by each PC. The red dashed line indicates the Kaiser criterion (eigenvalue = 1). The right panel shows the cumulative variance explained by the ordered PCs.

##### Principal Component Scores

Scores for PC1–PC3 represented the projection of each standardised observation (region–week) into the space of the retained components. Scatterplots of the scores for each pair of components (PC1 vs. PC2, PC1 vs. PC3, PC2 vs. PC3) were shown in Figure C.2. In these plots, each point corresponded to a region–week combination, coloured by region. The distribution of points showed no clear distinct regional clusters in any PC pair. Points from different regions were well mixed throughout the space, indicating that the dominant patterns captured by PC1–PC3 were shared across regions. We also observed continuous variation along each component axis rather than discrete grouping, suggesting that temporal dynamics rather than fixed regional differences explained the main variance patterns.

To formally assess whether the first three PCs capture systematic differences between regions, we applied two complementary multivariate tests to examine whether there is a statistically significant shift in the location of the region-specific point clouds in the PC space. First, we applied Permutational Multivariate Analysis of Variance (PERMANOVA), a non-parametric test applied to a distance matrix of observations (here, Euclidean distances in the PC1–PC3 score space). It tests the null hypothesis that the centroids of the groups (regions) are the same in multivariate space. The test statistic is computed as an ANOVA-like ratio of between-group to within-group variation, and significance is assessed by permuting the group labels many times. If regions form distinct clusters in PC space, the between-group variance should be larger than expected under permutation, yielding a small $p$-value. In our case, $p = 1.0$, indicating no evidence for separation of provinces in PC1–PC3 space. Next, we applied MANOVA, a parametric test applied directly to the PC scores as multivariate response variables, with region as a categorical predictor. MANOVA tests whether the mean vector of PC scores differs across groups. It assumes multivariate normality and equality of covariance matrices, which are approximations in our

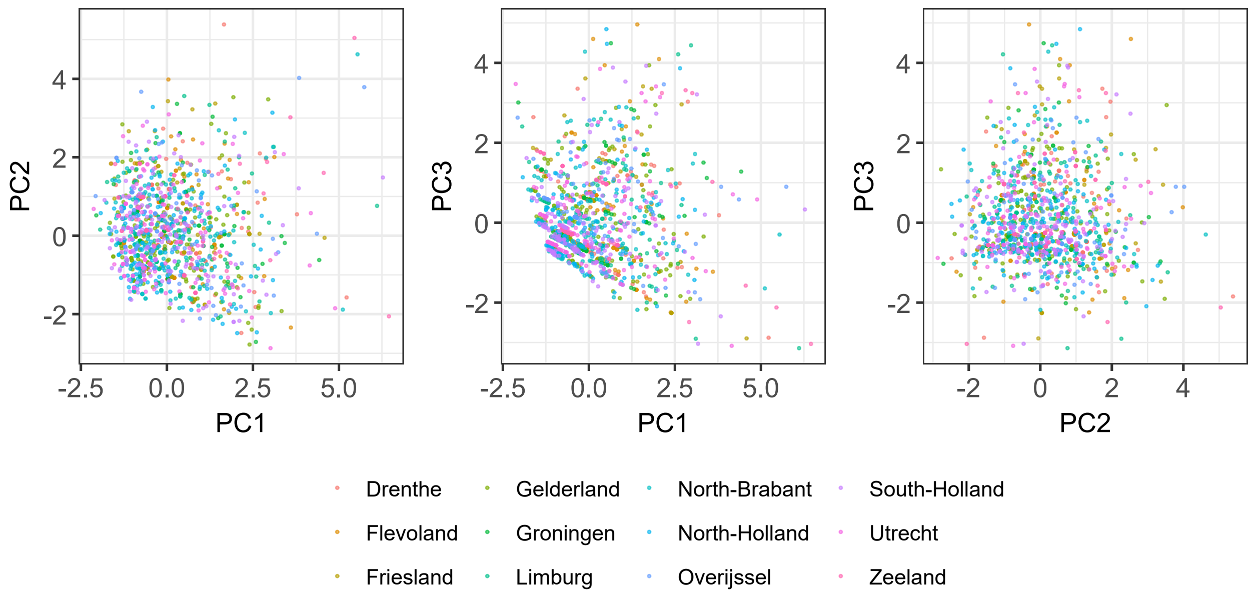


Figure C.2: Pairwise scatterplots of PC scores from the pooled analysis. Each point represents a standardised weekly observation for a given region, coloured by region. The panels show the relationships between PC1 and PC2 (left), PC1 and PC3 (middle), and PC2 and PC3 (right).

context, but still provides a useful complement to PERMANOVA. Again, $p = 1.0$ was obtained, reinforcing the conclusion from PERMANOVA.

##### Interpretation of Loadings

Loadings quantify the contribution of each disease to a principal component. Diseases with large absolute loadings exert strong influence; the sign indicates whether they vary positively or negatively with the component score. Diseases with the same sign within a PC tend to covary, whereas those with opposite signs exhibit asynchronous or opposing temporal patterns.

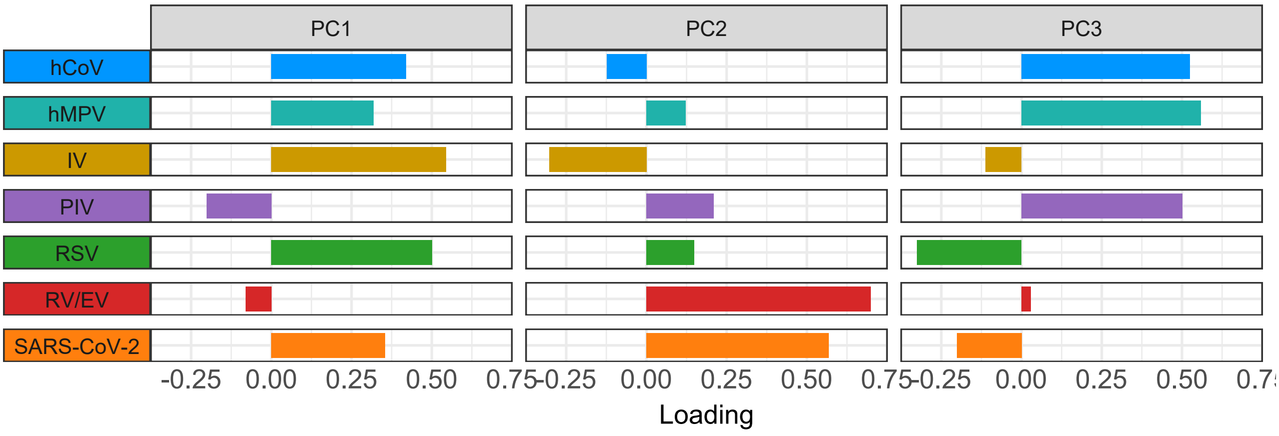


Figure C.3: Loadings of the seven respiratory pathogens on the first three PCs from the pooled PCA. Bars represent the contribution of each pathogen to the respective principal component. Positive and negative values indicate associations with the direction of the component axis, and the magnitude of each loading reflects the strength of that pathogen's influence, with higher absolute values indicating greater contribution.

To quantify similarity in loading patterns between pathogens, we computed cosine similarity as in (C.4) between their loading vectors across the first three PCs, where $\mathbf{a}$ and $\mathbf{b}$ were loading vectors of two pathogens restricted to the first $k = 3$ components. This metric takes values between $-1$ and 1, where 1 indicates identical orientation

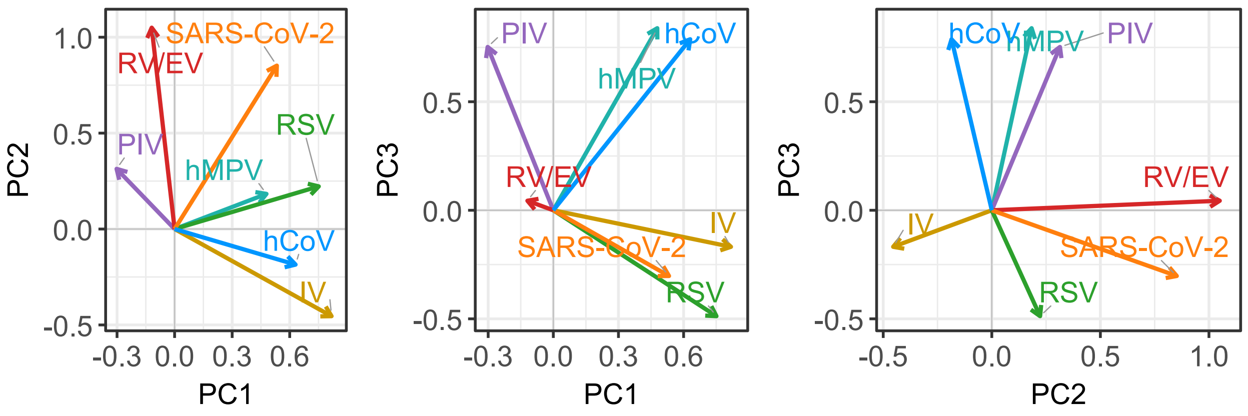


Figure C.4: PCA loading vectors for the first three PCs. Arrows represent the loadings of each respiratory pathogen on the respective PC. The direction and magnitude of each arrow indicate the strength and orientation of each pathogen's contribution in PC space.

in PC space (perfectly similar patterns of contribution), 0 indicates orthogonality (no relationship), and $-1$ indicates opposite orientations (inverse patterns of contribution). By focusing on the angle between vectors rather than their magnitude, cosine similarity highlights similarities in patterns of association regardless of absolute loading strength. Figure C.5 shows the results.

$$\cos - \text{sim}(\mathbf{a}, \mathbf{b}) = \frac{\sum_{i=1}^{k} a_i b_i}{\sqrt{\sum_{i=1}^{k} a_i^2}\,\sqrt{\sum_{i=1}^{k} b_i^2}} \tag{C.4}$$

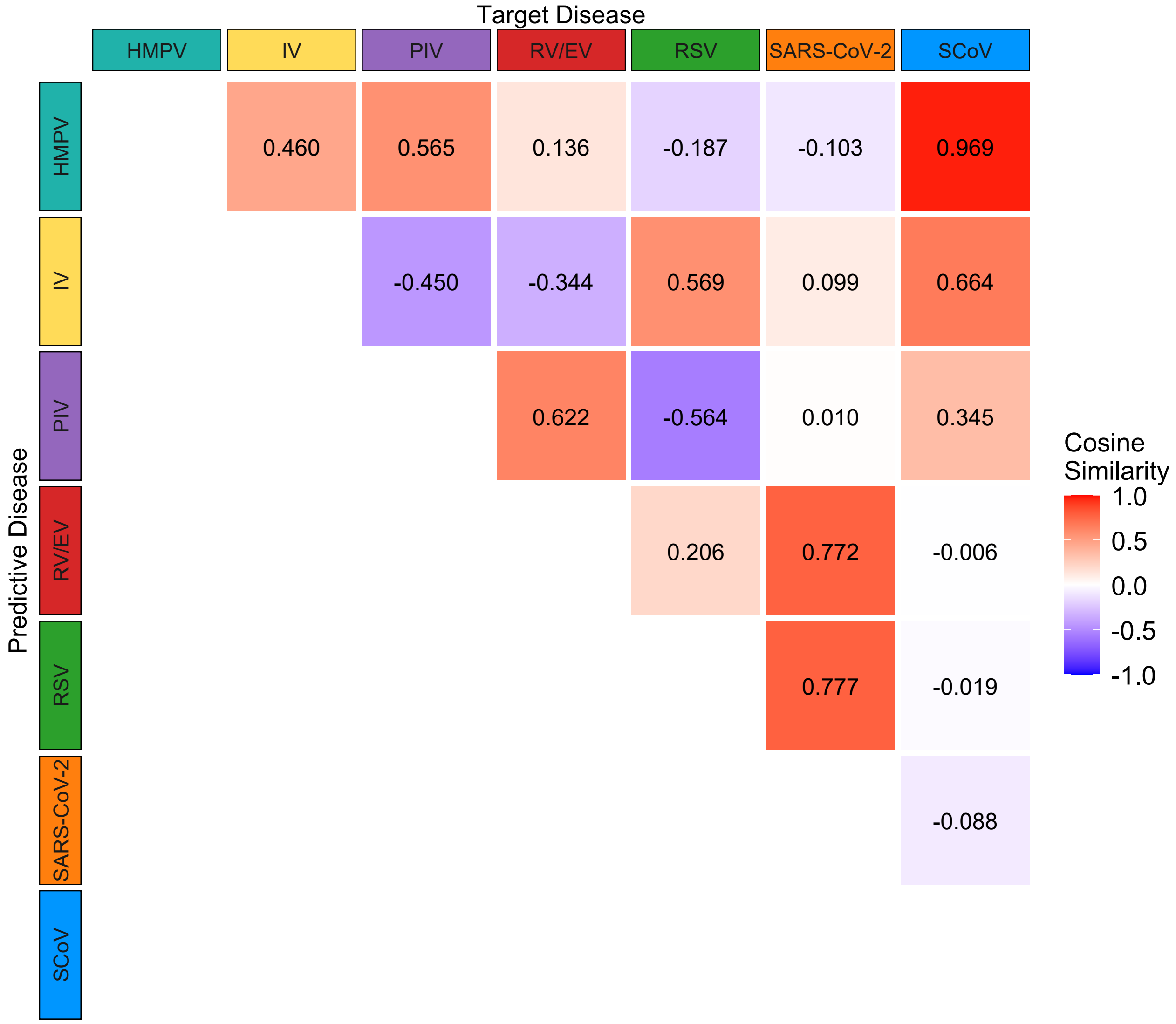


Figure C.5: Cosine similarity between pathogen loading vectors. Values show the cosine similarity between each pathogen pair across the first three principal components.

#### C.4 Region-specific analysis

To assess regional variability in pathogen contributions to overall temporal patterns, we performed separate PCA analyses for each province using weekly case counts.

##### Component retention

Across all regions, PC1 accounted for 19.7%–24.7% (mean: 22.2%) of the variance in the standardised weekly disease counts, while PC2 and PC3 contributed an additional 16.6%–19.5% (mean: 17.9%) and 14.1%–17.3% (mean: 15.7%), respectively, yielding a cumulative explained variance of 50.4%–60.3% (mean: 55.8%) for the first three components. The proportion of variance explained by the first components was similar across regions, with PC1 consistently capturing the dominant temporal co-variation between pathogens. The relatively modest cumulative variance reflected the complex, multi-dimensional nature of the data, where no small subset of components could fully describe the observed variation.

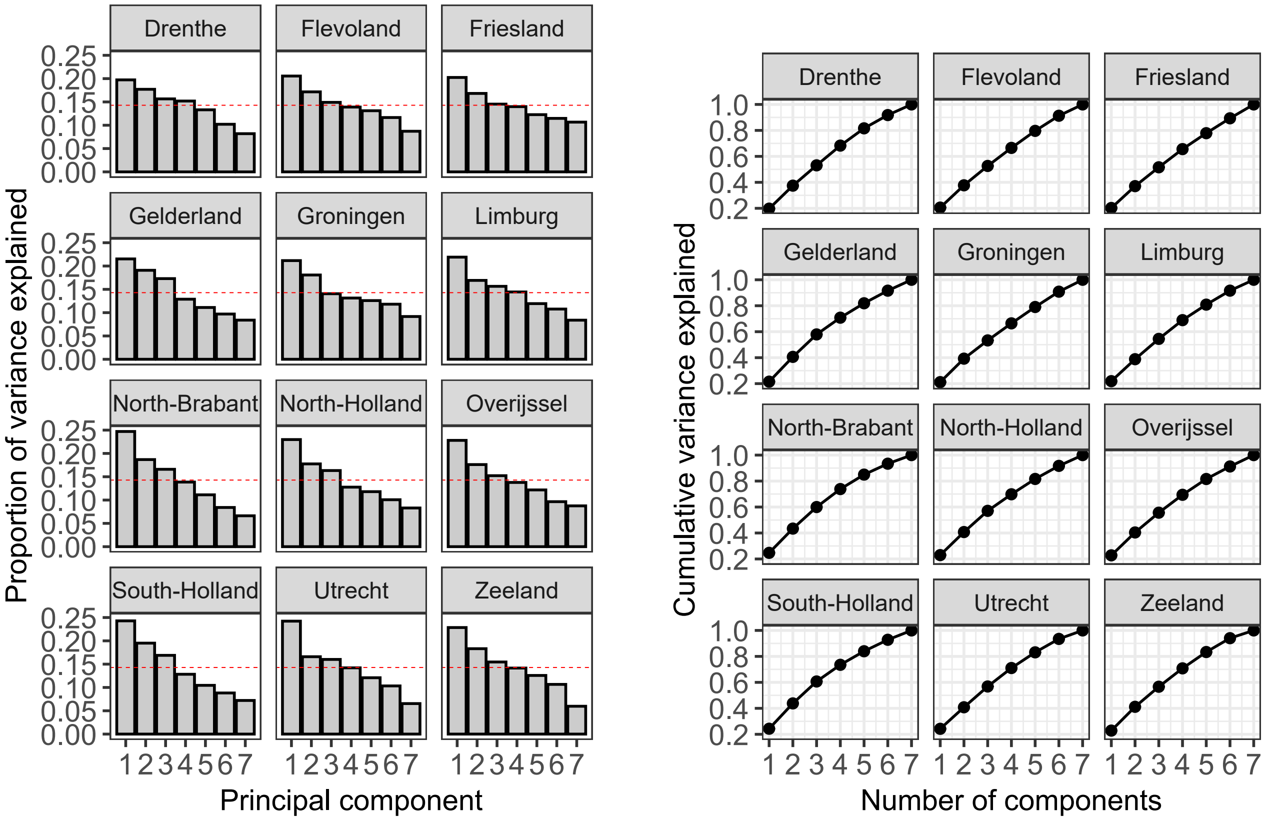


(a) (b)

Figure C.6: Variance explained in the PCA analysis by region. The left panel contains the scree plots showing the proportion of variance explained by each PC. The red dashed line indicates the Kaiser criterion (eigenvalue = 1) and the right panel contains the cumulative variances explained by the ordered PCs.

##### Interpretation of Loadings

The regional PCA revealed both consistent national-scale pathogen relationships and local deviations (Figure C.7). While pooled analysis suggested strong alignment between hCoV and hMPV (0.92) and between RSV and SARS-CoV (0.76), and opposition between IV and PIV ($-0.62$) and RSV and PIV ($-0.65$), regional analyses showed greater heterogeneity. Cosine similarity patterns (Figure C.8) highlighted stable cooperative associations such as hCoV–hMPV (mean: 0.44, 95% CI: $[0.13;, 0.75]$) and RSV–SARS-CoV-2 (mean: 0.56, 95% CI: $[0.28;, 0.84]$), and a consistent competing association of IV–PIV (mean: $-0.40$, 95% CI: $[-0.64;, -0.17]$). Wider confidence intervals for other pairs reflected substantial spatial variability, indicating that local epidemiological conditions can substantially modify inter-pathogen dynamics.

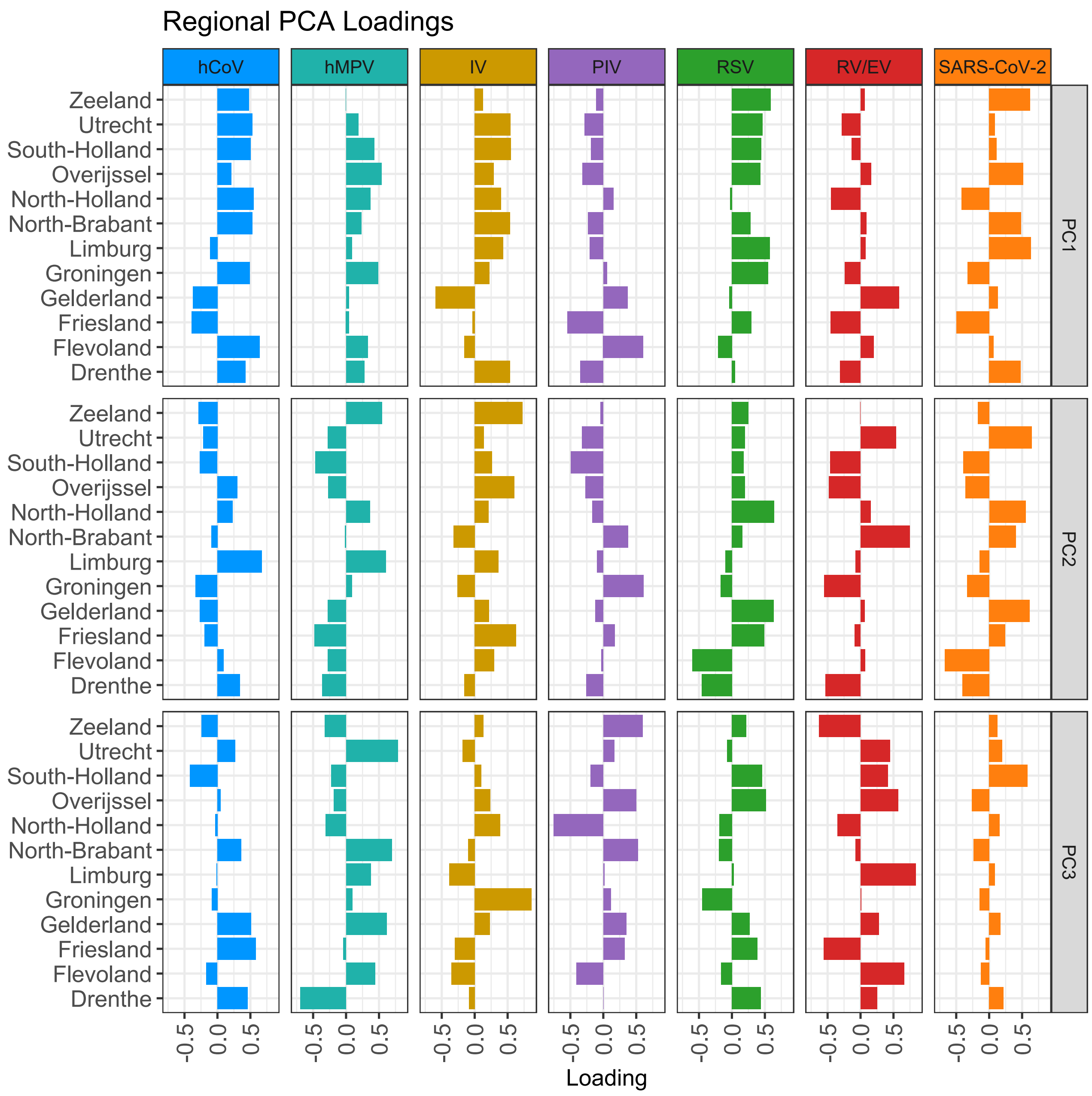


Figure C.7: Regional PCA loadings for the first three PCs (PC1–PC3). Each bar represents the loading of a pathogen for a given PC within a specific region. Positive values indicate positive associations, while negative values indicate inverse relationships.

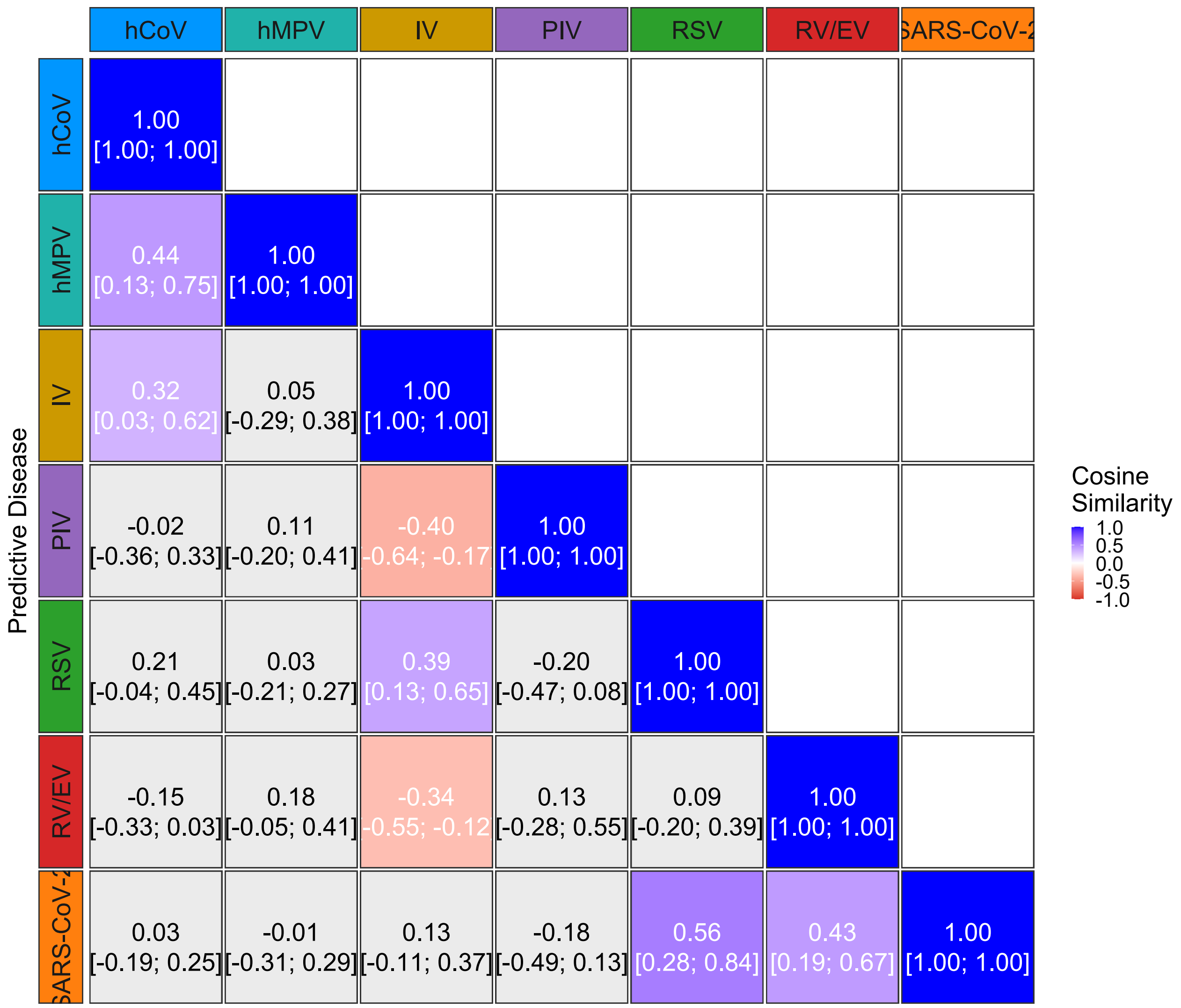


Figure C.8: Mean cosine similarity with 95% confidence intervals between loading vectors of pathogen pairs across all regions, based on the first three PCs from the regional PCA.

#### C.5 Model Specification and Inference Details

Weekly counts $Y_{d,r,a,t}$ were modelled as

$$Y_{d,r,a,t} \sim \text{NegBin}\big(\mu_{d,r,a,t},\ \kappa\big), \tag{C.5}$$

with log-mean structure

$$\log(\mu_{d,r,a,t}) = \alpha_d + \beta_{d,a} + b_{d,r} + \gamma_t + e_{t,d}. \tag{C.6}$$

The shared over-dispersion parameter $\kappa$ followed a weakly informative prior $\kappa \sim \text{Gamma}(1, 0.01)$.

**Hierarchical structure.** All latent Gaussian terms were assigned exponential penalised-complexity (PC) priors on their standard deviations:

$$\begin{aligned} \alpha_d &\sim \mathcal{N}(0, \sigma^2_{\alpha_d}), & \sigma_{\alpha_d} &\sim \text{Exp}(\lambda_{\text{pc}}), \\ b_{d,r} &\sim \mathcal{N}(0, \sigma^2_{b_d}), & \sigma_{b_d} &\sim \text{Exp}(\lambda_{\text{pc}}), \\ \beta_{d,a} &\sim \mathcal{N}\big(\mu_{\beta_{d,a}}, \sigma^2_{\beta}\big), & \sigma_{\beta} &\sim \text{Exp}\left(-\frac{\log(0.05)}{0.3}\right), \end{aligned} \tag{C.7}$$

with

$$\mu_{\beta_{d,a}} = \sum_{a'=1}^{A} C_{a,a'}\, \tilde{\beta}_{d,a'}. \tag{C.8}$$

The temporal component $\gamma_t$ was expressed as a fixed spline basis of dimension 5:

$$\gamma_t = \sum_{m=1}^{5} B_{t,m} c_m, \qquad c_m \sim \mathcal{N}(0, \sigma^2_{\gamma}), \qquad \sigma_{\gamma} \sim \text{Exp}(\lambda_{\text{pc}}), \tag{C.9}$$

where $B_{t,m}$ are pre-computed natural spline basis functions (`ns_t1-ns_t5`).

**Residual correlation.** Residuals at week $t$ were modelled as

$$\mathbf{e}_t = (e_{t,1}, \ldots, e_{t,K})^{\top} \sim \mathcal{N}_K(\mathbf{0}, \Sigma_e), \qquad \Sigma_e = L_e L_e^{\top}, \tag{C.10}$$

with $L_e = \text{diag}(\sigma_e) L_{\text{corr}}$. The marginal standard deviations $\sigma_{e,k}$ followed $\text{Exp}(\lambda_{\text{pc}})$ priors, and the correlation structure was defined by an LKJ prior:

$$L_{\text{corr}} \sim \text{LKJ\_corr\_Cholesky}(\eta = 1). \tag{C.11}$$

**Inference.** The complete latent Gaussian model was fitted using `Nimble` (R 4.4.1) with derivative support enabled for gradient-based sampling. The Cholesky factor $L_{\text{corr}}$ was updated with a Barker sampler to improve mixing; all remaining parameters used adaptive random-walk updates. Each MCMC chain comprised 80,000 iterations (40,000 burn-in, thinning = 2), yielding 20,000 posterior draws. Posterior summaries and 95% credible intervals were computed from the retained samples.

### D Short-term lagged models

#### D.1 Technical Model Details

This section provides supplementary technical details for the short-term lagged regression models introduced in the main text.

##### Distributional assumptions

All models were fitted with negative binomial likelihoods, allowing variance to exceed the mean. For a count outcome $Y_{d,r,a,t}$ with mean $\mu_{d,r,a,t}$, the variance function under each framework is:

$$\mathrm{Var}(Y_{d,r,a,t}) = \begin{cases} \mu_{d,r,a,t} + \mu^2_{d,r,a,t}/\theta, & \text{GLMM (common dispersion)} \\ \mu_{d,r,a,t} + \mu^2_{d,r,a,t}/\kappa_d, & \text{VGLM (pathogen-specific dispersion)} \\ \mu_{d,r,a,t} + \mu^2_{d,r,a,t}/\sigma_{d,r,a,t}, & \text{GAMLSS (potentially covariate-dependent dispersion)} \end{cases}$$

##### Dispersion parametrisation

In the GAMLSS, the dispersion parameter was modelled on the log scale as $\log(\kappa_d) = \delta_d$, with $\kappa_d$ pathogen-specific but constant across strata. More elaborate covariate-dependent dispersion models were considered but not supported by model selection.

#### D.2 Model Selection

##### Choice of Distribution for Count Data

To determine an appropriate distributional assumption for the weekly pathogen-specific counts, we evaluated both the Poisson and negative binomial (NB) families across all three modelling frameworks (GLMM, VGLM, GAMLSS).

We first calculated the ratio of variance to mean for each pathogen stratified by region and age group. Across all diseases this ratio was close to or greater than one, with several pathogens exhibiting clear overdispersion (Figure D.1).

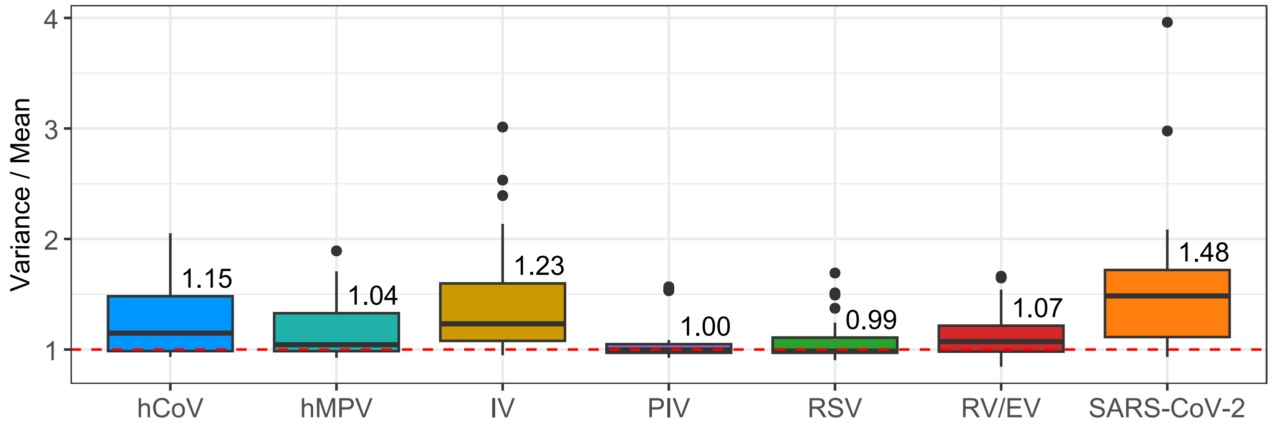


Figure D.1: Dispersion of weekly case counts per disease, stratified by age group and region. The vertical axis shows the variance-to-mean ratio, with a red dashed line at 1 indicating the expected value under a Poisson distribution. Each box summarizes the distribution across strata for the respective disease. Numbers above boxes give the median variance-to-mean ratio.

For GLMMs and GAMLSS models, the negative binomial consistently yielded lower AIC and BIC values compared to Poisson (Table D.1), indicating a superior fit. For VGLMs, the Poisson model returned slightly higher AIC values.

We further inspected model adequacy for the VGLM using randomized quantile residuals (RQRs). Both quantile–quantile plots (Figure D.2) and residual histograms (Figure D.3) favoured the negative binomial distribution.

Table D.1: Model fit comparison (Poisson vs. Negative Binomial) across frameworks.

| | Poisson | | Negative Binomial | |
|---|---|---|---|---|
| | **AIC** | **BIC** | **AIC** | **BIC** |
| **GLMM** | 24,533 | 25,159 | 24,302 | 24,936 |
| **VGLM** | 24,543 | 25,481 | 24,697 | 25,678 |
| **GAMLSS** | 24,543 | 25,781 | 24,325 | 25,570 |

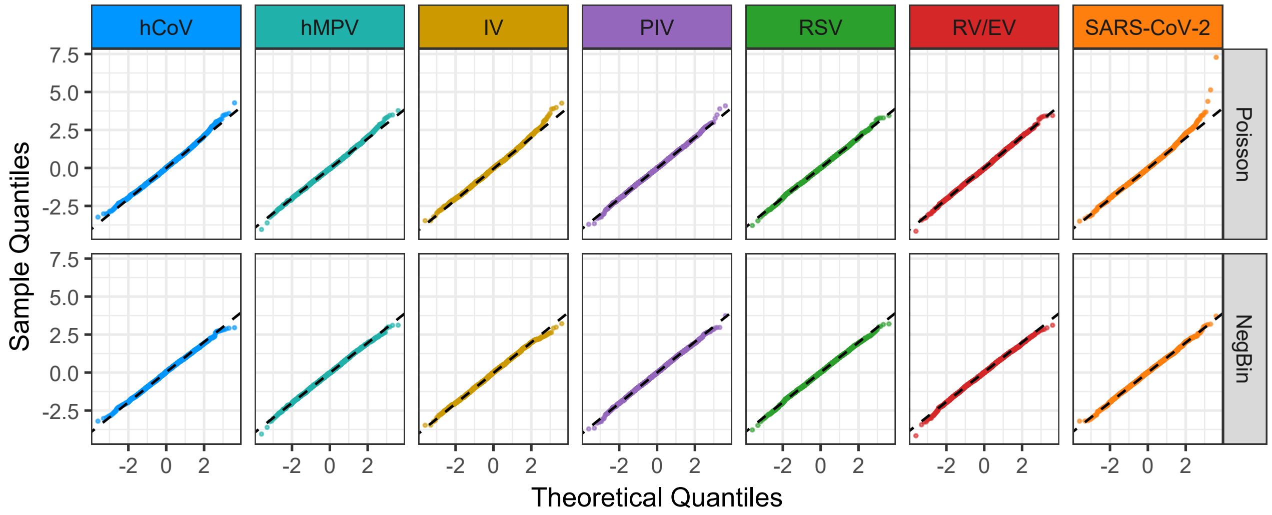


Figure D.2: Quantile–quantile plots of randomized quantile residuals for Poisson (top panel) and negative binomial (bottom panel) models, stratified by pathogen. The dashed line indicates the theoretical expectation under a standard normal distribution.

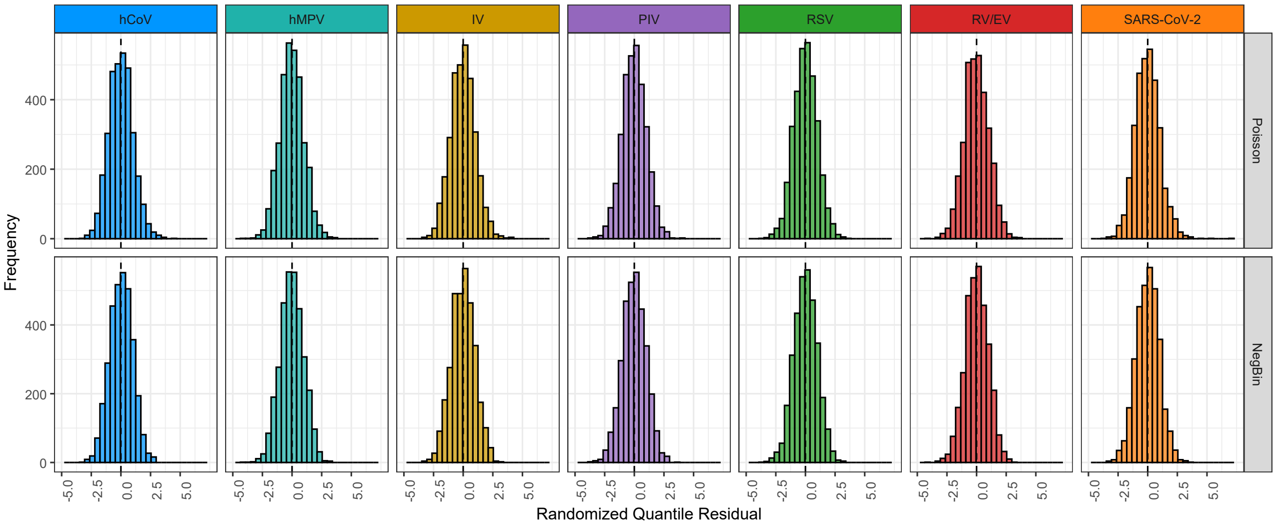


Figure D.3: Histograms of randomised quantile residuals for Poisson (top panel) and negative binomial (bottom panel) models, stratified by pathogen. The dashed vertical line marks zero.

Taken together, these results support the use of the negative binomial distribution for all model classes. Although the Poisson achieved a marginally lower AIC in the VGLM, the negative binomial provided a better theoretical match to the observed overdispersion and showed superior residual behaviour. For comparability across frameworks and robustness in inference, all subsequent analyses were therefore based on the negative binomial distribution.

##### Choice of Time and Seasonal Component

We evaluated a range of specifications for the temporal trend and seasonal component, including linear time, natural cubic splines with 2–5 degrees of freedom, sine/cosine Fourier terms, and standardized temperature covariates [47]. Model fit was assessed using AIC and BIC across frameworks (GLMM, VGLM, GAMLSS), with convergence considered as an additional criterion.

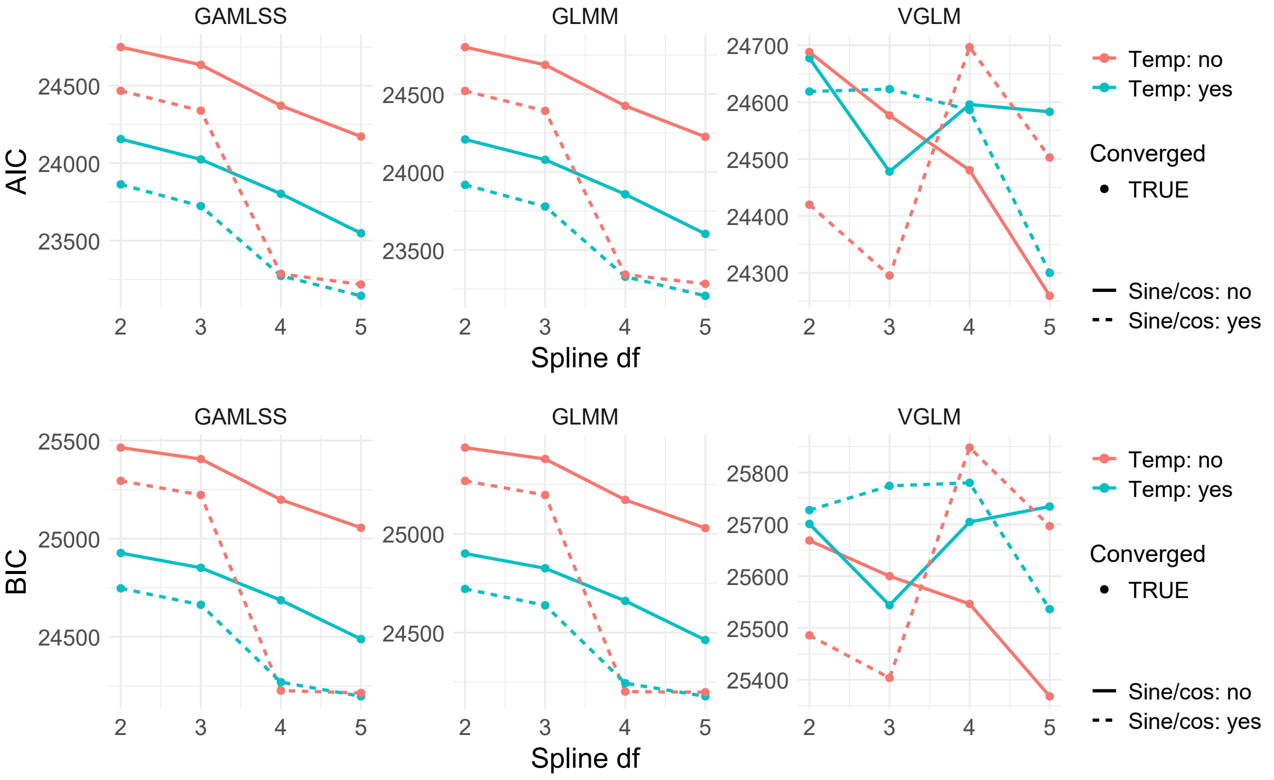


Figure D.4: Model fit (upper panel: AIC, lower panel: BIC) for temporal specifications across spline degrees, temperature inclusion, and Fourier terms, stratified by model framework.

Figures D.4 summarizes the results. For the GLMM, fit improved monotonically with spline complexity, with the lowest AIC/BIC at 4–5 df. At these df, adding Fourier terms further reduced AIC/BIC while temperature gave only small additional gains. For the GAMLSS, fit also improved up to 4–5 df. Fourier terms consistently lowered AIC/BIC, and temperature yielded a modest further reduction. For the VGLM, patterns were less regular, with the best fit at 5 df without Fourier and without temperature. Adding temperature generally worsened fit, and Fourier terms did not improve fit.

Taken together, we adopt natural cubic splines with 5 df and Fourier sine/cosine terms as the common temporal specification across frameworks, and omit temperature, as shown in (D.1), where $B_k(t)$ are the basis functions of a natural cubic spline with 5 degrees of freedom, $t$ is the standardized week index, and the sine/cosine terms capture annual seasonality. The coefficients $\beta_{k,r,d}$ and $\gamma_{1,r,d}, \gamma_{2,r,d}$ are disease- and region-specific. This matches the best configurations in GLMM and GAMLSS and remains competitive for VGLM, while keeping a single, comparable structure.

$$f_{\text{time},r,d}(t) = \sum_{k=1}^{5} \beta_{k,r,d} B_k(t) + \gamma_{1,r,d} \sin\left(\tfrac{2\pi t}{52}\right) + \gamma_{2,r,d} \cos\left(\tfrac{2\pi t}{52}\right) \tag{D.1}$$

##### Assessment of Zero Inflation

We investigated whether an explicit zero-inflation component was required. For the GLMM, a negative binomial model was fitted and the `DHARMa` zero-inflation test applied. The ratio of observed to simulated zeros was close to one (1.0024) with $p = 0.9840$, indicating no evidence of zero-inflation beyond that captured by the negative binomial distribution. For the GAMLSS, negative binomial and zero-inflated negative binomial specifications were compared using AIC and BIC.

For the VGLM, zero-inflation is only implemented for univariate responses. We therefore fitted negative binomial and zero-inflated negative binomial models separately for each pathogen and compared AIC and BIC. Summed criteria across pathogens provided an overall measure, and per-pathogen differences were also examined.

Table D.2: Model fit comparison (Negative Binomial vs. Zero–inflated Negative Binomial) across frameworks. For VGLM, results are by pathogen.

| | | **Negative Binomial** | | **Zero–inflated NB** | |
|---|---|---|---|---|---|
| | | **AIC** | **BIC** | **AIC** | **BIC** |
| GLMM | | 26,325 | 26,912 | – | – |
| GAMLSS | | 23,279 | 24,806 | 23,281 | 24,816 |
| VGLM | hMPV | 1,687 | 1,858 | 1,902 | 2,079 |
| | IV | 2,681 | 2,852 | 2,907 | 3,084 |
| | PIV | 2,227 | 2,397 | 2,536 | 2,713 |
| | RV/EV | 7,343 | 7,514 | 7,345 | 7,522 |
| | RSV | 1,532 | 1,702 | 1,962 | 2,139 |
| | SARS-CoV-2 | 4,388 | 4,559 | 4,390 | 4,567 |
| | hCoV | 4,080 | 4,250 | 4,082 | 4,258 |

Across all three frameworks, we found no evidence of excess zeros beyond what was captured by the negative binomial distribution. The inclusion of a zero-inflation component did not improve model fit and in several cases led to substantial deterioration. Although a zero-inflation component was initially considered, systematic tests across frameworks showed no improvement in fit. We therefore retained the standard negative binomial distribution without a zero-inflation term in all subsequent analyses.

##### Variance Component in GAMLSS

In addition to the mean structure, the GAMLSS framework allows for covariate effects on the dispersion parameter $\kappa_d$ of the negative binomial distribution. We compared several candidate specifications for $\log(\kappa_d)$, including (i) a constant dispersion common to all pathogens, (ii) pathogen-specific dispersion, and (iii) extensions with age, region, temperature, time, or Fourier terms interacted with disease. Model selection was based on AIC and BIC, with convergence verified in all cases.

Table D.3: Model fit comparison for alternative variance specifications in the GAMLSS framework. The baseline model is pathogen-specific dispersion $\log(\kappa_d) \sim 0 + \text{disease}$.

| **Variance specification** | **AIC** | **BIC** |
|---|---|---|
| $\sim 1$ | 23,279 | 24,806 |
| $\sim 0 + \text{disease}$ | 23,261 | 24,836 |
| $\sim 0 + \text{disease} + [\cos(2\pi t/52) + \sin(2\pi t/52)] : \text{disease}$ | 29,981 | 31,661 |
| $\sim 0 + \text{disease} + \text{temp} : \text{disease}$ | 23,258 | 24,890 |
| $\sim 0 + \text{disease} + t : \text{disease}$ | 23,269 | 24,900 |
| $\sim 0 + \text{disease} + \text{age group} : \text{disease}$ | 23,272 | 24,904 |
| $\sim 0 + \text{disease} + \text{region} : \text{disease}$ | 107,413 | 109,607 |

Results showed that AIC slightly favoured pathogen-specific dispersion ($\log(\kappa_d) \sim 0 + \text{disease}$, AIC $=$ 23,261 vs. 23,279). In contrast, BIC clearly favoured the simplest specification with a common dispersion parameter ($\sim 1$, BIC $=$ 24,806 vs. 24,836). Adding further covariates increased both AIC and BIC. We therefore retained the pathogen-specific specification. This choice aligns with the VGLM framework, where dispersion is also estimated separately for each pathogen, and acknowledges inherent variability differences between pathogens. Although BIC penalised the added complexity, the AIC preference and biological plausibility justified the pathogen-specific model.

#### D.3 Model diagnostics

##### GLMM

The adequacy of the GLMM was evaluated using simulation-based residual diagnostics from the `DHARMa` package.

Across pathogens, residuals were approximately uniformly distributed (Figures D.5 and D.6), and QQ-plots aligned closely with theoretical uniform quantiles, indicating broadly adequate calibration of the fitted negative binomial distribution.

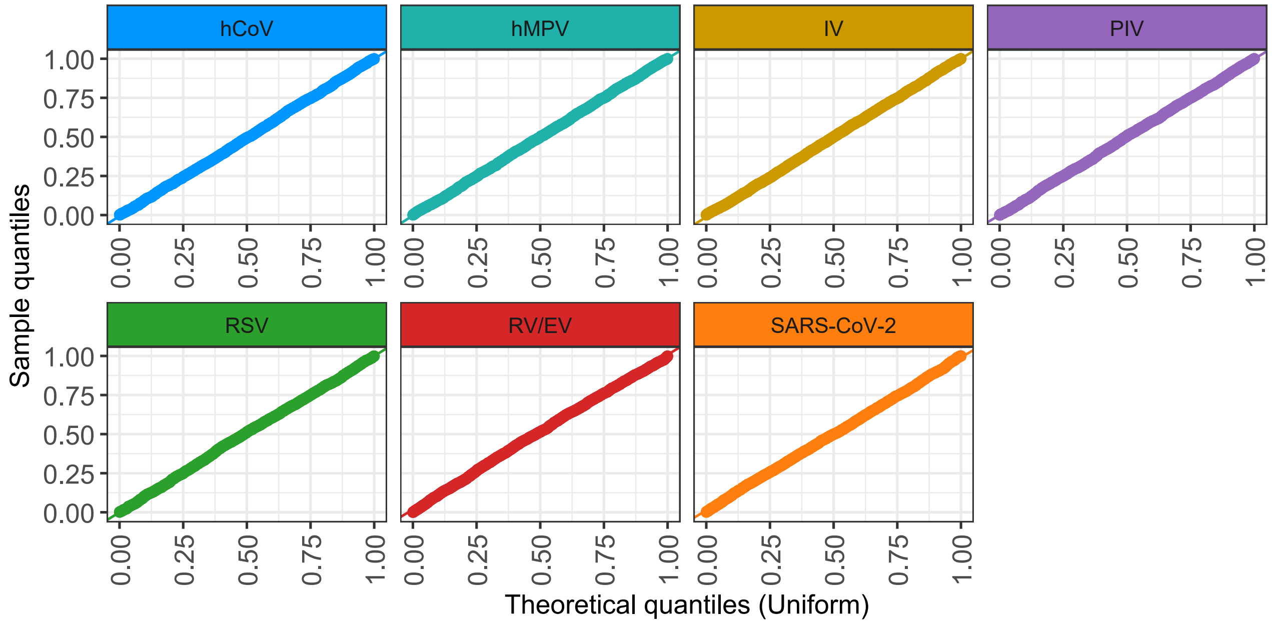


Figure D.5: Disease-specific QQ-plots of scaled residuals against the Uniform(0,1) distribution for the GLMM.

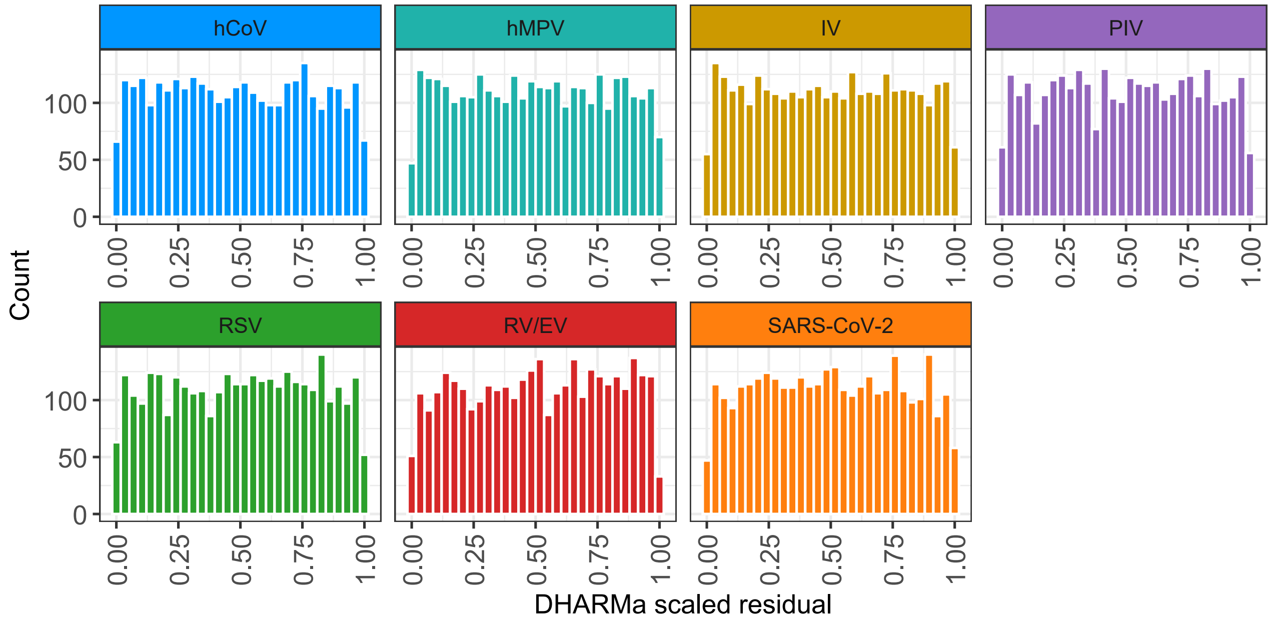


Figure D.6: Disease-specific histograms of scaled residuals for the GLMM.

Disease-specific residual-versus-fitted plots (Figure D.7) revealed some curvature: residuals increased at higher fitted values for hMPV and RV/EV, and decreased slightly for IV and SARS-CoV-2. These patterns indicate minor remaining mean–variance misfit.

Formal DHARMa tests indicated no evidence of overdispersion (dispersion test $p = 0.982$) or zero inflation (zero-inflation test $p = 0.914$). Taken together, these diagnostics, including pooled residual plots (Figure D.8),

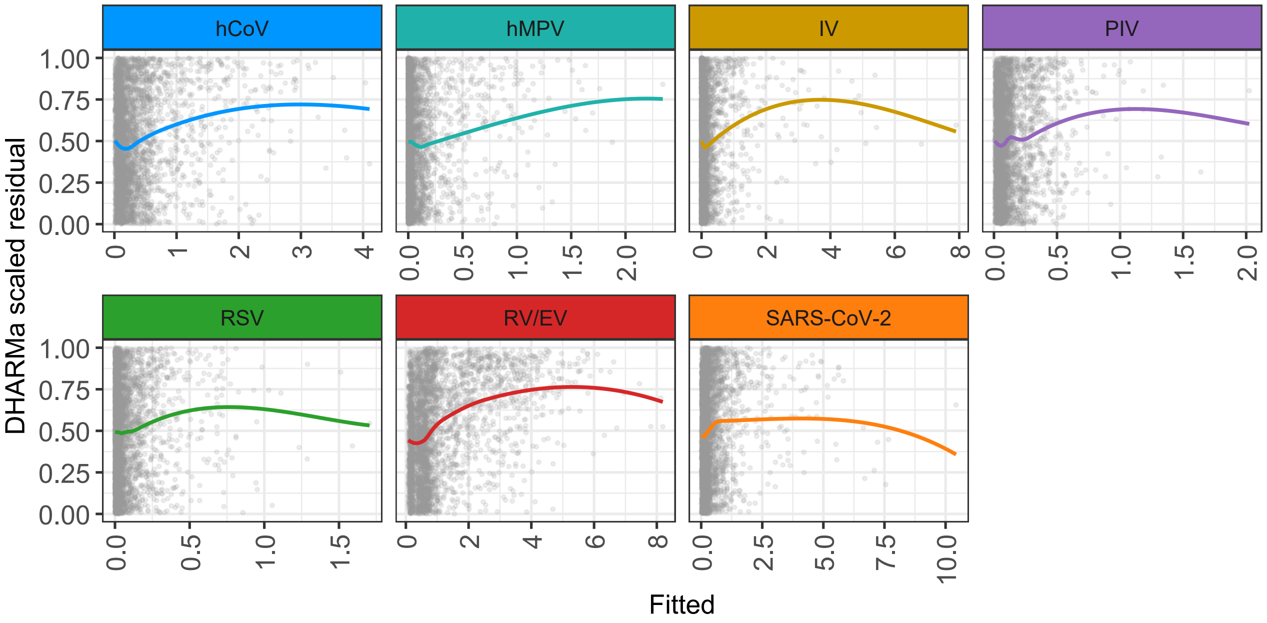


Figure D.7: Disease-specific scaled residuals versus fitted values for the GLMM. Each panel shows DHARMa scaled residuals against fitted values with loess smooth (coloured by pathogen).

support the adequacy of the negative binomial specification with random region intercepts and justify its use for inference on cross-pathogen lag effects.

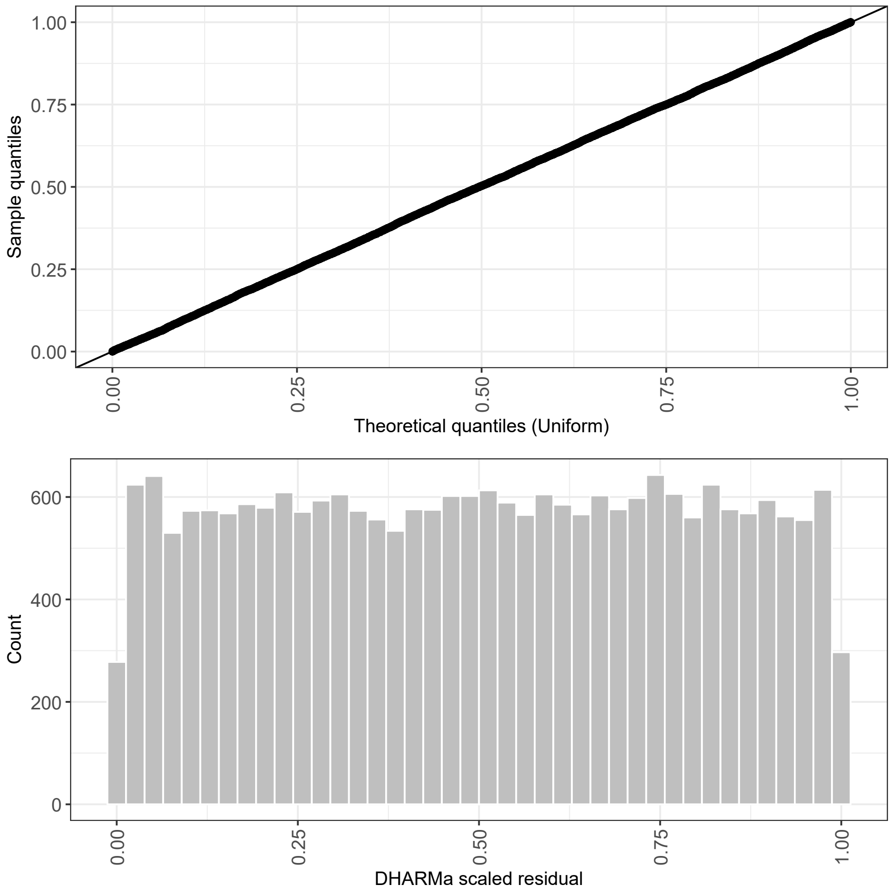


Figure D.8: Global QQ-plot and histogram of scaled residuals across all pathogens for the GLMM.

#### VGLM

The VGLM with negative binomial responses and pathogen-specific dispersion was also evaluated using randomized quantile residuals. QQ-plots of randomized quantile residuals (Figure D.9) showed close alignment with the theoretical normal distribution across all pathogens, with only mild deviations in the extremes. The corresponding histograms (Figure D.10) were centered near zero and exhibited approximately Gaussian shapes, with minor pathogen-specific differences in spread. Together, these diagnostics indicate that the negative binomial VGLM provided an adequate distributional fit, though small tail departures remain.

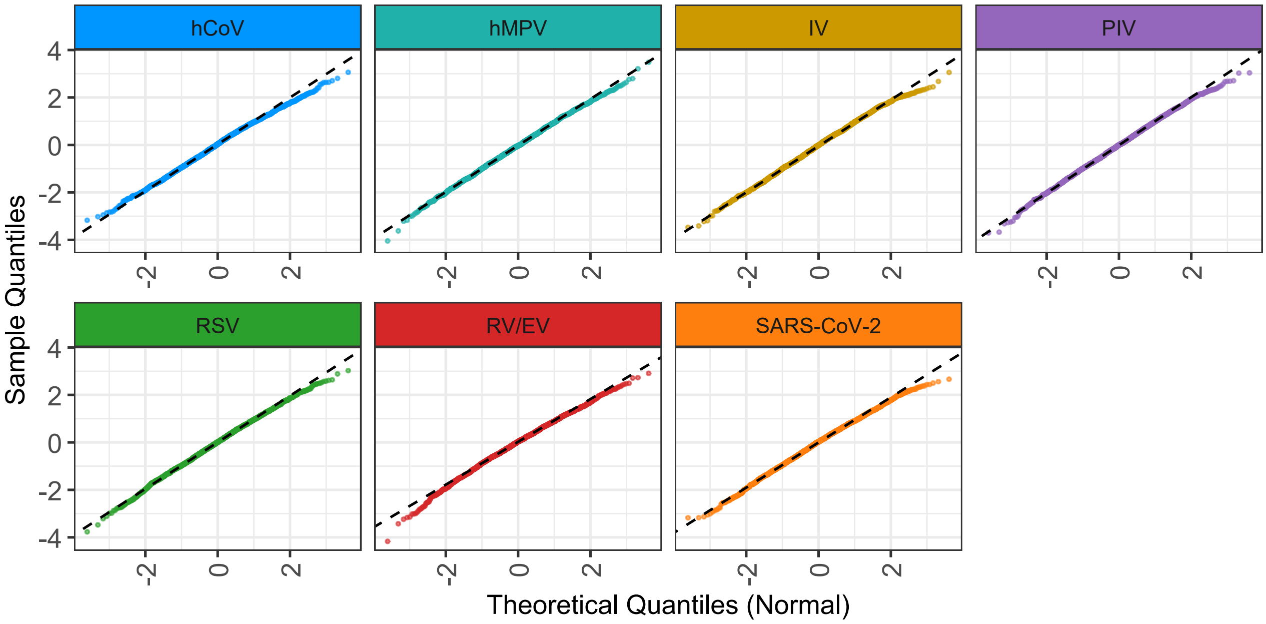


Figure D.9: Disease-specific QQ-plots of randomized quantile residuals for the VGLM with negative binomial responses.

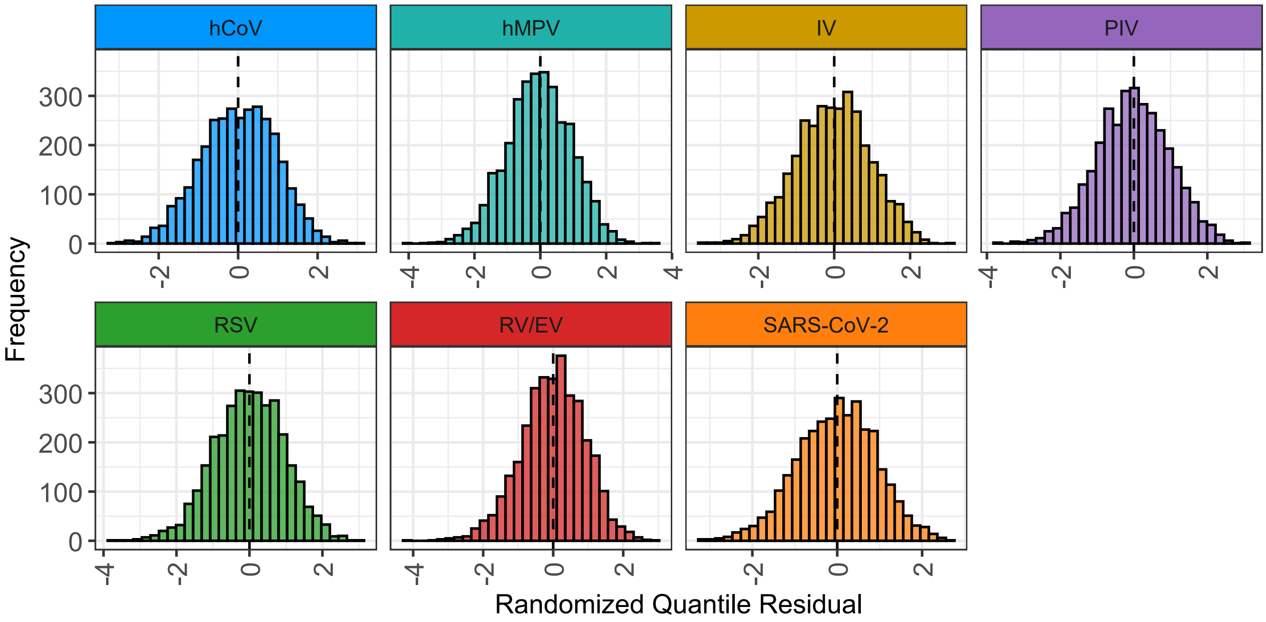


Figure D.10: Disease-specific histograms of randomized quantile residuals for the VGLM.

Residuals versus fitted means (Figure D.11) were broadly well-centered. Variance–mean plots (Figure D.12) followed the negative-binomial expectation closely, with only mild excess variance at higher fitted means. Dispersion patterns varied between pathogens, but no strong model misspecification was evident.

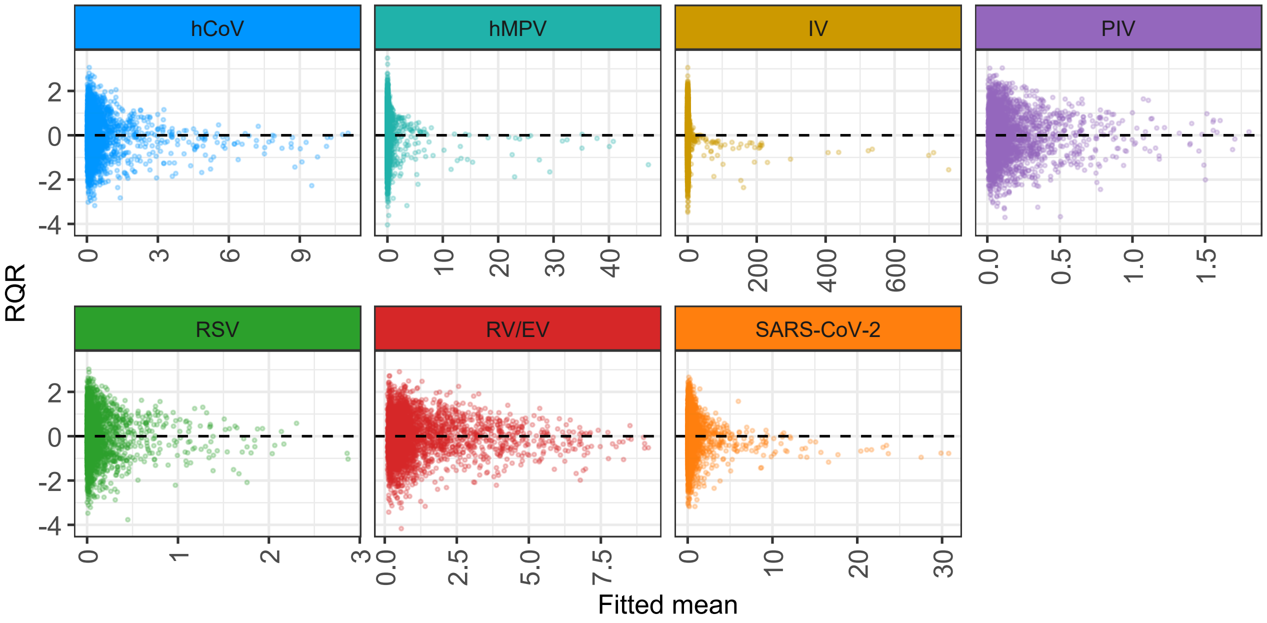


Figure D.11: Randomized quantile residuals versus fitted means by pathogen for the VGLM.

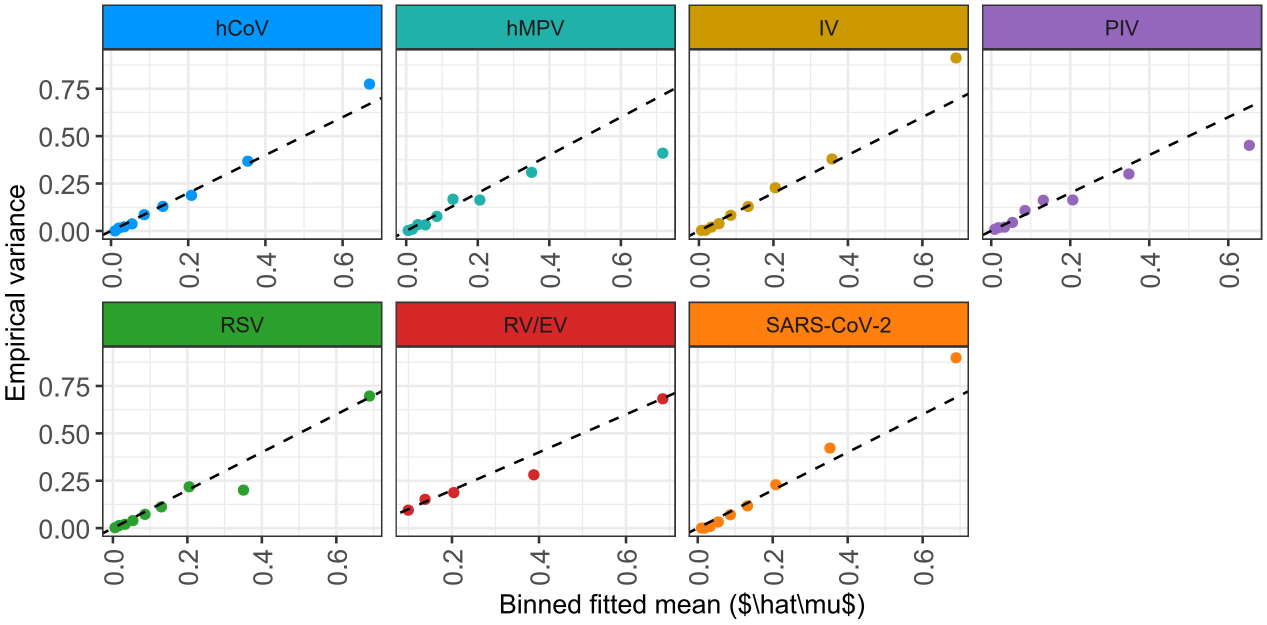


Figure D.12: Binned fitted mean ($\hat{\mu}$) versus empirical variance by pathogen for the VGLM. The dashed line indicates the Poisson mean–variance identity.

Autocorrelation functions of residuals (Figure D.13) showed minimal temporal dependence beyond lag 1. No substantial persistence was evident, indicating that spline and seasonal terms adequately absorbed epidemic structure.

##### GAMLSS

For the GAMLSS, Q–Q plots (Figure D.14) and histograms (Figure D.15) indicated approximate normality of randomized quantile residuals, with moderate tail deviations most visible for RV/EV and SARS-CoV-2.

Residuals versus fitted means (Figure D.16) showed little systematic curvature.

Variance–mean comparisons (Figure D.17) matched the negative-binomial expectation closely, confirming adequacy of pathogen-specific dispersion.

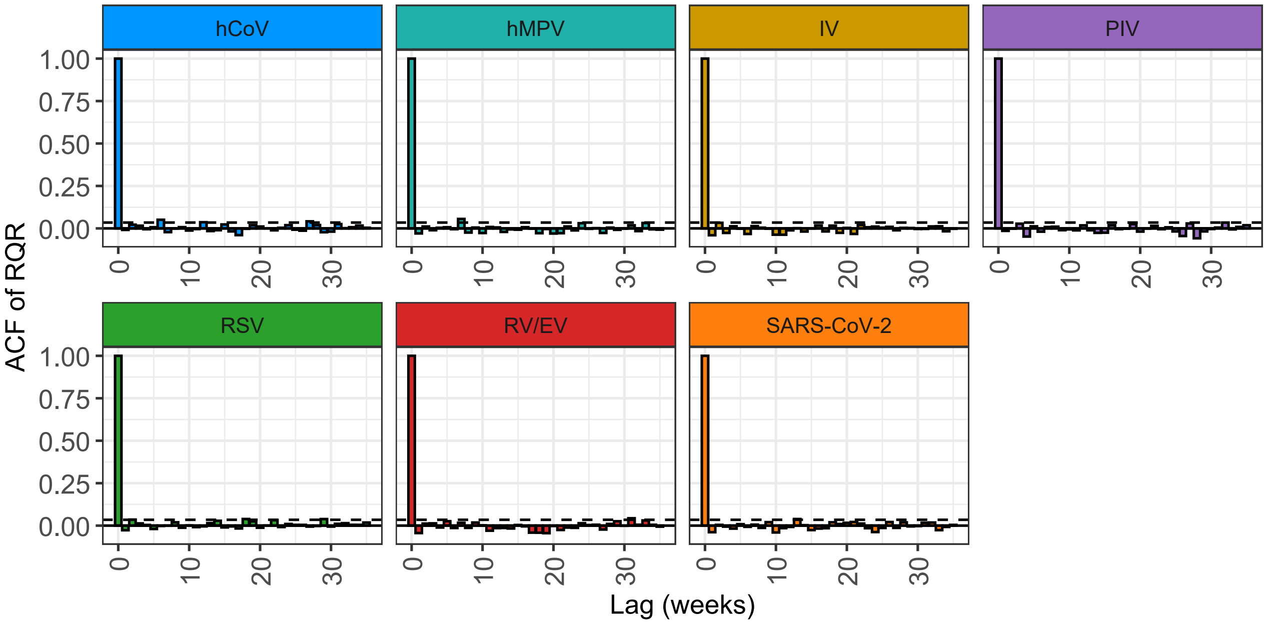


Figure D.13: Autocorrelation functions of randomized quantile residuals for the VGLM. Dashed lines mark approximate $\pm 1.96/\sqrt{n}$ bounds.

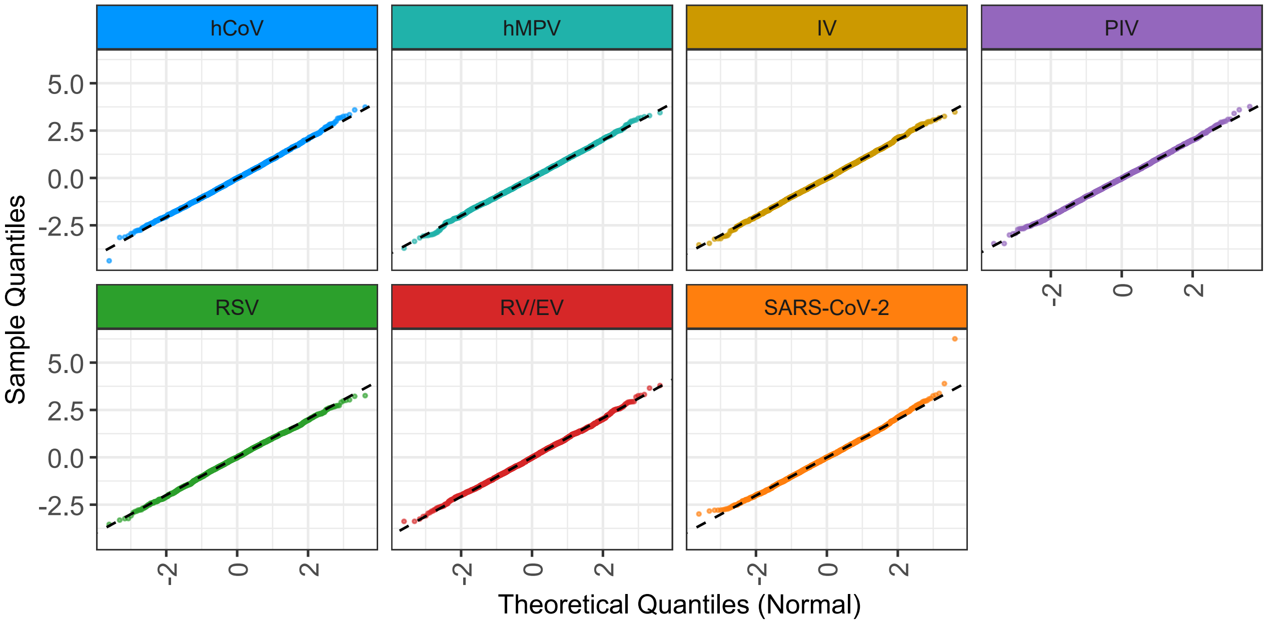


Figure D.14: Disease-specific QQ-plots of randomized quantile residuals for the GAMLSS.

Autocorrelation functions (Figure D.18) showed little residual temporal dependence beyond lag 1. Overall, GAMLSS achieved good variance alignment across pathogens, with only mild tail departures and limited residual correlation.

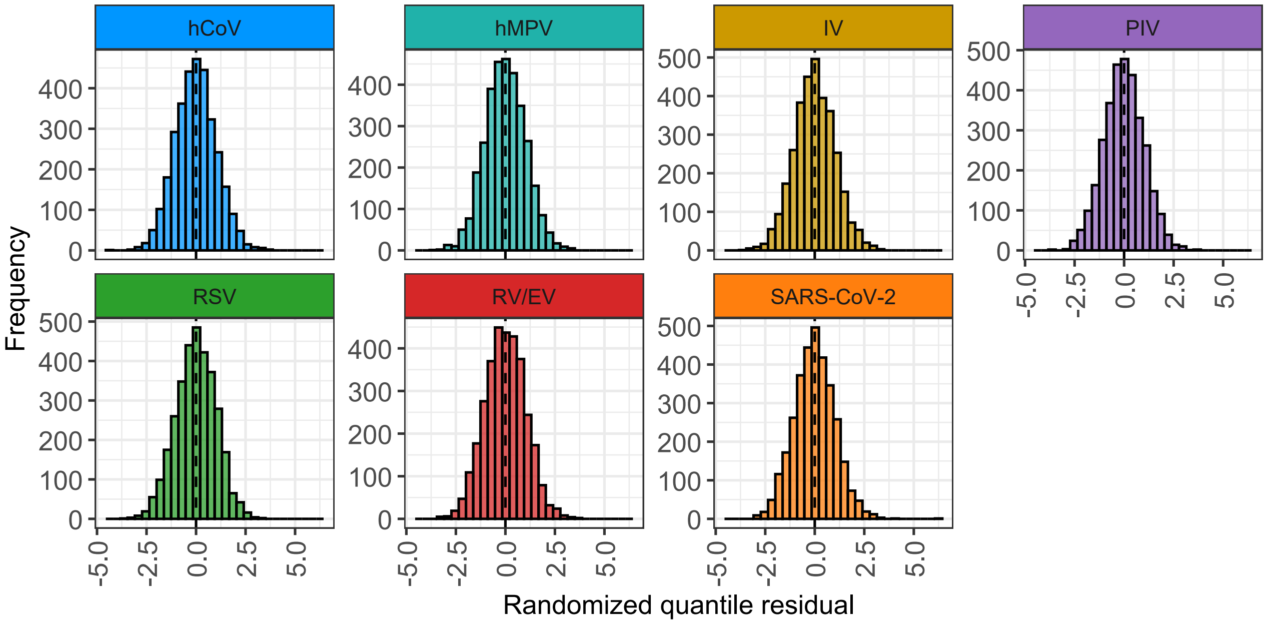


Figure D.15: Histograms of randomized quantile residuals for the GAMLSS.

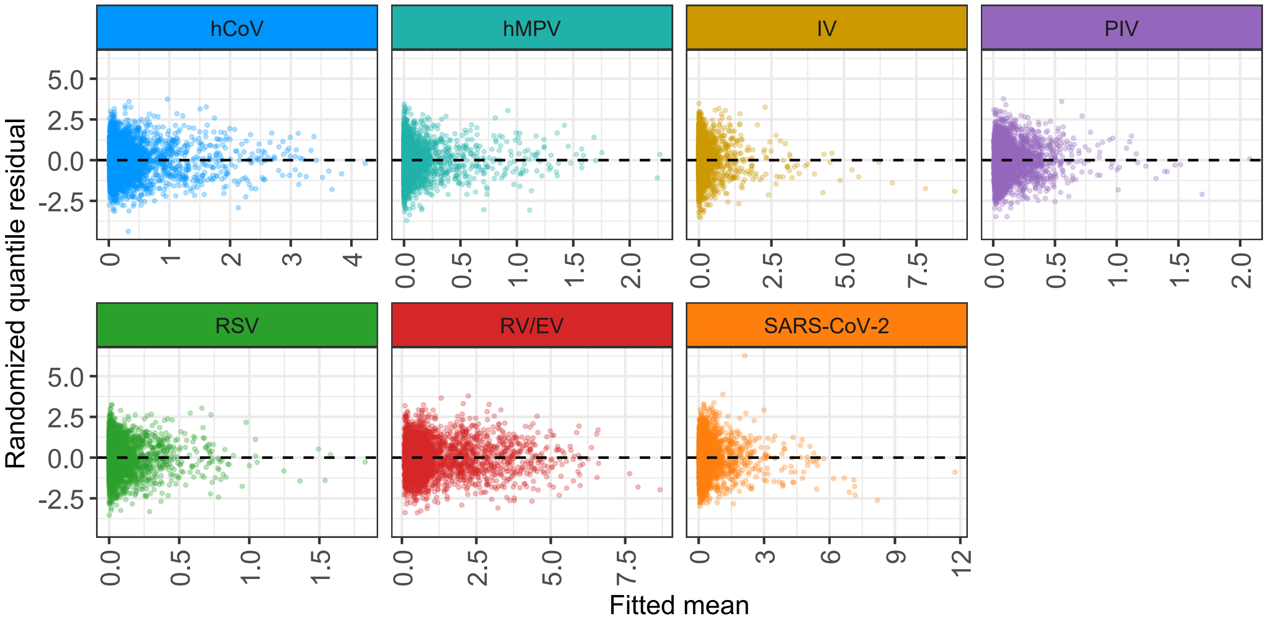


Figure D.16: Randomized quantile residuals versus fitted means by pathogen for the GAMLSS.

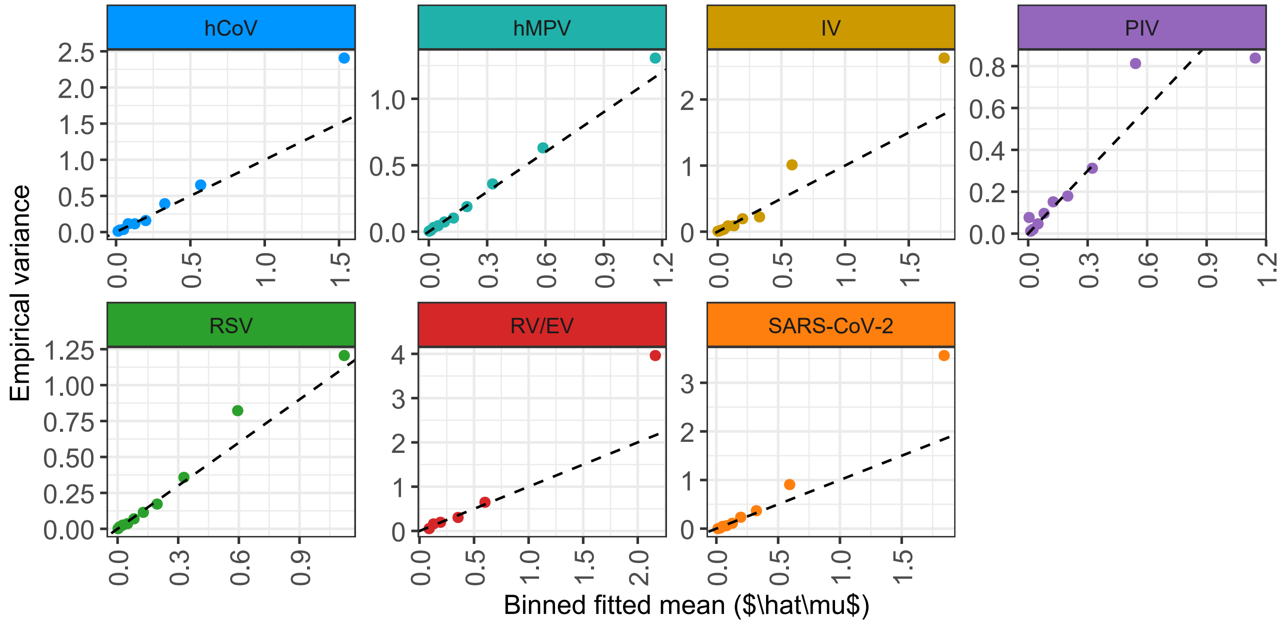


Figure D.17: Binned fitted mean ($\hat{\mu}$) versus empirical variance by pathogen under the GAMLSS. The dashed line indicates the Poisson variance identity.

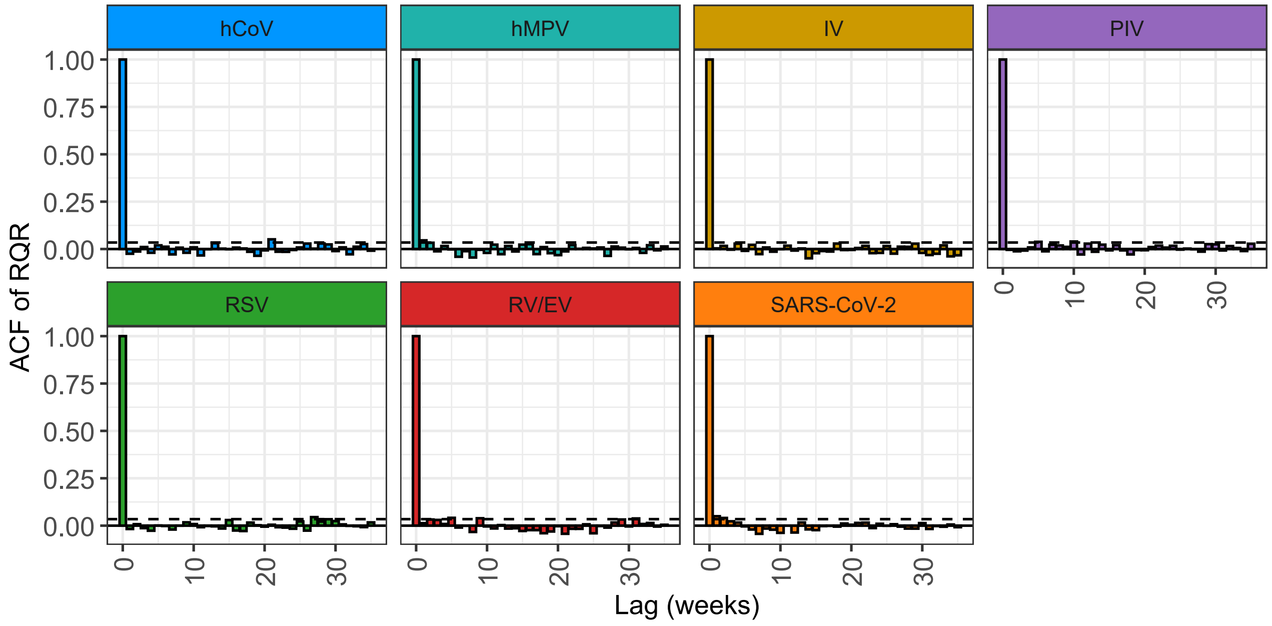


Figure D.18: Autocorrelation functions of randomized quantile residuals by pathogen for the GAMLSS. Dashed lines show approximate $\pm 1.96/\sqrt{n}$ bounds.

##### D.4 Additional Results

Across the three regression frameworks (GLMM, VGLM, GAMLSS), model-specific features captured different aspects of between-region and between-pathogen variability, excluding lagged effects which are reported separately.

The GLMM identified clear spatial heterogeneity. The regional random intercept had variance $\widehat{\text{Var}}(b_r) = 0.46$ (SD = 0.68) across 12 regions, corresponding to roughly two-fold baseline differences in incidence on the log scale. The negative binomial dispersion parameter was $\hat{\theta} = 6.47$ (NB2), consistent with moderate overdispersion since $\text{Var}(Y) \approx \mu + \mu^2/\theta$. Age showed a consistent negative contrast for the older group. Broad temporal variation was absorbed by seasonal spline and Fourier terms, which were not interpreted individually.

In the VGLM, dispersion was pathogen-specific ($\kappa_d$) but estimated jointly. The model achieved a log-likelihood of $-12055.36$. Hauck–Donner warnings appeared for several intercept and spline terms, reflecting separation or extreme leverage in nuisance components, but these did not affect the substantive cross-pathogen findings (see Diagnostics).

In the GAMLSS, dispersion was pathogen-specific but constant within each pathogen. Under the NB1 parametrisation, larger $\sigma_d$ implied stronger overdispersion. Dispersion was highest for SARS-CoV-2 ($\sigma \approx 0.32$) and influenza ($\sigma \approx 0.26$), intermediate for hCoV and PIV ($\sigma \approx 0.23$–$0.25$), and lowest for hMPV and RV/EV ($\sigma \approx 0.05$–$0.07$). RSV showed moderate overdispersion ($\sigma \approx 0.10$). These estimates indicate clear between-pathogen differences in variability beyond mean incidence.

#### E Sensitivity Analysis - Effect of Season

We conducted a sensitivity analysis in which epidemic seasons were defined categorically, with September–April periods numbered sequentially and off-season weeks coded as 0. This allowed baseline incidence to vary across successive epidemic periods while controlling for weeks outside the respiratory season. Figures E.1–E.3 show the resulting cross-pathogen estimates for the GLMM, VGLM, and GAMLSS frameworks, directly compared to the corresponding baseline models.

The overall picture was consistent across specifications. Associations that appeared in the baseline models generally persisted under the categorical season coding, with effect sizes and directions closely aligned. Likewise, the sets of non-significant pairs were stable, indicating that adjustment for season did not introduce spurious associations. Crucially, no associations reversed sign under this specification. The categorical-season models therefore support the robustness of our main findings: cross-pathogen interactions identified in the baseline regressions are not artefacts of seasonal adjustment, but remain detectable when an alternative representation of seasonality is applied.

To further explore temporal heterogeneity, we additionally estimated cross-pathogen effects separately for each season using the VGLM framework. This analysis showed that many associations were reproduced across multiple seasons, while others appeared only in individual epidemic periods. Off-season estimates were generally weak or absent, reflecting minimal circulation outside the September–April window. These findings highlight that while the direction of associations is stable, their detectability can vary depending on the degree of overlap between seasonal epidemics, underlining the importance of pooling across seasons to obtain stable interaction estimates.seasonality is applied.

Table D.4: Summary of the incidence rate ratios (IRRs) for the GLMM, GAMLSS, VGLM and E&E models across all pathogen pairs with corresponding 95% confidence intervals.

| | | **Target** | | | | | | |
|---|---|---|---|---|---|---|---|---|
| **Predictor** | **Model** | **hMPV** | **IV** | **PIV** | **RSV** | **RV/EV** | **SARS-CoV-2** | **hCoV** |
| **hMPV** | **GLMM** | 1.0907<br>[0.9449; 1.2590] | 0.9633<br>[0.8335; 1.1134] | **1.3259**<br>[1.1272; 1.5597] | 1.1466<br>[0.9238; 1.4231] | 1.0622<br>[0.9828; 1.1480] | 0.9086<br>[0.8063; 1.0238] | 1.0847<br>[0.9861; 1.1931] |
| | **GAMLSS** | 1.0577<br>[0.9214; 1.2142] | 0.9529<br>[0.8177; 1.1104] | **1.2998**<br>[1.1001; 1.5358] | 1.1159<br>[0.8994; 1.3844] | 1.0552<br>[0.9836; 1.1319] | 0.9075<br>[0.7975; 1.0327] | 1.0623<br>[0.9584; 1.1775] |
| | **VGLM** | **4.5245**<br>[3.4687; 5.9016] | 0.9791<br>[0.7362; 1.3020] | **1.6629**<br>[1.3634; 2.0282] | **1.5228**<br>[1.1714; 1.9795] | 1.0927<br>[0.9926; 1.2028] | 0.9927<br>[0.8290; 1.1888] | **1.4877**<br>[1.2850; 1.7225] |
| **IV** | **GLMM** | 0.9896<br>[0.8780; 1.1155] | **1.1219**<br>[1.0493; 1.1995] | 0.8419<br>[0.7084; 1.0005] | 0.8894<br>[0.7808; 1.0131] | **0.8831**<br>[0.8333; 0.9358] | 1.0316<br>[0.9522; 1.1175] | 1.0605<br>[0.9950; 1.1303] |
| | **GAMLSS** | 0.9851<br>[0.8744; 1.1099] | **1.1281**<br>[1.0493; 1.2127] | **0.8189**<br>[0.6861; 0.9775] | 0.8967<br>[0.7855; 1.0235] | **0.8784**<br>[0.8320; 0.9274] | 1.0316<br>[0.9476; 1.1230] | 1.0500<br>[0.9816; 1.1231] |
| | **VGLM** | 1.1151<br>[0.9199; 1.3516] | **5.1971**<br>[4.4546; 6.0634] | **0.8049**<br>[0.6639; 0.9758] | 1.0985<br>[0.9414; 1.2818] | **0.8790**<br>[0.8204; 0.9418] | 1.0621<br>[0.9497; 1.1879] | **1.3377**<br>[1.2171; 1.4703] |
| **PIV** | **GLMM** | 1.0809<br>[0.9088; 1.2855] | 0.9888<br>[0.8095; 1.2078] | 1.0993<br>[0.9306; 1.2987] | 1.0377<br>[0.7809; 1.3791] | 1.0131<br>[0.9386; 1.0935] | 0.9003<br>[0.7917; 1.0238] | 0.9539<br>[0.8474; 1.0737] |
| | **GAMLSS** | 1.0700<br>[0.9020; 1.2693] | 0.9824<br>[0.7971; 1.2108] | 1.0766<br>[0.9082; 1.2763] | 1.0407<br>[0.7823; 1.3844] | 1.0035<br>[0.9371; 1.0746] | 0.9103<br>[0.7922; 1.0461] | 0.9426<br>[0.8332; 1.0664] |
| | **VGLM** | **1.8028**<br>[1.3324; 2.4391] | 1.0084<br>[0.7242; 1.4041] | 1.1896<br>[0.9771; 1.4482] | 1.0197<br>[0.7192; 1.4457] | 1.0658<br>[0.9721; 1.1686] | 0.9433<br>[0.7848; 1.1339] | 1.0161<br>[0.8615; 1.1985] |
| **RSV** | **GLMM** | 1.0399<br>[0.9706; 1.1141] | 0.9880<br>[0.9324; 1.0469] | 1.0250<br>[0.9707; 1.0824] | 0.9650<br>[0.8954; 1.0401] | **1.0742**<br>[1.0509; 1.0981] | 1.0089<br>[0.9764; 1.0425] | **0.9534**<br>[0.9166; 0.9916] |
| | **GAMLSS** | 1.0513<br>[0.9766; 1.1317] | 1.0003<br>[0.9375; 1.0673] | 1.0167<br>[0.9565; 1.0806] | 0.9422<br>[0.8693; 1.0212] | **1.0612**<br>[1.0393; 1.0835] | 1.0130<br>[0.9749; 1.0525] | **0.9345**<br>[0.8946; 0.9762] |
| | **VGLM** | 1.1018<br>[0.9863; 1.2308] | 0.9395<br>[0.8472; 1.0419] | 1.0658<br>[0.9943; 1.1425] | 0.9704<br>[0.8798; 1.0704] | **1.1404**<br>[1.1083; 1.1735] | **1.0594**<br>[1.0066; 1.1151] | **0.9370**<br>[0.8862; 0.9909] |
| **RV/EV** | **GLMM** | 1.1709<br>[0.9488; 1.4450] | 0.9656<br>[0.8339; 1.1182] | 0.8947<br>[0.6533; 1.2254] | 0.8526<br>[0.6790; 1.0706] | 0.9507<br>[0.8635; 1.0467] | 0.9787<br>[0.8627; 1.1103] | 0.9794<br>[0.8526; 1.1250] |
| | **GAMLSS** | 1.1427<br>[0.9378; 1.3923] | 0.9772<br>[0.8392; 1.1379] | 0.9081<br>[0.6581; 1.2531] | 0.8302<br>[0.6606; 1.0435] | 0.9718<br>[0.8892; 1.0620] | 1.0051<br>[0.8796; 1.1485] | 0.9682<br>[0.8387; 1.1176] |
| | **VGLM** | **1.8808**<br>[1.3242; 2.6714] | 1.3291<br>[0.9848; 1.7938] | 0.8944<br>[0.6303; 1.2692] | 1.0308<br>[0.7822; 1.3583] | 0.9633<br>[0.8552; 1.0851] | 1.1497<br>[0.9529; 1.3871] | 0.9823<br>[0.8094; 1.1920] |
| **SARS-CoV-2** | **GLMM** | 1.0888<br>[0.9922; 1.1949] | **1.0681**<br>[1.0085; 1.1312] | 1.0263<br>[0.9293; 1.1334] | **1.1468**<br>[1.0517; 1.2506] | 1.0157<br>[0.9814; 1.0511] | **1.1063**<br>[1.0645; 1.1499] | **1.0554**<br>[1.0046; 1.1087] |
| | **GAMLSS** | 1.0855<br>[0.9915; 1.1884] | **1.0676**<br>[1.0068; 1.1321] | 1.0473<br>[0.9463; 1.1591] | **1.1267**<br>[1.0295; 1.2331] | 1.0197<br>[0.9876; 1.0528] | **1.1089**<br>[1.0633; 1.1564] | 1.0464<br>[0.9953; 1.1000] |
| | **VGLM** | **1.1682**<br>[1.0170; 1.3419] | 1.0682<br>[0.9454; 1.2068] | 1.0441<br>[0.9299; 1.1723] | **1.3795**<br>[1.2403; 1.5342] | 1.0422<br>[0.9994; 1.0869] | **1.5275**<br>[1.4347; 1.6262] | **1.0918**<br>[1.0176; 1.1715] |
| **hCoV** | **GLMM** | 0.9741<br>[0.8907; 1.0652] | **1.0862**<br>[1.0111; 1.1669] | 1.0361<br>[0.9393; 1.1429] | 1.0667<br>[0.9505; 1.1971] | 1.0017<br>[0.9615; 1.0436] | 0.9730<br>[0.9138; 1.0360] | 1.0507<br>[0.9969; 1.1074] |
| | **GAMLSS** | 0.9665<br>[0.8836; 1.0571] | **1.0939**<br>[1.0131; 1.1812] | 1.0184<br>[0.9201; 1.1274] | 1.0574<br>[0.9410; 1.1881] | 0.9862<br>[0.9496; 1.0243] | 0.9669<br>[0.9030; 1.0352] | 1.0250<br>[0.9689; 1.0843] |
| | **VGLM** | **1.1811**<br>[1.0105; 1.3804] | **1.3488**<br>[1.1711; 1.5533] | 1.0619<br>[0.9454; 1.1928] | 1.1371<br>[0.9795; 1.3201] | 0.9975<br>[0.9482; 1.0493] | 1.0022<br>[0.9132; 1.0999] | **1.2181**<br>[1.1267; 1.3170] |

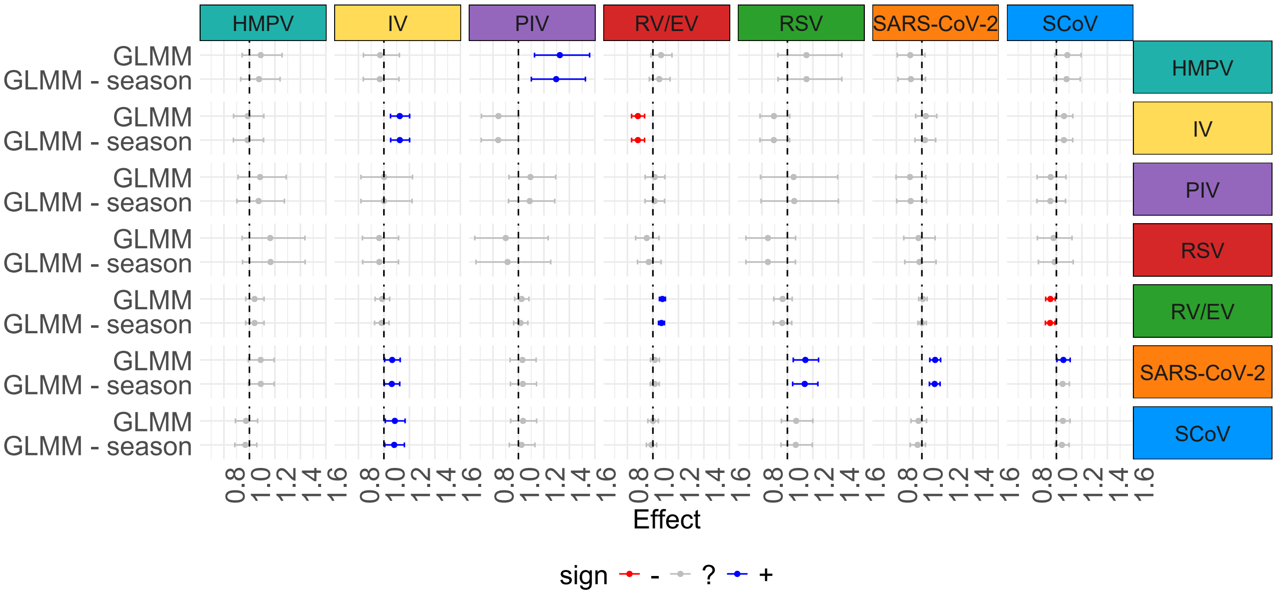


Figure E.1: Comparative plot of lagged incidence rate ratios (IRRs) for the GLMM model with inclusion and exclusion of the effect of different epidemiological seasons. Rows indicate the outcome disease, and columns indicate the lagged predictor disease. In blue are highlighted significant positive effects, in red significant negative effects, and in gray non-significant effects.

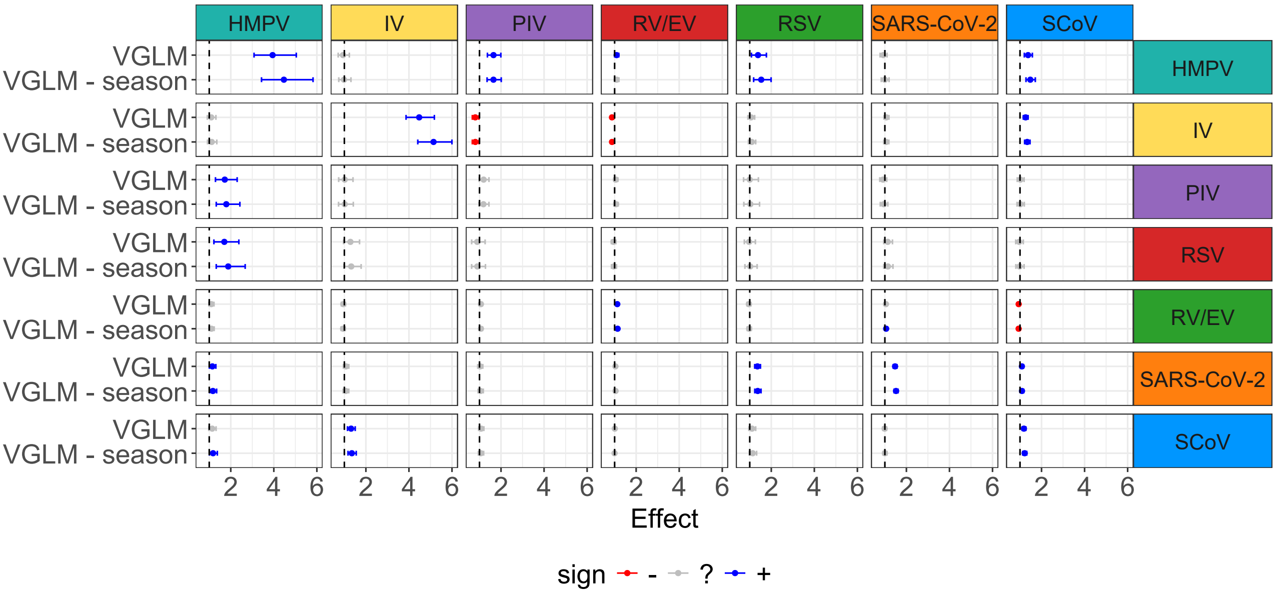


Figure E.2: Comparative plot of lagged incidence rate ratios (IRRs) for the VGLM model with inclusion and exclusion of the effect of different epidemiological seasons. Rows indicate the outcome disease, and columns indicate the lagged predictor disease. In blue are highlighted significant positive effects, in red significant negative effects, and in gray non-significant effects.

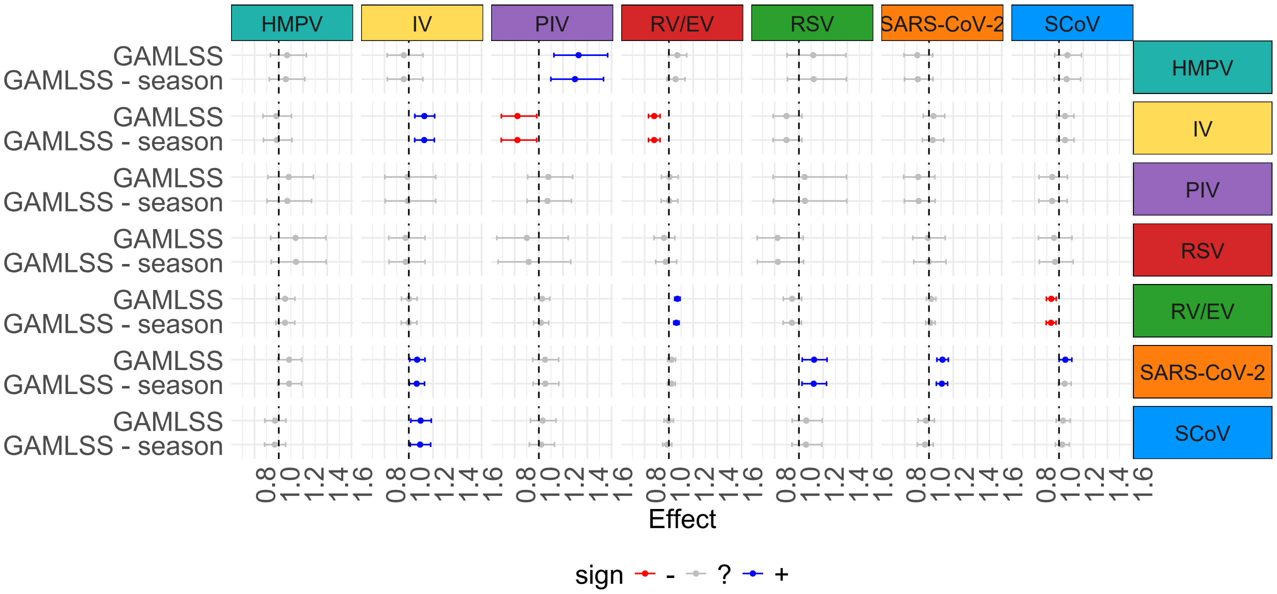


Figure E.3: Comparative plot of lagged incidence rate ratios (IRRs) for the GAMLSS model with inclusion and exclusion of the effect of different epidemiological seasons. Rows indicate the outcome disease, and columns indicate the lagged predictor disease. In blue are highlighted significant positive effects, in red significant negative effects, and in gray non-significant effects

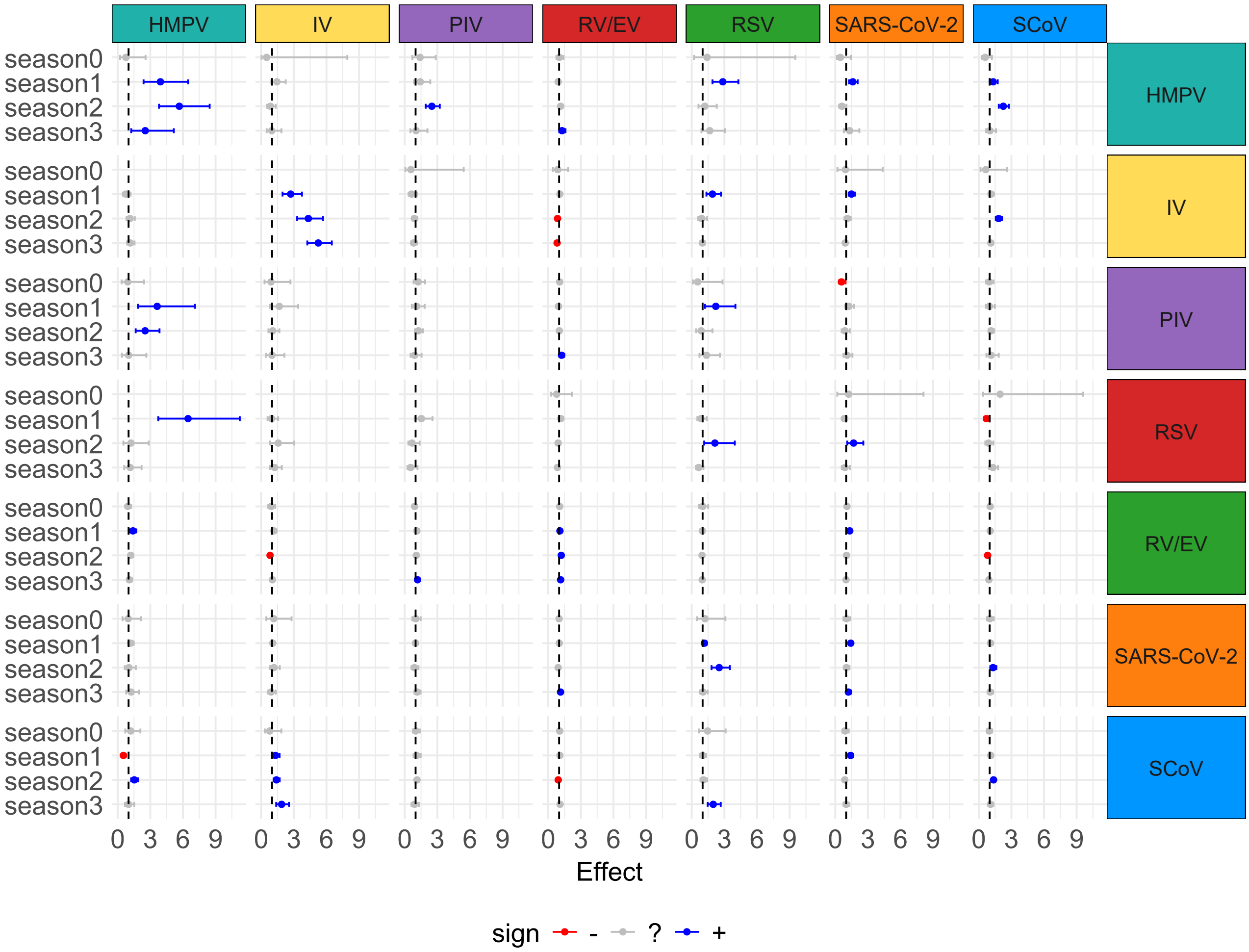


Figure E.4: Comparative plot of lagged incidence rate ratios (IRRs) for the VGLM model with season-specific cross-pathogen effects. Rows indicate the outcome disease, and columns indicate the lagged predictor disease. In blue are highlighted significant positive effects, in red significant negative effects, and in gray non-significant effects

### F Endemic-Epidemic

#### F.1 Model Selection

##### Choice of time and seasonal component

We evaluated a range of alternative specifications for the temporal trend and seasonal component in the E&E models, including linear time, natural cubic splines with 1–5 degrees of freedom, sine/cosine Fourier terms, and standardised temperature covariates. Model fit was assessed using AIC and BIC, with convergence considered as an additional criterion.

Figure F.1 summarises the results. Across pathogens, fit improved monotonically with spline complexity, with the lowest AIC/BIC typically observed at 4–5 degrees of freedom. At these df, adding Fourier terms further reduced AIC/BIC, while temperature provided no systematic gains and in some cases impaired convergence. Taken together, we adopt natural cubic splines with 5 df and Fourier sine/cosine terms as the common temporal specification for the E&E models, omitting temperature, as shown in (F.1), where $B_k(t*)$ are the basis functions of a natural cubic spline with 5 degrees of freedom, $t*$ is the standardised week index, and the sine/cosine terms capture annual seasonality. The coefficients $\beta_{k,r,d}$ and $\gamma_{1,r,d}, \gamma_{2,r,d}$ are disease- and region-specific. This matches the configuration chosen for the GLM-based models and thereby ensures comparability across frameworks.

$$f_{\text{time},r,d}(t*) = \sum_{k=1}^{5} \beta_{k,r,d} B_k(t*) + \gamma_{1,r,d} \sin\left(\tfrac{2\pi t}{52}\right) + \gamma_{2,r,d} \cos\left(\tfrac{2\pi t}{52}\right), \tag{F.1}$$

##### Family selection

We compared alternative distributions (Poisson, NegBin1, and NegBinM) across all pathogens. For each pathogen, model fit was evaluated using AIC and BIC, retaining only models that converged successfully.

As shown in Table F.1, Poisson models consistently had the poorest fit, while negative binomial families substantially improved AIC and BIC values. NegBin1 provided the best fit for most pathogens (hMPV, IV, PIV, RSV, SARS-CoV-2), while NegBinM occasionally achieved lower AIC values (hCoV, RV/EV). However, NegBinM models were less stable, with several convergence issues and missing estimates.

When aggregated across pathogens, both negative binomial families outperformed Poisson, confirming substantial overdispersion in the data. Given its consistent performance and stability, we proceeded with NegBin1 as the model family for all pathogens.

Table F.1: Model fit comparison (Poisson vs. NegBin1 vs. NegBinM) across diseases.

| | **Poisson** | | | **NegBin1** | | | **NegBinM** | | |
|---|---|---|---|---|---|---|---|---|---|
| | **AIC** | **BIC** | **Converged** | **AIC** | **BIC** | **Converged** | **AIC** | **BIC** | **Converged** |
| **hMPV** | 1698 | 1789 | Yes | 1676 | 1774 | Yes | NA | NA | No |
| **IV** | 2476 | 2567 | Yes | 2414 | 2511 | Yes | NA | NA | No |
| **PIV** | 2433 | 2525 | Yes | 2412 | 2510 | Yes | NA | NA | No |
| **RV/EV** | 8413 | 8504 | Yes | 8164 | 8261 | Yes | 8048 | 8286 | Yes |
| **RSV** | 1635 | 1726 | Yes | 1624 | 1722 | Yes | NA | NA | No |
| **SARS-CoV-2** | 4731 | 4823 | Yes | 4600 | 4697 | Yes | NA | NA | No |
| **hCoV** | 4706 | 4797 | Yes | 4547 | 4645 | Yes | 4433 | 4670 | Yes |

##### Inclusion of Lagged Counts

Although the endemic specification consistently yielded the best fit for most pathogens (Table F.2), we also estimated models with lagged counts placed in the epidemic and combined endemic–epidemic components. This was done to enable interpretation of cross-pathogen effects within the epidemic compartment, which is most directly comparable to the regression-based analyses. Hence, while the endemic placement improved fit, the epidemic terms provide the epidemiologically relevant estimates of potential transmission-like dependencies.

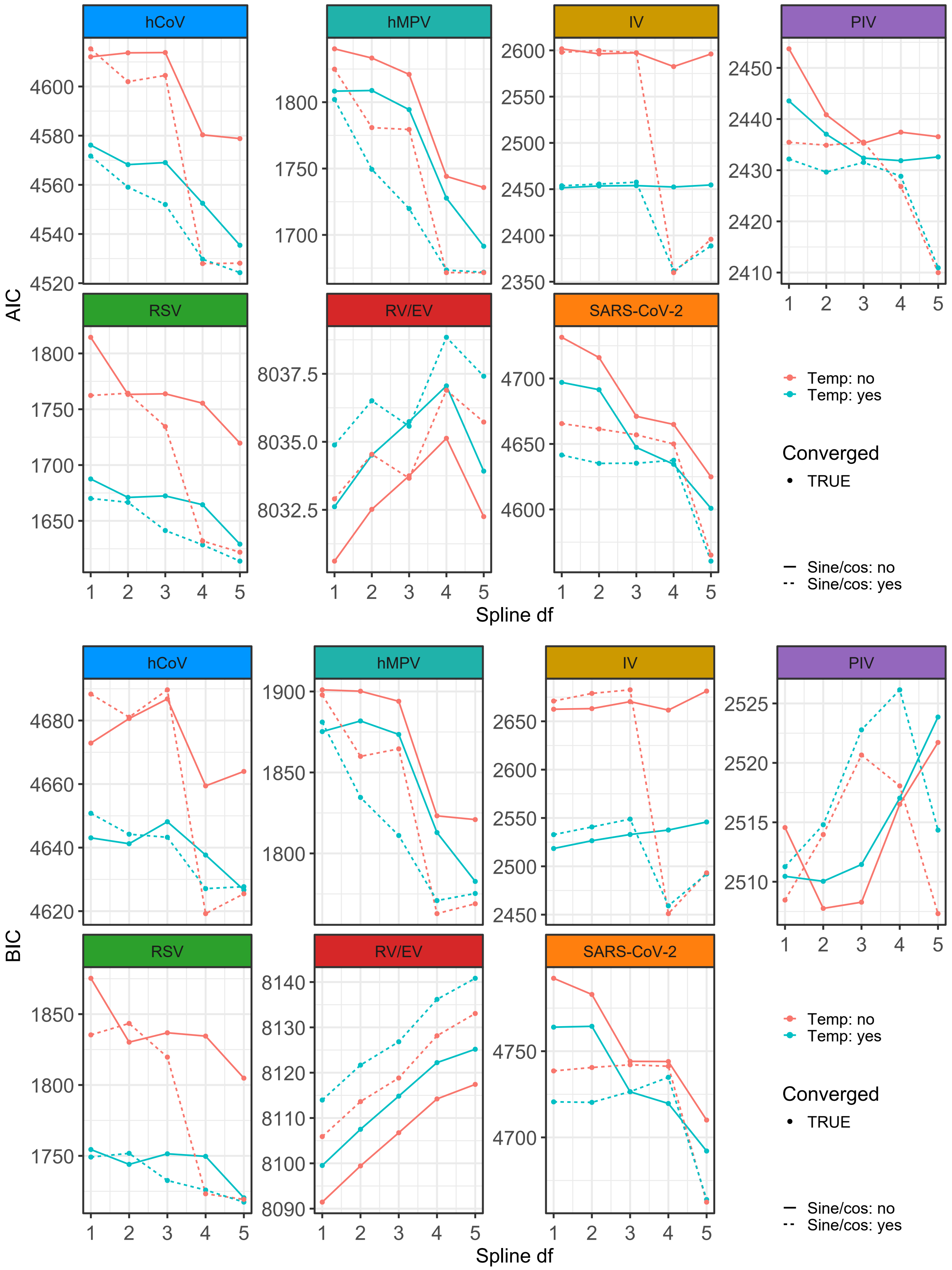


Figure F.1: Model fit (upper panel: AIC and lower panel: BIC) for temporal specifications across spline degrees, temperature inclusion, and Fourier sine/cosine terms, stratified by pathogen. Points indicate convergence status (filled = converged, open = not converged).

Table F.2: Model fit comparison (lagged counts in endemic, epidemic or both components) across diseases.

| | epidemic | | endemic | | both | |
|---|---|---|---|---|---|---|
| | **AIC** | **BIC** | **AIC** | **BIC** | **AIC** | **BIC** |
| **hMPV** | 1676 | 1780 | 1574 | 1678 | 1577 | 1723 |
| **IV** | 2415 | 2519 | 2339 | 2443 | 2346 | 2492 |
| **PIV** | 2414 | 2517 | 2302 | 2405 | 2305 | 2451 |
| **RV/EV** | 8160 | 8263 | 8054 | 8157 | 8052 | 8198 |
| **RSV** | 1626 | 1730 | 1541 | 1644 | 1551 | 1697 |
| **SARS-CoV-2** | 4599 | 4703 | 4498 | 4602 | 4503 | 4649 |
| **hCoV** | 4549 | 4652 | 4384 | 4487 | 4388 | 4534 |

#### F.2 Results

#### Epidemic vs endemic contributions

Across pathogens, the endemic component accounted for most of the fitted mean counts, though its relative importance varied (Table F.3). For RSV, hMPV, and PIV, the epidemic share averaged below 10%, indicating predominantly endemic dynamics. In contrast, influenza, SARS-CoV-2, and seasonal coronaviruses showed epidemic contributions around 15–20% on average, while RV/EV stood out with nearly 40% and occasional peaks above 90%. These ranges reflect the strong temporal variability: during low-incidence periods the epidemic part was negligible, but during peaks it could dominate locally.

Table F.3: Average endemic and epidemic contributions (%) to fitted mean counts in the EE models, with observed ranges in parentheses.

| **configuration:** | epidemic | | endemic | | both | |
|---|---|---|---|---|---|---|
| **contribution:** | **end (%)** | **epi (%)** | **end (%)** | **epi (%)** | **end (%)** | **epi (%)** |
| **hMPV** | 93.6 | 6.4 | 92.4 | 7.6 | 93.1 | 7.0 |
| **IV** | 84.6 | 15.4 | 83.9 | 16.1 | 84.5 | 15.5 |
| **PIV** | 93.6 | 6.4 | 92.6 | 7.4 | 93.9 | 6.1 |
| **RV/EV** | 62.5 | 37.5 | 58.4 | 41.6 | 62.1 | 37.9 |
| **RSV** | 95.5 | 4.5 | 94.5 | 5.5 | 95.6 | 4.4 |
| **SARS-CoV-2** | 81.6 | 18.4 | 77.9 | 22.1 | 81.3 | 18.7 |
| **hCoV** | 85.7 | 14.3 | 82.1 | 17.9 | 85.5 | 14.5 |

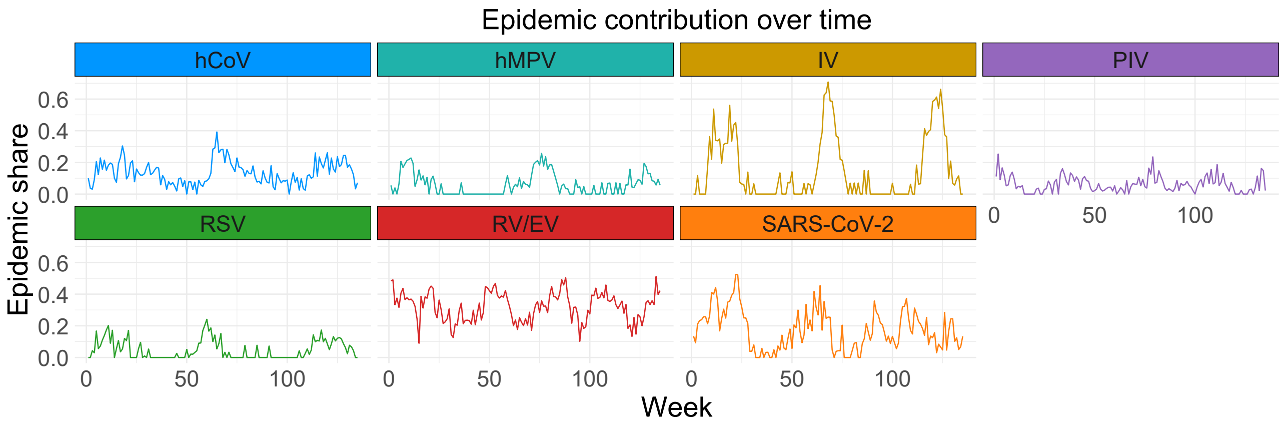


Figure F.2: Weekly proportion of fitted mean incidence attributed to the epidemic component in the EE models.

Table F.4: Summary of the RRs for cross-pathogen interactions from the E&E models across all pathogen pairs with corresponding 95% confidence intervals by model configuration and component.

| | | | Target | | | | | | |
|---|---|---|---|---|---|---|---|---|---|
| **Predictor** | **Configuration** | | **hMPV** | **IV** | **PIV** | **RV/EV** | **RSV** | **SARS-COV-2** | **SCOV** |
| **hMPV** | end | end | | 1.3052 [0.9698; 1.7565] | 1.4587 [1.1853; 1.7950] | 1.3374 [1.1297; 1.5832] | 1.4952 [1.1231; 1.9906] | 1.1589 [0.9498; 1.4139] | 1.4136 [1.2100; 1.6516] |
| | epi | epi | | 1.1211 [0.8812; 1.4263] | 1.3461 [0.8309; 2.1810] | 1.1148 [1.0035; 1.2385] | 1.3193 [0.7172; 2.4269] | 1.1302 [0.9197; 1.3888] | 1.4718 [1.2137; 1.7848] |
| | both | epi | | 1.0914 [0.8296; 1.4359] | 0.9241 [0.3679; 2.3215] | 0.9972 [0.7920; 1.2554] | 1.2803 [0.3703; 4.4266] | 1.0593 [0.7608; 1.4749] | 1.3987 [0.8577; 2.2809] |
| | both | end | | 1.2848 [0.9274; 1.7799] | 1.4668 [1.1896; 1.8085] | 1.3632 [1.0909; 1.7034] | 1.4758 [1.1012; 1.9778] | 1.1502 [0.9079; 1.4570] | 1.3010 [1.0098; 1.6761] |
| **IV** | end | end | 1.3863 [1.1912; 1.6133] | | 0.8906 [0.7324; 1.0831] | 0.9549 [0.8480; 1.0752] | 1.2596 [1.0772; 1.4729] | 1.1951 [1.0589; 1.3490] | 1.5821 [1.4339; 1.7456] |
| | epi | epi | 1.1059 [0.8509; 1.4375] | | 0.3747 [0.1226; 1.1457] | 0.9520 [0.8732; 1.0378] | 1.1183 [0.8216; 1.5222] | 1.0874 [0.9619; 1.2293] | 1.3101 [1.1629; 1.4759] |
| | both | epi | 0.9623 [0.6573; 1.4088] | | 0.1758 [0.0044; 7.1067] | 0.9156 [0.7990; 1.0493] | 0.8670 [0.3019; 2.4899] | 1.0513 [0.8932; 1.2374] | 1.0633 [0.8222; 1.3750] |
| | both | end | 1.4370 [1.2216; 1.6903] | | 0.9416 [0.7835; 1.1315] | 1.0174 [0.9008; 1.1491] | 1.2663 [1.0714; 1.4966] | 1.1888 [1.0421; 1.3563] | 1.5780 [1.4038; 1.7740] |
| **PIV** | end | end | 1.3624 [1.0779; 1.7219] | 0.9936 [0.7078; 1.3949] | | 1.6494 [1.4267; 1.9069] | 1.0059 [0.7061; 1.4331] | 0.9200 [0.7307; 1.1582] | 1.2411 [1.0388; 1.4828] |
| | epi | epi | 1.0246 [0.6705; 1.5659] | 0.9431 [0.6723; 1.3230] | | 1.1953 [1.0714; 1.3336] | 0.9472 [0.4102; 2.1876] | 1.0165 [0.7866; 1.3136] | 1.1045 [0.8339; 1.4628] |
| | both | epi | 0.7821 [0.3824; 1.5995] | 0.9370 [0.6470; 1.3570] | | 1.0764 [0.9151; 1.2662] | 0.8940 [0.1395; 5.7307] | 1.2016 [0.8621; 1.6747] | 0.8611 [0.4816; 1.5396] |
| | both | end | 1.4563 [1.1400; 1.8604] | 1.0359 [0.7361; 1.4577] | | 1.5378 [1.2806; 1.8466] | 1.0252 [0.7048; 1.4913] | 0.8616 [0.6494; 1.1433] | 1.2771 [1.0523; 1.5499] |
| **RV/EV** | end | end | 1.2263 [1.1265; 1.3349] | 1.0327 [0.9344; 1.1414] | 1.2812 [1.2074; 1.3596] | | 1.0851 [0.9981; 1.1797] | 1.2552 [1.1988; 1.3143] | 1.1753 [1.1171; 1.2365] |
| | epi | epi | 1.2273 [1.0572; 1.4246] | 1.0770 [0.9886; 1.1732] | 1.1926 [1.0358; 1.3732] | | 1.1567 [0.9168; 1.4594] | 1.0883 [1.0325; 1.1472] | 1.0207 [0.9316; 1.1184] |
| | both | epi | 1.1470 [0.9396; 1.4002] | 1.0809 [0.9860; 1.1849] | 0.9295 [0.6833; 1.2645] | | 1.1101 [0.8089; 1.5235] | 0.9648 [0.8828; 1.0544] | 0.8932 [0.7676; 1.0394] |
| | both | end | 1.1920 [1.0810; 1.3144] | 1.0105 [0.9110; 1.1208] | 1.2923 [1.2156; 1.3738] | | 1.0817 [0.9929; 1.1785] | 1.2702 [1.2073; 1.3364] | 1.1919 [1.1308; 1.2563] |
| **RSV** | end | end | 1.7380 [1.2941; 2.3342] | 2.0293 [1.5414; 2.6717] | 0.9275 [0.6366; 1.3515] | 1.1872 [0.9700; 1.4530] | | 1.3322 [1.0749; 1.6512] | 1.2975 [1.0476; 1.6071] |
| | epi | epi | 1.2363 [0.6513; 2.3466] | 1.1618 [0.9150; 1.4753] | 0.8545 [0.4473; 1.6324] | 1.1164 [0.9730; 1.2809] | | 1.0887 [0.8838; 1.3411] | 1.2076 [0.8788; 1.6593] |
| | both | epi | 1.0709 [0.4617; 2.4838] | 1.0359 [0.7735; 1.3873] | 0.9801 [0.3676; 2.6129] | 1.0470 [0.8358; 1.3115] | | 0.9815 [0.7034; 1.3696] | 1.2726 [0.8372; 1.9345] |
| | both | end | 1.7344 [1.2809; 2.3486] | 2.0247 [1.5260; 2.6864] | 0.8908 [0.5628; 1.4099] | 1.1515 [0.8676; 1.5281] | | 1.3416 [1.0455; 1.7215] | 1.2418 [0.9850; 1.5655] |
| **SARS-COV-2** | end | end | 1.1473 [1.0009; 1.3151] | 1.0988 [0.9760; 1.2371] | 1.0247 [0.9146; 1.1480] | 1.1613 [1.0823; 1.2461] | 1.2659 [1.1078; 1.4466] | | 1.1506 [1.0541; 1.2559] |
| | epi | epi | 1.0371 [0.8498; 1.2656] | 1.0124 [0.9196; 1.1146] | 1.3018 [1.0649; 1.5915] | 1.0645 [1.0191; 1.1119] | 1.0146 [0.7998; 1.2872] | | 1.0636 [0.9553; 1.1841] |
| | both | epi | 0.9766 [0.7204; 1.3239] | 1.0038 [0.9030; 1.1158] | 1.4086 [1.1073; 1.7918] | 1.0819 [1.0201; 1.1474] | 0.8922 [0.5376; 1.4806] | | 1.1091 [0.9568; 1.2855] |
| | both | end | 1.1647 [1.0130; 1.3392] | 1.1072 [0.9795; 1.2515] | 0.9800 [0.8670; 1.1077] | 1.0953 [1.0090; 1.1890] | 1.2626 [1.1018; 1.4467] | | 1.1357 [1.0374; 1.2432] |
| **SCOV** | end | end | 1.1777 [1.0289; 1.3481] | 1.5591 [1.3632; 1.7831] | 1.1661 [1.0349; 1.3139] | 1.3072 [1.2052; 1.4177] | 1.3753 [1.1801; 1.6027] | 1.1303 [1.0186; 1.2542] | |
| | epi | epi | 1.2367 [1.0386; 1.4725] | 1.1803 [1.0560; 1.3192] | 1.2162 [0.9403; 1.5731] | 1.1080 [1.0456; 1.1741] | 1.0148 [0.6887; 1.4952] | 1.0989 [0.9887; 1.2214] | |
| | both | epi | 1.2929 [1.0535; 1.5868] | 1.1360 [1.0011; 1.2892] | 1.3928 [0.9359; 2.0729] | 0.9470 [0.8426; 1.0642] | 0.4782 [0.1089; 2.0998] | 1.1057 [0.9549; 1.2803] | |
| | both | end | 1.0511 [0.8883; 1.2437] | 1.5251 [1.3231; 1.7579] | 1.1386 [1.0041; 1.2911] | 1.3564 [1.2316; 1.4938] | 1.4091 [1.2140; 1.6357] | 1.0923 [0.9650; 1.2364] | |

#### F.3 Model diagnostics

We assessed the adequacy of the E&E models using Pearson residuals, which standardise observed counts by the fitted variance under the negative binomial assumption. Diagnostics included mean squared Pearson residuals (dispersion fit), QQ-plots (distributional adequacy), residuals versus fitted values (mean–variance calibration), autocorrelation (short-term dependence), and median residuals over time (temporal bias).

##### Residual scale

Figure F.3 shows mean squared Pearson residuals by pathogen and model variant. Values near one indicate adequate variance fit. For the model variants in which the lagged counts were added to the endemic component only and to both components, the residual scale was near one for PIV (end: 0.99, both: 1.00) and hCoV (end: 0.98, both: 0.99), mildly overdispersed for RV/EV (end: 1.05, both: 1.06) and SARS-CoV-2 (end: 1.04, both: 1.05), clearly underdispersed for hMPV (end: 0.85, epi: 0.85) and RSV (end: 0.80, both: 0.80), and overdispersed for IV (end: 1.13, both: 1.14). The model variant in whoch the lagged counts were only added to the epidemic component increased the scale for all pathogens (hMPV: 0.97, IV: 1.28, PIV: 1.05, RSV: 0.97, RV/EV: 1.10, SARS-CoV-2: 1.10, hCoV: 1.09), giving the weakest variance fit, especially for IV.

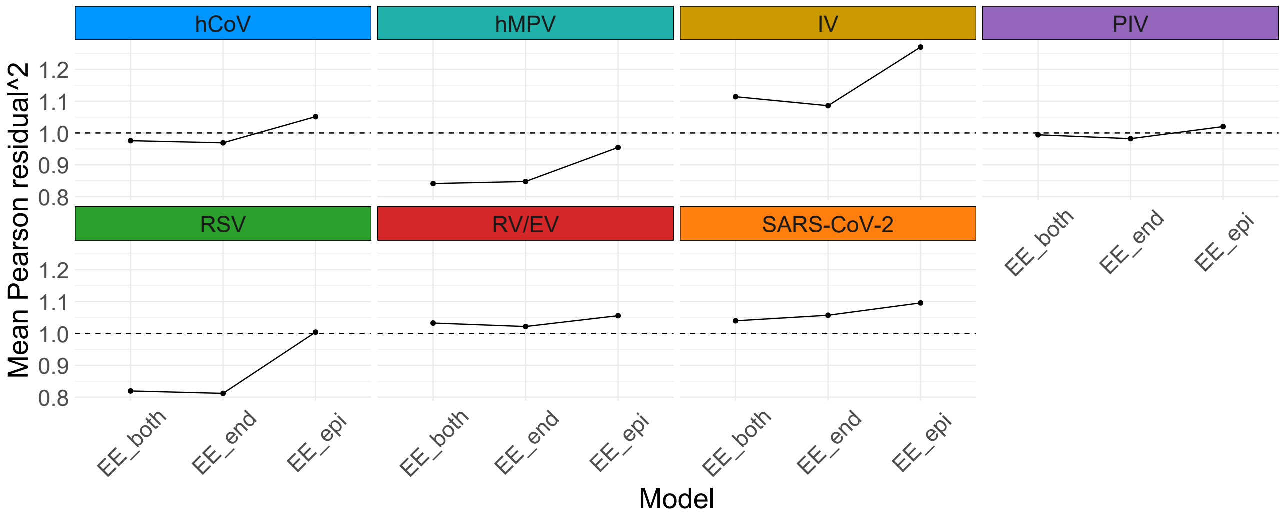


Figure F.3: Mean squared Pearson residuals by pathogen and E&E model variant. The dashed line marks the target value of one.

##### QQ-plots

QQ plots of Pearson residuals (Figure F.4) revealed heavy upper tails across pathogens, reflecting outbreak peaks that are difficult to reproduce with a constant dispersion parameter. The central bulk of the distribution aligned well with the normal reference, indicating adequate fit for routine incidence. This tail behaviour is expected in infectious disease surveillance data, where a single constant dispersion parameter must reconcile routine baseline variation with rare outbreak extremes.

##### Residuals versus fitted values

Binned residual plots (Figure F.5) showed residuals centred around zero, with minor deviations at low fitted means and during peaks of RV/EV and SARS-CoV-2. Overall, the negative binomial variance captured the mean–variance relationship satisfactorily.

##### Residual autocorrelation

Figure F.6 summarizes the lag-1 autocorrelation of Pearson residuals across units. Medians were close to zero for all pathogens and model variants, indicating that the autoregressive component generally accounted for short-term dependence. The interquartile ranges were narrow and centered around zero, further supporting the absence of systematic residual autocorrelation.

A subset of units, however, showed elevated positive autocorrelations, most notably for hMPV, IV, and RSV. These outliers suggest that in some regions and age groups, local epidemic persistence was not fully captured by the autoregressive structure. Overall, the results indicate adequate modelling of short-term dependence at the population level, with residual autocorrelation confined to a limited number of units.

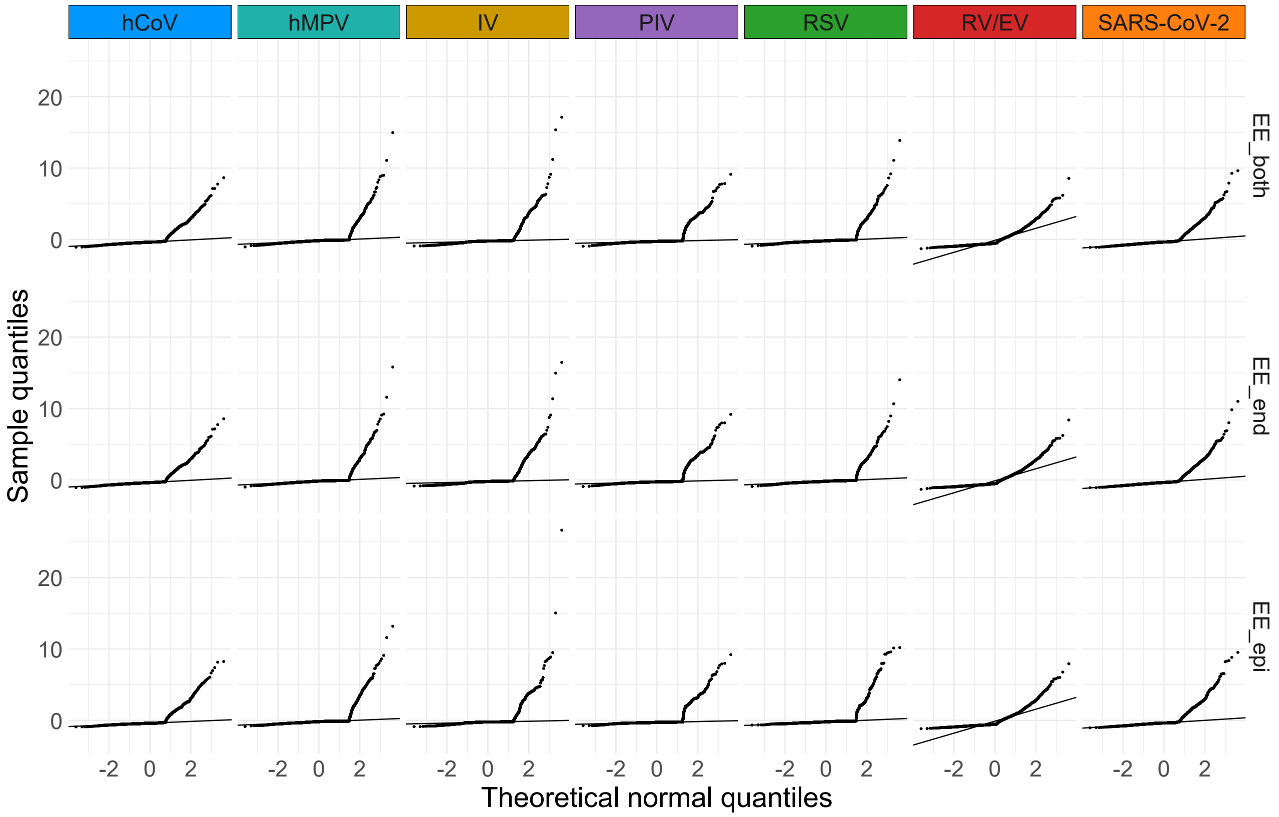


Figure F.4: QQ plots of Pearson residuals by pathogen and E&E model variant. The solid line is the reference.

##### Residuals over time

Median residuals over time (Figure F.7) highlighted different patterns across pathogens. For IV, RV/EV, and SARS-CoV-2, deviations were short-lived and coincided with epidemic peaks. By contrast, hMPV, RSV, and hCoV showed a tendency for residuals to remain slightly below zero, suggesting mild systematic overprediction of incidence. Overall, the trajectories indicate that while the E&E models did not suffer from large-scale temporal drift, some pathogens exhibited persistent small biases in baseline fit.

#### Summary of residual diagnostics

The diagnostics show that the `EE_end` and `EE_both` variants generally provided better dispersion and autocorrelation fit than the `EE_epi` specification. QQ-plots indicated that routine incidence was well represented, with heavy upper tails reflecting rare outbreak peaks that a constant dispersion parameter cannot fully capture. Residuals versus fitted values showed adequate calibration across most of the fitted range, though deviations appeared at very low means and during epidemic spikes. Median residuals over time revealed pathogen-specific patterns: influenza, RV/EV, and SARS-CoV-2 exhibited transient deviations during peaks, while hMPV, RSV, and hCoV tended to remain slightly below zero, suggesting mild systematic overprediction. Lag-1 autocorrelations were centered near zero, with only a few units showing elevated values. Overall, the EE models achieved an adequate fit for inference on cross-pathogen associations, with remaining deviations reflecting the inherent difficulty of modelling both endemic baselines and rare epidemic extremes within a parsimonious framework.

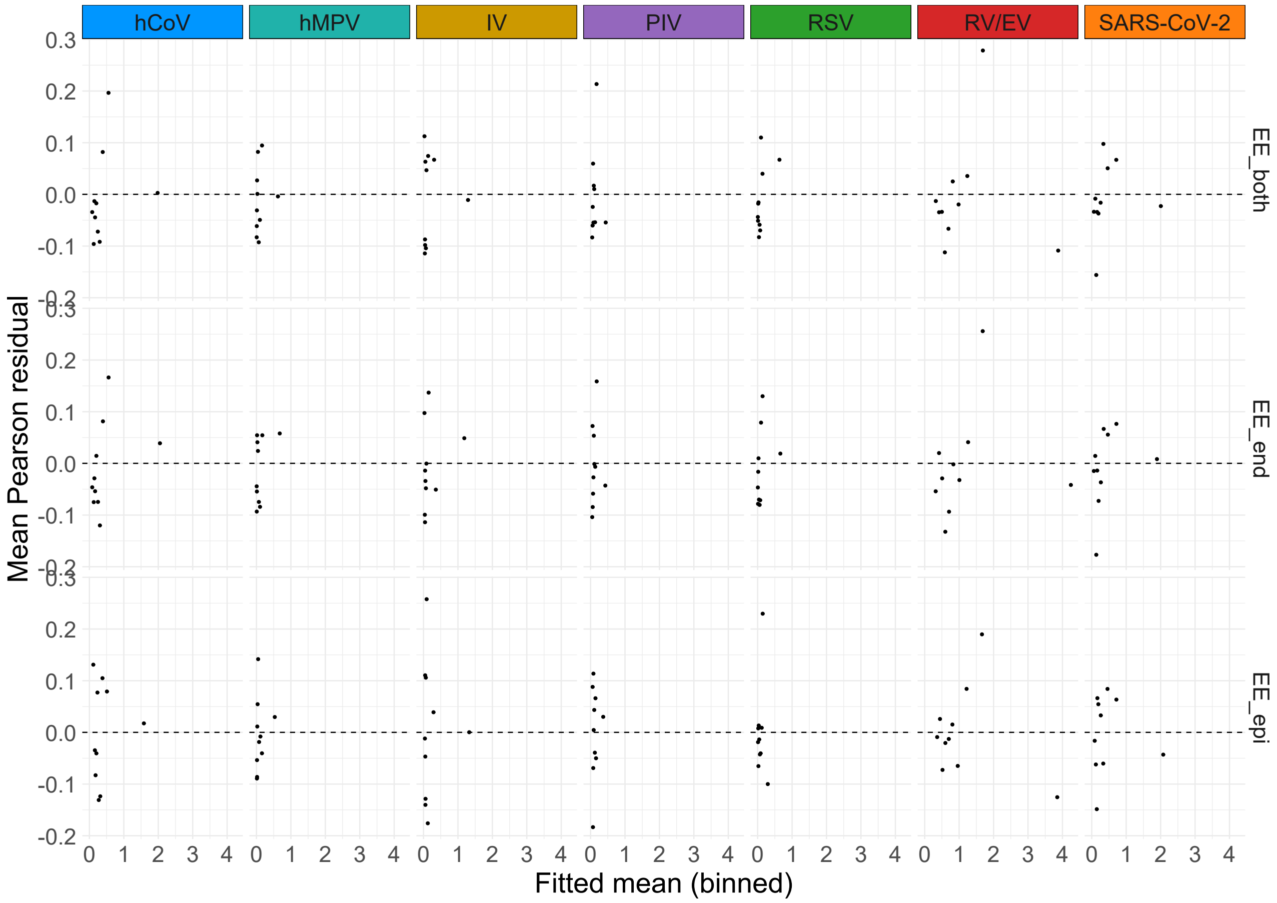


Figure F.5: Binned Pearson residuals versus fitted values by pathogen and EE model variant.

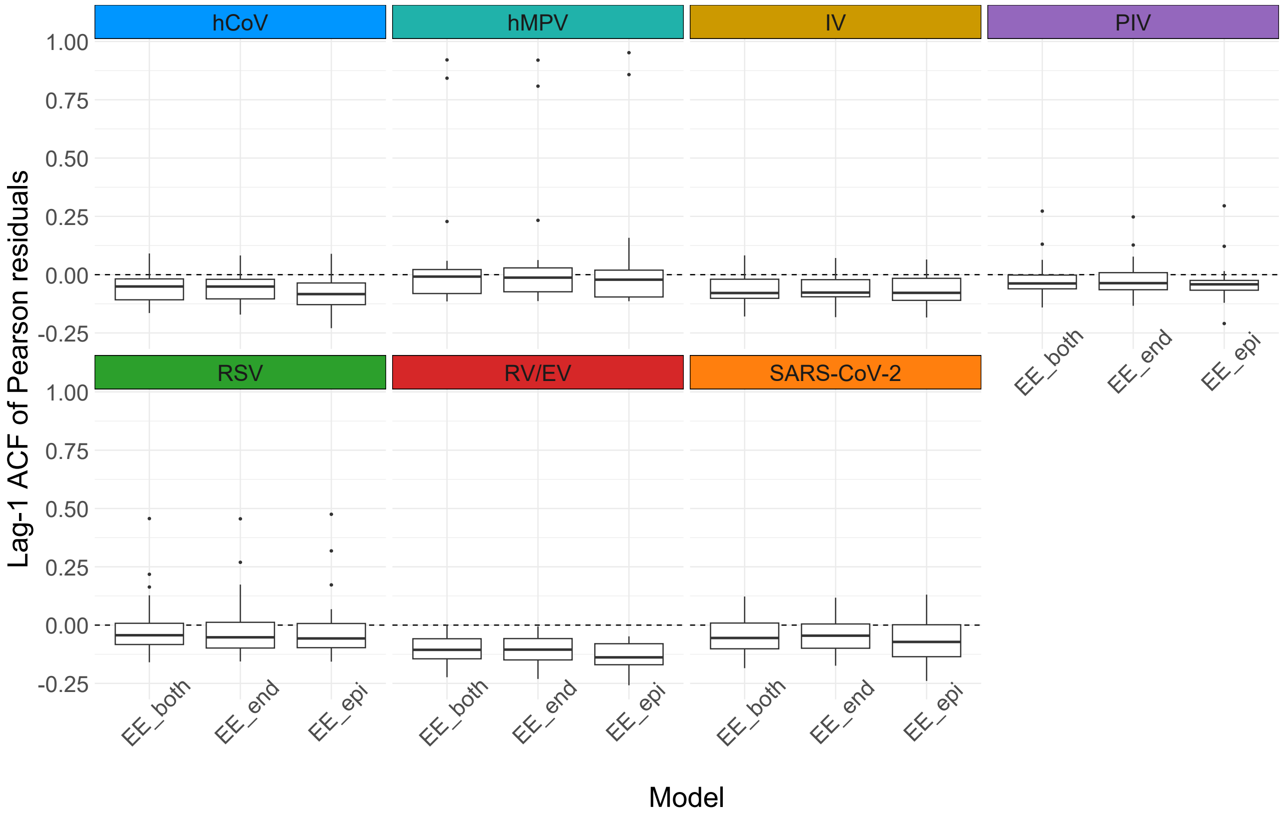


Figure F.6: Lag-1 autocorrelation of Pearson residuals by pathogen and E&E model variant. Boxplots summarize the distribution across units.

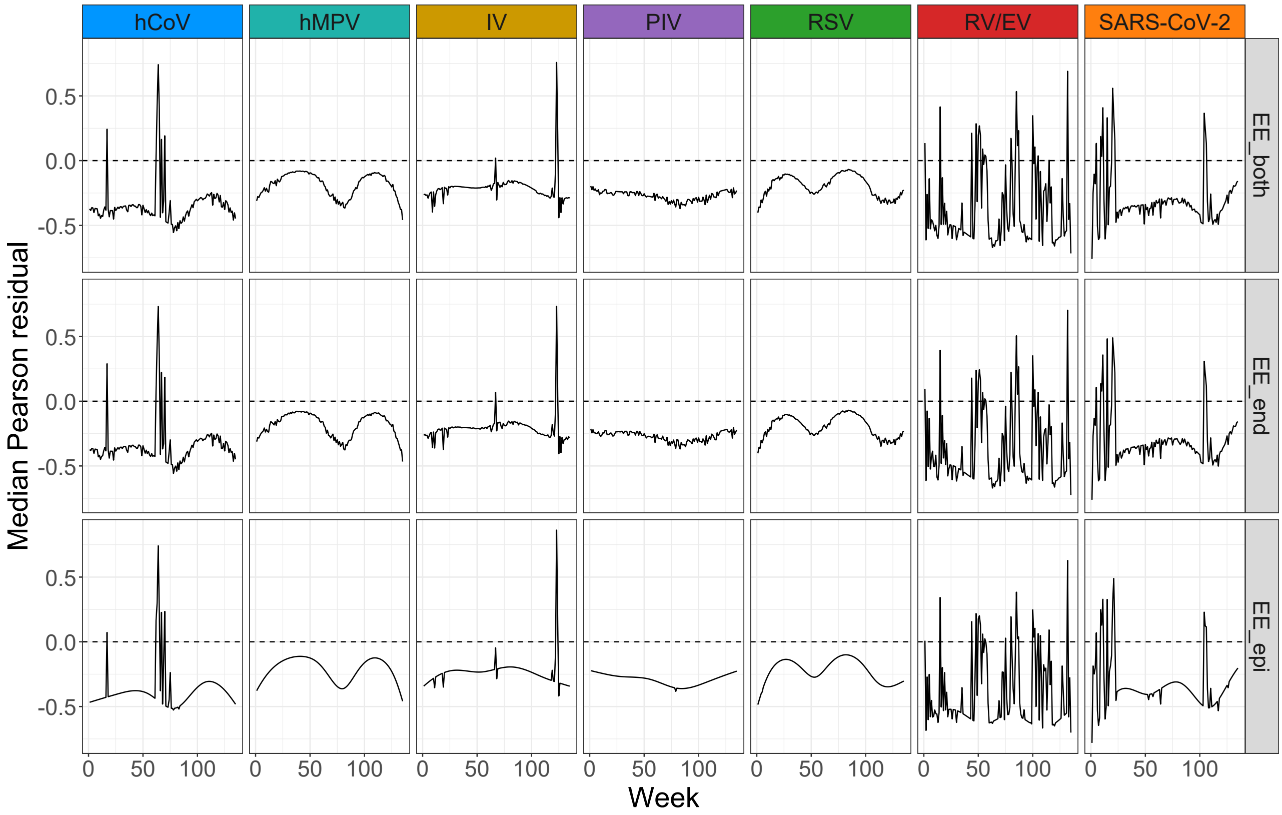


Figure F.7: Median Pearson residuals over time by pathogen and E&E model variant.

### G DLNM

#### G.1 Model Details

Let $\ell$ denote the lag, representing the number of weeks in the past. A set of basis functions $B_{\text{lag},k}(\ell)$ is defined for $k = 1, \ldots, K_{\text{lag}}$, where $K_{\text{lag}}$ is the number of B-spline basis functions along the lag dimension. This flexible functional form allows for a potentially non-linear relationship between the lagged effect and time, enabling the model to capture complex, time-varying impacts of past disease counts.

Similarly, a second set of basis functions $B_{\text{var},m}(x)$ for $m = 1, \ldots, K_{\text{var}}$ is defined over the exposure dimension, where $x$ denotes the lagged value of the predictor variable (e.g., weekly case counts of another pathogen).

Combining both dimensions yields a two-dimensional *cross-basis* expansion, which represents the joint effect of exposure level and lag on the response variable. For pathogen $d$ and exposure pathogen $d'$, this cross-basis can be expressed as

$$\mathbf{CB}_{d',r,a,t} = \sum_{\ell=0}^{L} B_{\text{lag}}(\ell)\, B_{\text{var}}\big(Y_{d',r,a,t-\ell}\big), \tag{G.1}$$

where $L$ is the maximum lag (here, $L = 8$ weeks). The combined term $\mathbf{CB}_{d',r,a,t}$ serves as a structured set of predictors in the linear predictor of the model (Equation 10 in the main text).

Formally, let $b_{\text{lag}}(\ell) \in \mathbb{R}^{K_{\text{lag}}}$ and $b_{\text{var}}(x) \in \mathbb{R}^{K_{\text{var}}}$ denote the evaluated basis vectors for lag and exposure, respectively. The corresponding coefficient matrix $\boldsymbol{\Theta}_{d,d'} \in \mathbb{R}^{K_{\text{lag}} \times K_{\text{var}}}$ links these two dimensions. The contribution of pathogen $d'$ to the predictor for pathogen $d$ can thus be written as

$$f_{d,d'}\big(\mathbf{CB}_{d',r,a,t}\big) = \sum_{\ell=0}^{L} b_{\text{lag}}(\ell)^{\top} \boldsymbol{\Theta}_{d,d'} b_{\text{var}}\big(Y_{d',r,a,t-\ell}\big). \tag{G.2}$$

The smooth bivariate function

$$f_{d,d'}(x,\ell) = b_{\text{lag}}(\ell)^{\top} \boldsymbol{\Theta}_{d,d'} b_{\text{var}}(x) \tag{G.3}$$

defines the lag–response surface, describing the combined effect of exposure level $x$ at lag $\ell$ on the expected mean $\mu_{d,r,a,t}$. The corresponding relative risk surface is obtained as

$$RR_{d,d'}(x,\ell) = \exp\big(f_{d,d'}(x,\ell)\big), \tag{G.4}$$

allowing the derivation of both cumulative and lag-specific relative risks.

For all models, we considered a maximum lag of $L = 8$ weeks, with B-splines of degree 1 used for the lag dimension and a linear exposure basis. Quadratic and cubic alternatives were evaluated in sensitivity analyses. The cross-basis design and estimation were implemented using the `dlnm` package in R, following the framework of Gasparrini et al. (2010, 2011).

#### G.2 Model Selection Process

We compared B-spline lag bases over a small grid of degrees $\{1, 3\}$ and degrees of freedom $\{2, 3, 4, 5\}$, keeping only valid pairs with df $\geq p + 1$. Models were ranked by AIC and BIC. To limit the candidate set, we varied the flexibility of the exposure–response function between a simple linear term (degree $= 1$) and a cubic spline (degree $= 3$), combined with lag dimensions of 2–5 degrees of freedom. We did not include quadratic exposure–response terms (degree $= 2$), as these impose a symmetric parabolic shape that is rarely appropriate for pathogen incidence data and are not commonly used in DLNM applications. The comparison between linear and cubic forms provided a balance between parsimony and flexibility while avoiding unnecessarily constrained intermediate shapes.

The linear lag basis with df $= 2$ (deg1_df2) minimized both AIC and BIC. Higher flexibility (larger degree or df) worsened fit and did not change substantive conclusions. We therefore retained deg1_df2 as the primary specification and report others as sensitivity checks.

#### G.3 Model diagnostics

We assessed the adequacy of the DLNM models using randomized quantile residuals (RQR), mean–variance comparisons, residual autocorrelation, and lag-kernel behaviour. Unless stated otherwise, diagnostics are shown per pathogen.

Table G.1: Information-criterion comparison for lag-basis specifications.

| Model tag | **AIC** | **BIC** |
|---|---|---|
| deg1_df2 | 22503 | 23981 |
| deg1_df3 | 22573 | 24345 |
| deg1_df4 | 22672 | 24740 |
| deg3_df4 | 22704 | 24772 |
| deg1_df5 | 22792 | 25156 |
| deg3_df5 | 23001 | 25365 |

#### Distributional fit

QQ plots of residuals (Figure G.1) showed close adherence to the reference line for most pathogens, indicating that the fitted distributions captured the central quantiles well. Deviations were primarily limited to the tails. The corresponding histograms (Figure G.2) were approximately symmetric and centered near zero.

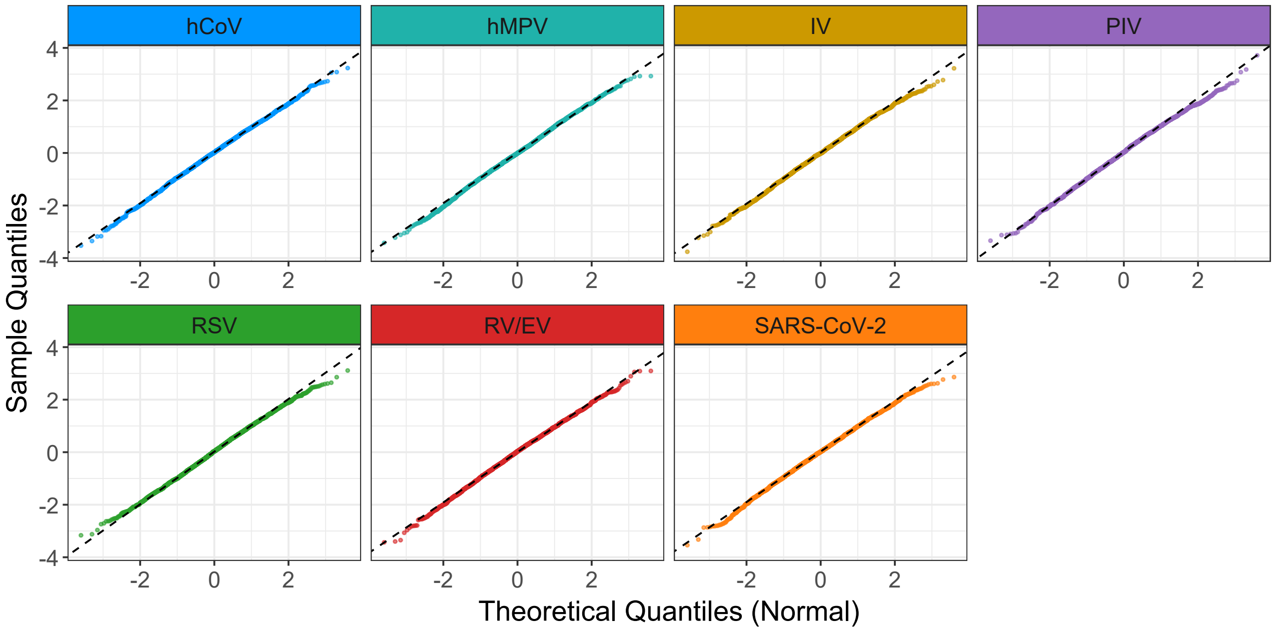


Figure G.1: Disease-specific Q–Q plots of randomized quantile residuals under the DLNM specification.

#### Residuals versus fitted means

Residuals versus fitted means (Figure G.3) were broadly well-centered around zero, consistent with appropriate calibration of the conditional mean. Slight systematic deviations were visible for RSV, PIV, and hMPV, where residuals were more positive at low fitted values and trended negative at higher means. In contrast, residuals for RV/EV, SARS-CoV-2, and hCoV formed tight, symmetric bands across the fitted range. Some heteroskedasticity was evident in IV, but no strong patterns indicative of model misspecification were observed.

#### Mean–variance adequacy

Variance–mean plots (Figure G.4) followed the negative binomial expectation closely, with empirical variance tracking the fitted mean across all pathogens. Minor deviations were observed at higher fitted values for RSV, PIV, and hMPV, but no consistent overdispersion or heteroskedasticity was evident. These patterns suggest that the DLNM formulation captured the dispersion structure adequately across diverse incidence ranges.

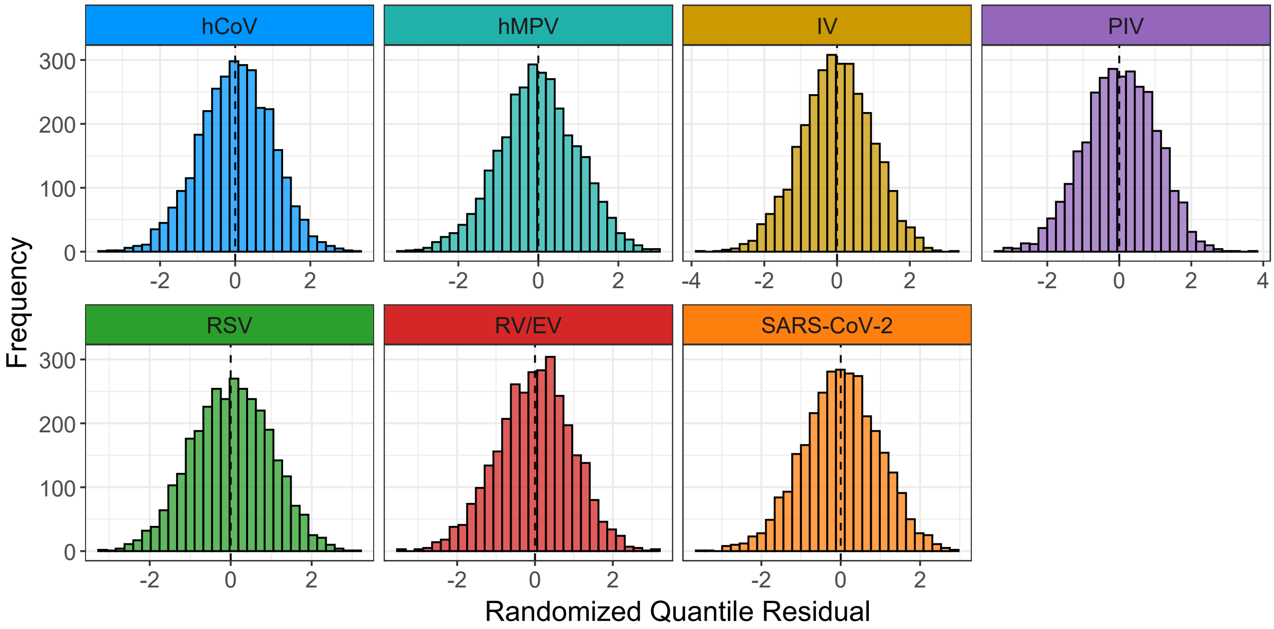


Figure G.2: Histograms of randomized quantile residuals by pathogen for the DLNM.

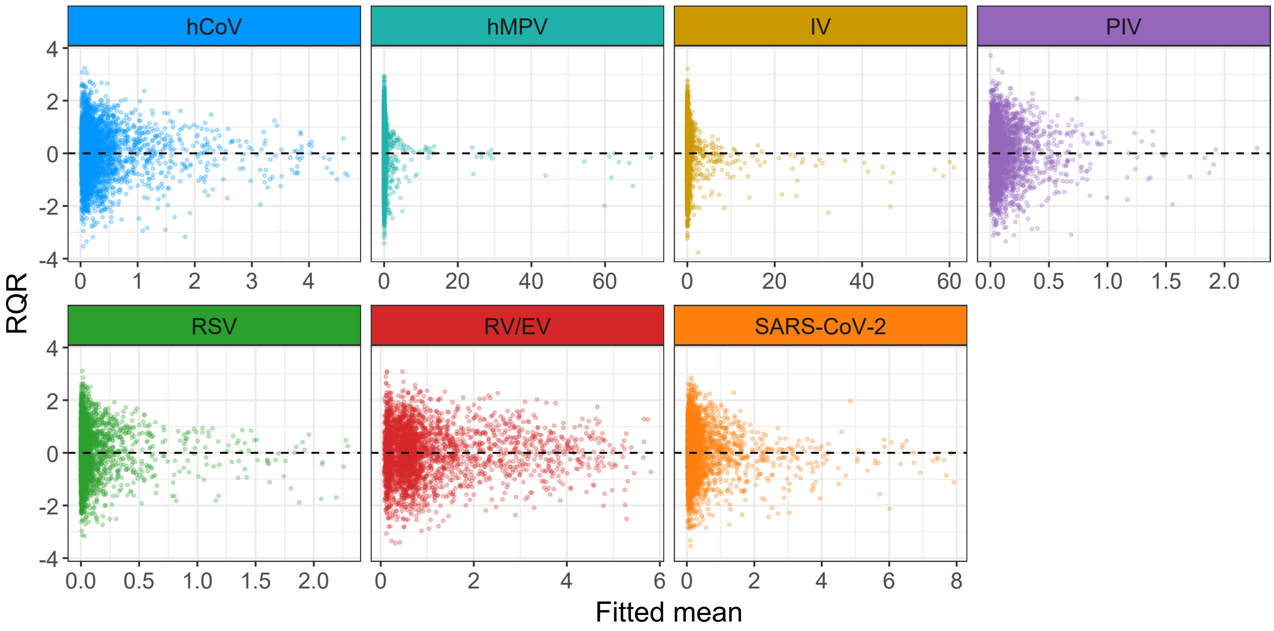


Figure G.3: Randomized quantile residuals versus fitted means by pathogen under the DLNM.

##### Residual autocorrelation

Autocorrelation functions of residuals (Figure G.5) showed minimal temporal dependence beyond lag 1 across all pathogens. Residuals were largely uncorrelated at higher lags, indicating that the distributed-lag and seasonal spline terms sufficiently accounted for temporal structure in the data. No substantial residual autocorrelation was observed, supporting the adequacy of the model's dynamic components.

#### G.4 Additional Results

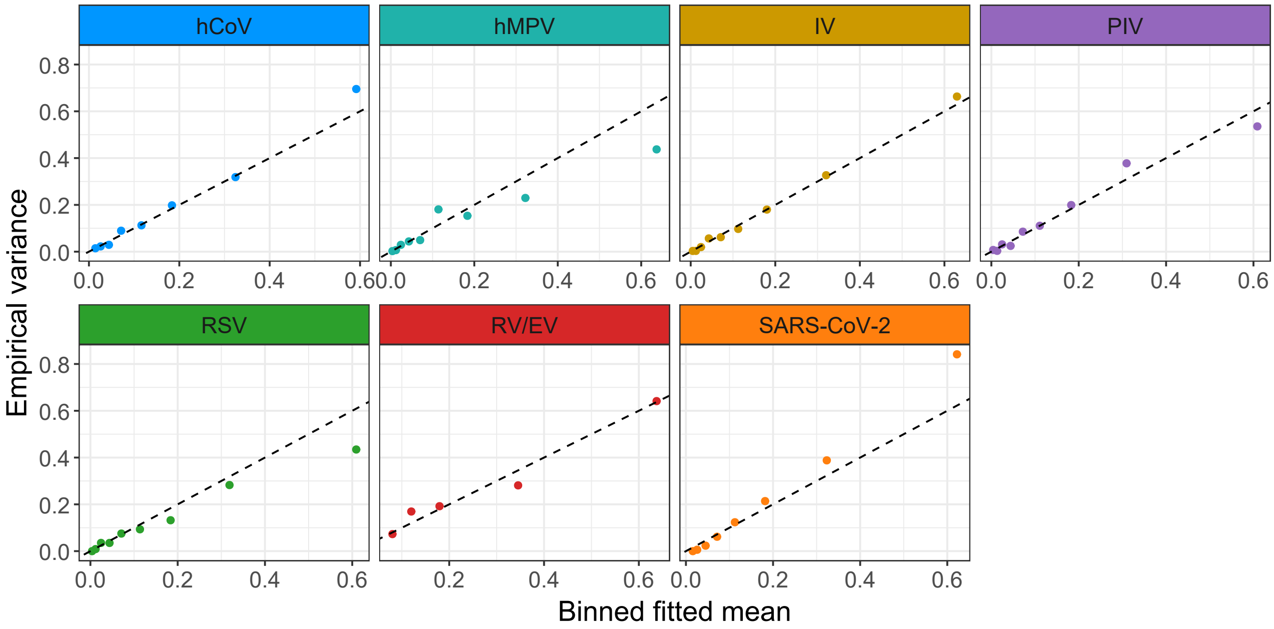


Figure G.4: Binned fitted mean ($\hat{\mu}$) versus empirical variance by pathogen for the DLNM. The dashed line indicates the negative binomial variance expectation.

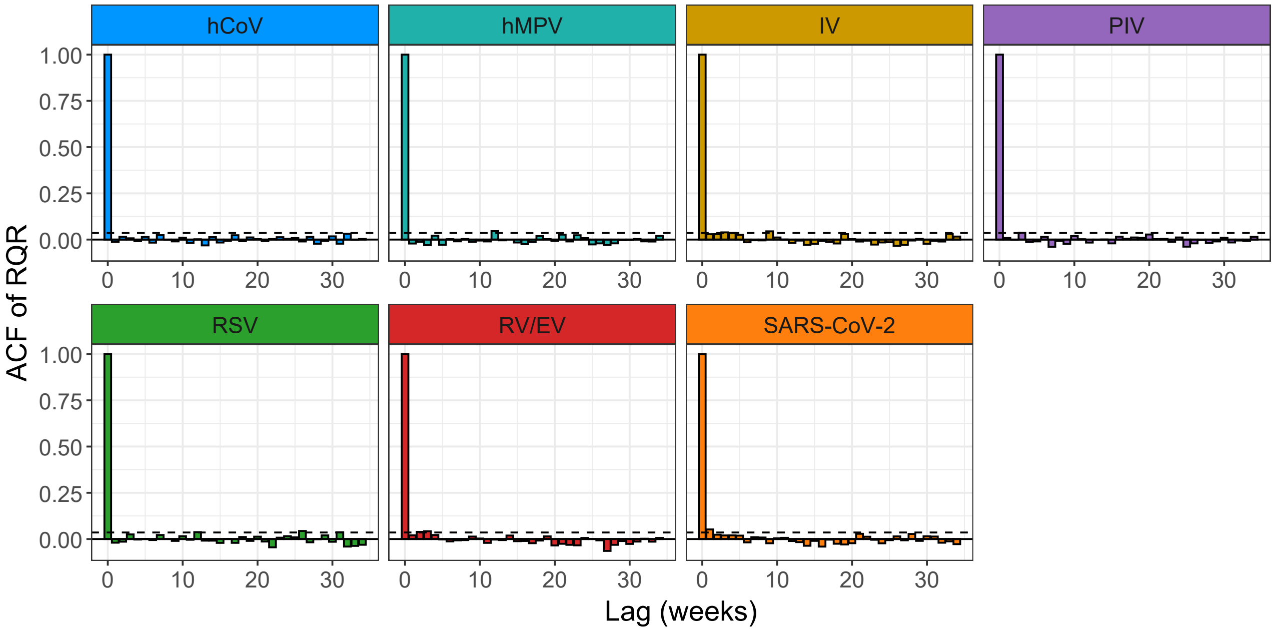


Figure G.5: Autocorrelation functions of randomized quantile residuals by pathogen for the DLNM. Dashed lines mark approximate $\pm 1.96/\sqrt{n}$ bounds.

Table G.2: RR of the DLNM across all pathogen outcomes with 95% CI for a predictor value of 1.

| | | Target | | | | | | |
|---|---|---|---|---|---|---|---|---|
| | **Lag** | **hMPV** | **IV** | **PIV** | **RV/EV** | **RSV** | **SARS-COV-2** | **SCOV** |
| **hMPV** | **1** | 1.1370 [1.0880; 1.1883] | 0.9592 [0.9210; 0.9989] | 1.1117 [1.0749; 1.1498] | 0.9922 [0.9796; 1.0049] | 1.0302 [0.9818; 1.0810] | 0.9673 [0.9433; 0.9918] | 0.9924 [0.9719; 1.0133] |
| | **2** | 1.2928 [1.1837; 1.4120] | 0.9201 [0.8483; 0.9979] | 1.2359 [1.1554; 1.3221] | 0.9844 [0.9596; 1.0099] | 1.0613 [0.9639; 1.1686] | 0.9356 [0.8898; 0.9837] | 0.9849 [0.9447; 1.0268] |
| | **3** | 1.4699 [1.2878; 1.6778] | 0.8825 [0.7813; 0.9968] | 1.3740 [1.2420; 1.5202] | 0.9767 [0.9400; 1.0149] | 1.0934 [0.9463; 1.2633] | 0.9049 [0.8393; 0.9757] | 0.9774 [0.9182; 1.0405] |
| | **4** | 1.6714 [1.4011; 1.9937] | 0.8465 [0.7196; 0.9958] | 1.5276 [1.3350; 1.7479] | 0.9691 [0.9208; 1.0199] | 1.1264 [0.9291; 1.3657] | 0.8753 [0.7917; 0.9678] | 0.9700 [0.8924; 1.0543] |

*Continued on next page*

Table G.2: (continued)

| Predictor | Lag | hMPV | IV | PIV | RV/EV | RSV | SARS-COV-2 | SCOV |
|---|---|---|---|---|---|---|---|---|
| | 5 | 1.3972 [1.2335; 1.5825] | 0.8484 [0.7562; 0.9519] | 1.3685 [1.2477; 1.5011] | 0.9980 [0.9636; 1.0336] | 1.0358 [0.9023; 1.1889] | 0.9453 [0.8799; 1.0155] | 0.9798 [0.9247; 1.0382] |
| | 6 | 1.1679 [1.0325; 1.3211] | 0.8504 [0.7607; 0.9507] | 1.2261 [1.1185; 1.3440] | 1.0277 [0.9946; 1.0619] | 0.9524 [0.8228; 1.1025] | 1.0209 [0.9542; 1.0922] | 0.9897 [0.9368; 1.0456] |
| | 7 | 0.9763 [0.8208; 1.1614] | 0.8523 [0.7301; 0.9951] | 1.0985 [0.9613; 1.2552] | 1.0583 [1.0106; 1.1083] | 0.8758 [0.7096; 1.0808] | 1.1025 [1.0063; 1.2079] | 0.9997 [0.9255; 1.0799] |
| | 8 | 0.8162 [0.6384; 1.0434] | 0.8543 [0.6864; 1.0632] | 0.9841 [0.8127; 1.1917] | 1.0898 [1.0197; 1.1648] | 0.8053 [0.5990; 1.0826] | 1.1906 [1.0472; 1.3537] | 1.0098 [0.9044; 1.1275] |
| IV | 1 | 1.0190 [0.9932; 1.0454] | 1.0757 [1.0550; 1.0969] | 0.9716 [0.9484; 0.9953] | 0.9915 [0.9839; 0.9991] | 0.9947 [0.9704; 1.0196] | 1.0013 [0.9864; 1.0164] | 0.9915 [0.9802; 1.0030] |
| | 2 | 1.0383 [0.9865; 1.0928] | 1.1572 [1.1130; 1.2032] | 0.9439 [0.8994; 0.9907] | 0.9830 [0.9680; 0.9982] | 0.9895 [0.9417; 1.0396] | 1.0026 [0.9730; 1.0331] | 0.9831 [0.9607; 1.0060] |
| | 3 | 1.0579 [0.9798; 1.1423] | 1.2448 [1.1742; 1.3197] | 0.9171 [0.8529; 0.9860] | 0.9746 [0.9524; 0.9974] | 0.9842 [0.9139; 1.0600] | 1.0039 [0.9597; 1.0501] | 0.9748 [0.9417; 1.0091] |
| | 4 | 1.0780 [0.9731; 1.1942] | 1.3391 [1.2387; 1.4476] | 0.8910 [0.8089; 0.9814] | 0.9663 [0.9370; 0.9965] | 0.9790 [0.8869; 1.0808] | 1.0052 [0.9467; 1.0673] | 0.9665 [0.9230; 1.0121] |
| | 5 | 1.1633 [1.0831; 1.2496] | 1.1603 [1.1011; 1.2226] | 0.9118 [0.8539; 0.9738] | 0.9956 [0.9751; 1.0165] | 0.9944 [0.9301; 1.0631] | 1.0375 [0.9958; 1.0809] | 0.9835 [0.9529; 1.0150] |
| | 6 | 1.2554 [1.1750; 1.3414] | 1.0053 [0.9529; 1.0606] | 0.9332 [0.8822; 0.9872] | 1.0258 [1.0072; 1.0447] | 1.0100 [0.9453; 1.0791] | 1.0709 [1.0304; 1.1129] | 1.0007 [0.9717; 1.0306] |
| | 7 | 1.3548 [1.2369; 1.4840] | 0.8710 [0.8039; 0.9438] | 0.9551 [0.8846; 1.0311] | 1.0569 [1.0302; 1.0842] | 1.0258 [0.9304; 1.1310] | 1.1053 [1.0464; 1.1675] | 1.0183 [0.9767; 1.0616] |
| | 8 | 1.4621 [1.2841; 1.6646] | 0.7547 [0.6720; 0.8476] | 0.9774 [0.8742; 1.0929] | 1.0889 [1.0491; 1.1302] | 1.0419 [0.9047; 1.1999] | 1.1408 [1.0543; 1.2345] | 1.0361 [0.9757; 1.1003] |
| PIV | 1 | 1.0363 [0.9773; 1.0989] | 1.0224 [0.9690; 1.0788] | 0.9732 [0.9373; 1.0103] | 1.0158 [1.0018; 1.0300] | 1.0003 [0.9402; 1.0642] | 0.9572 [0.9278; 0.9874] | 0.9798 [0.9545; 1.0059] |
| | 2 | 1.0739 [0.9551; 1.2076] | 1.0453 [0.9390; 1.1638] | 0.9470 [0.8786; 1.0208] | 1.0319 [1.0036; 1.0609] | 1.0005 [0.8840; 1.1325] | 0.9161 [0.8609; 0.9749] | 0.9601 [0.9111; 1.0118] |
| | 3 | 1.1129 [0.9333; 1.3271] | 1.0688 [0.9098; 1.2554] | 0.9216 [0.8236; 1.0313] | 1.0482 [1.0054; 1.0927] | 1.0008 [0.8311; 1.2052] | 0.8769 [0.7988; 0.9626] | 0.9408 [0.8696; 1.0177] |
| | 4 | 1.1534 [0.9121; 1.4584] | 1.0927 [0.8816; 1.3543] | 0.8969 [0.7720; 1.0419] | 1.0647 [1.0072; 1.1255] | 1.0011 [0.7814; 1.2825] | 0.8393 [0.7412; 0.9505] | 0.9218 [0.8300; 1.0237] |
| | 5 | 1.0962 [0.9238; 1.3006] | 1.0738 [0.9170; 1.2573] | 0.8903 [0.7983; 0.9929] | 1.0616 [1.0194; 1.1055] | 0.9833 [0.8208; 1.1778] | 0.8750 [0.7999; 0.9572] | 0.9403 [0.8711; 1.0150] |
| | 6 | 1.0418 [0.8929; 1.2155] | 1.0551 [0.9169; 1.2142] | 0.8838 [0.8009; 0.9754] | 1.0584 [1.0211; 1.0970] | 0.9658 [0.8259; 1.1294] | 0.9122 [0.8444; 0.9854] | 0.9592 [0.8960; 1.0269] |
| | 7 | 0.9901 [0.8135; 1.2052] | 1.0368 [0.8706; 1.2348] | 0.8774 [0.7733; 0.9954] | 1.0552 [1.0090; 1.1036] | 0.9486 [0.7825; 1.1500] | 0.9510 [0.8641; 1.0466] | 0.9785 [0.8978; 1.0665] |
| | 8 | 0.9411 [0.7172; 1.2349] | 1.0189 [0.8020; 1.2944] | 0.8710 [0.7313; 1.0374] | 1.0521 [0.9890; 1.1191] | 0.9317 [0.7147; 1.2146] | 0.9914 [0.8679; 1.1325] | 0.9982 [0.8863; 1.1242] |
| RV/EV | 1 | 1.0077 [0.9908; 1.0249] | 0.9777 [0.9651; 0.9905] | 1.0115 [1.0010; 1.0221] | 1.0086 [1.0051; 1.0121] | 1.0016 [0.9884; 1.0150] | 1.0102 [1.0038; 1.0168] | 0.9920 [0.9851; 0.9989] |
| | 2 | 1.0156 [0.9818; 1.0505] | 0.9560 [0.9315; 0.9811] | 1.0231 [1.0020; 1.0447] | 1.0172 [1.0102; 1.0244] | 1.0032 [0.9769; 1.0302] | 1.0206 [1.0075; 1.0339] | 0.9841 [0.9705; 0.9979] |
| | 3 | 1.0234 [0.9728; 1.0767] | 0.9347 [0.8990; 0.9718] | 1.0349 [1.0030; 1.0678] | 1.0260 [1.0153; 1.0368] | 1.0048 [0.9655; 1.0457] | 1.0311 [1.0113; 1.0512] | 0.9762 [0.9560; 0.9968] |
| | 4 | 1.0313 [0.9639; 1.1035] | 0.9138 [0.8676; 0.9625] | 1.0468 [1.0041; 1.0914] | 1.0348 [1.0204; 1.0493] | 1.0064 [0.9543; 1.0614] | 1.0416 [1.0151; 1.0689] | 0.9684 [0.9418; 0.9957] |
| | 5 | 1.0144 [0.9682; 1.0629] | 0.9561 [0.9238; 0.9895] | 1.0199 [0.9921; 1.0485] | 1.0165 [1.0071; 1.0260] | 1.0332 [0.9966; 1.0712] | 1.0333 [1.0157; 1.0511] | 0.9750 [0.9569; 0.9934] |
| | 6 | 0.9978 [0.9549; 1.0426] | 1.0003 [0.9706; 1.0309] | 0.9937 [0.9668; 1.0213] | 0.9986 [0.9897; 1.0075] | 1.0607 [1.0265; 1.0961] | 1.0249 [1.0083; 1.0419] | 0.9816 [0.9647; 0.9987] |
| | 7 | 0.9815 [0.9226; 1.0441] | 1.0465 [1.0021; 1.0929] | 0.9682 [0.9290; 1.0089] | 0.9809 [0.9680; 0.9940] | 1.0889 [1.0396; 1.1407] | 1.0167 [0.9924; 1.0416] | 0.9882 [0.9638; 1.0133] |
| | 8 | 0.9654 [0.8834; 1.0550] | 1.0949 [1.0270; 1.1672] | 0.9433 [0.8883; 1.0017] | 0.9636 [0.9451; 0.9825] | 1.1179 [1.0452; 1.1957] | 1.0085 [0.9736; 1.0447] | 0.9949 [0.9593; 1.0318] |
| RSV | 1 | 0.9434 [0.8850; 1.0057] | 1.1502 [1.1033; 1.1992] | 0.9742 [0.9176; 1.0342] | 0.9675 [0.9505; 0.9848] | 0.9642 [0.9178; 1.0129] | 1.0157 [0.9870; 1.0452] | 1.0024 [0.9758; 1.0297] |
| | 2 | 0.8900 [0.7831; 1.0114] | 1.3230 [1.2172; 1.4381] | 0.9490 [0.8420; 1.0696] | 0.9361 [0.9035; 0.9699] | 0.9296 [0.8424; 1.0260] | 1.0317 [0.9743; 1.0924] | 1.0047 [0.9521; 1.0602] |
| | 3 | 0.8396 [0.6931; 1.0171] | 1.5218 [1.3428; 1.7245] | 0.9245 [0.7727; 1.1062] | 0.9057 [0.8588; 0.9552] | 0.8963 [0.7731; 1.0392] | 1.0479 [0.9616; 1.1418] | 1.0071 [0.9291; 1.0916] |
| | 4 | 0.7920 [0.6133; 1.0228] | 1.7504 [1.4815; 2.0681] | 0.9006 [0.7090; 1.1440] | 0.8763 [0.8163; 0.9407] | 0.8642 [0.7096; 1.0526] | 1.0643 [0.9492; 1.1934] | 1.0094 [0.9066; 1.1240] |
| | 5 | 0.7802 [0.6460; 0.9421] | 1.6583 [1.4633; 1.8792] | 0.8822 [0.7443; 1.0456] | 0.8985 [0.8533; 0.9461] | 0.8639 [0.7432; 1.0044] | 0.9985 [0.9173; 1.0869] | 1.0355 [0.9570; 1.1204] |
| | 6 | 0.7685 [0.6429; 0.9185] | 1.5710 [1.3957; 1.7683] | 0.8641 [0.7282; 1.0254] | 0.9212 [0.8774; 0.9673] | 0.8637 [0.7412; 1.0063] | 0.9368 [0.8621; 1.0179] | 1.0622 [0.9877; 1.1424] |
| | 7 | 0.7569 [0.5998; 0.9552] | 1.4883 [1.2795; 1.7313] | 0.8464 [0.6647; 1.0779] | 0.9446 [0.8855; 1.0076] | 0.8634 [0.7052; 1.0571] | 0.8789 [0.7869; 0.9816] | 1.0896 [0.9920; 1.1969] |
| | 8 | 0.7456 [0.5413; 1.0270] | 1.4100 [1.1481; 1.7317] | 0.8291 [0.5895; 1.1661] | 0.9685 [0.8855; 1.0593] | 0.8631 [0.6558; 1.1359] | 0.8246 [0.7081; 0.9601] | 1.1178 [0.9820; 1.2723] |
| ARS-COV-2 | 1 | 1.0000 [0.9788; 1.0217] | 0.9990 [0.9842; 1.0139] | 1.0094 [0.9923; 1.0267] | 1.0014 [0.9959; 1.0069] | 1.0239 [1.0062; 1.0418] | 1.0291 [1.0204; 1.0379] | 1.0012 [0.9923; 1.0101] |
| | 2 | 1.0001 | 0.9979 | 1.0188 | 1.0027 | 1.0483 | 1.0591 | 1.0024 |

*Continued on next page*

Table G.2: (continued)

| Predictor | Lag | hMPV | IV | PIV | RV/EV | RSV | SARS-COV-2 | SCOV |
|---|---|---|---|---|---|---|---|---|
| | | [0.9581; 1.0439] | [0.9687; 1.0281] | [0.9848; 1.0540] | [0.9917; 1.0139] | [1.0125; 1.0853] | [1.0413; 1.0772] | [0.9847; 1.0204] |
| | **3** | 1.0001 | 0.9969 | 1.0283 | 1.0041 | 1.0733 | 1.0899 | 1.0036 |
| | | [0.9378; 1.0666] | [0.9534; 1.0424] | [0.9772; 1.0821] | [0.9876; 1.0209] | [1.0189; 1.1307] | [1.0626; 1.1180] | [0.9772; 1.0307] |
| | **4** | 1.0002 | 0.9959 | 1.0380 | 1.0055 | 1.0989 | 1.1216 | 1.0048 |
| | | [0.9180; 1.0898] | [0.9383; 1.0569] | [0.9697; 1.1110] | [0.9835; 1.0280] | [1.0252; 1.1779] | [1.0843; 1.1603] | [0.9697; 1.0412] |
| | **5** | 1.0153 | 1.0217 | 1.0281 | 1.0003 | 1.1234 | 1.0740 | 1.0202 |
| | | [0.9600; 1.0737] | [0.9819; 1.0631] | [0.9822; 1.0761] | [0.9860; 1.0148] | [1.0710; 1.1785] | [1.0501; 1.0985] | [0.9965; 1.0444] |
| | **6** | 1.0306 | 1.0482 | 1.0182 | 0.9952 | 1.1485 | 1.0284 | 1.0358 |
| | | [0.9791; 1.0847] | [1.0085; 1.0894] | [0.9764; 1.0619] | [0.9821; 1.0084] | [1.0997; 1.1995] | [1.0057; 1.0517] | [1.0145; 1.0576] |
| | **7** | 1.0461 | 1.0754 | 1.0085 | 0.9900 | 1.1741 | 0.9848 | 1.0517 |
| | | [0.9693; 1.1290] | [1.0156; 1.1387] | [0.9494; 1.0713] | [0.9708; 1.0097] | [1.1057; 1.2468] | [0.9522; 1.0185] | [1.0205; 1.0838] |
| | **8** | 1.0619 | 1.1033 | 0.9989 | 0.9849 | 1.2003 | 0.9430 | 1.0678 |
| | | [0.9486; 1.1886] | [1.0151; 1.1991] | [0.9147; 1.0908] | [0.9567; 1.0140] | [1.1010; 1.3086] | [0.8979; 0.9904] | [1.0214; 1.1163] |
| **SCOV** | **1** | 1.0619 | 1.0670 | 1.0194 | 0.9965 | 1.0021 | 1.0038 | 1.0248 |
| | | [1.0365; 1.0880] | [1.0462; 1.0882] | [1.0008; 1.0383] | [0.9897; 1.0032] | [0.9758; 1.0291] | [0.9905; 1.0174] | [1.0140; 1.0358] |
| | **2** | 1.1277 | 1.1385 | 1.0392 | 0.9929 | 1.0042 | 1.0077 | 1.0503 |
| | | [1.0743; 1.1837] | [1.0945; 1.1843] | [1.0016; 1.0781] | [0.9796; 1.0064] | [0.9521; 1.0591] | [0.9811; 1.0350] | [1.0282; 1.0728] |
| | **3** | 1.1975 | 1.2148 | 1.0593 | 0.9894 | 1.0063 | 1.0116 | 1.0764 |
| | | [1.1136; 1.2878] | [1.1450; 1.2888] | [1.0024; 1.1195] | [0.9696; 1.0096] | [0.9290; 1.0900] | [0.9718; 1.0530] | [1.0426; 1.1112] |
| | **4** | 1.2717 | 1.2962 | 1.0799 | 0.9859 | 1.0084 | 1.0155 | 1.1031 |
| | | [1.1542; 1.4011] | [1.1979; 1.4025] | [1.0032; 1.1624] | [0.9596; 1.0129] | [0.9065; 1.1217] | [0.9625; 1.0713] | [1.0573; 1.1510] |
| | **5** | 1.1428 | 1.1473 | 1.0769 | 0.9918 | 0.9953 | 0.9891 | 1.0681 |
| | | [1.0674; 1.2235] | [1.0854; 1.2127] | [1.0215; 1.1353] | [0.9732; 1.0109] | [0.9227; 1.0736] | [0.9522; 1.0275] | [1.0364; 1.1008] |
| | **6** | 1.0269 | 1.0155 | 1.0740 | 0.9978 | 0.9824 | 0.9635 | 1.0343 |
| | | [0.9611; 1.0973] | [0.9587; 1.0756] | [1.0216; 1.1290] | [0.9798; 1.0161] | [0.9099; 1.0608] | [0.9277; 1.0007] | [1.0042; 1.0652] |
| | **7** | 0.9229 | 0.8988 | 1.0710 | 1.0038 | 0.9697 | 0.9385 | 1.0015 |
| | | [0.8412; 1.0124] | [0.8271; 0.9767] | [1.0012; 1.1457] | [0.9788; 1.0295] | [0.8699; 1.0810] | [0.8899; 0.9898] | [0.9612; 1.0435] |
| | **8** | 0.8293 | 0.7956 | 1.0681 | 1.0099 | 0.9572 | 0.9142 | 0.9697 |
| | | [0.7273; 0.9456] | [0.7071; 0.8951] | [0.9718; 1.1740] | [0.9743; 1.0467] | [0.8212; 1.1157] | [0.8481; 0.9855] | [0.9151; 1.0276] |

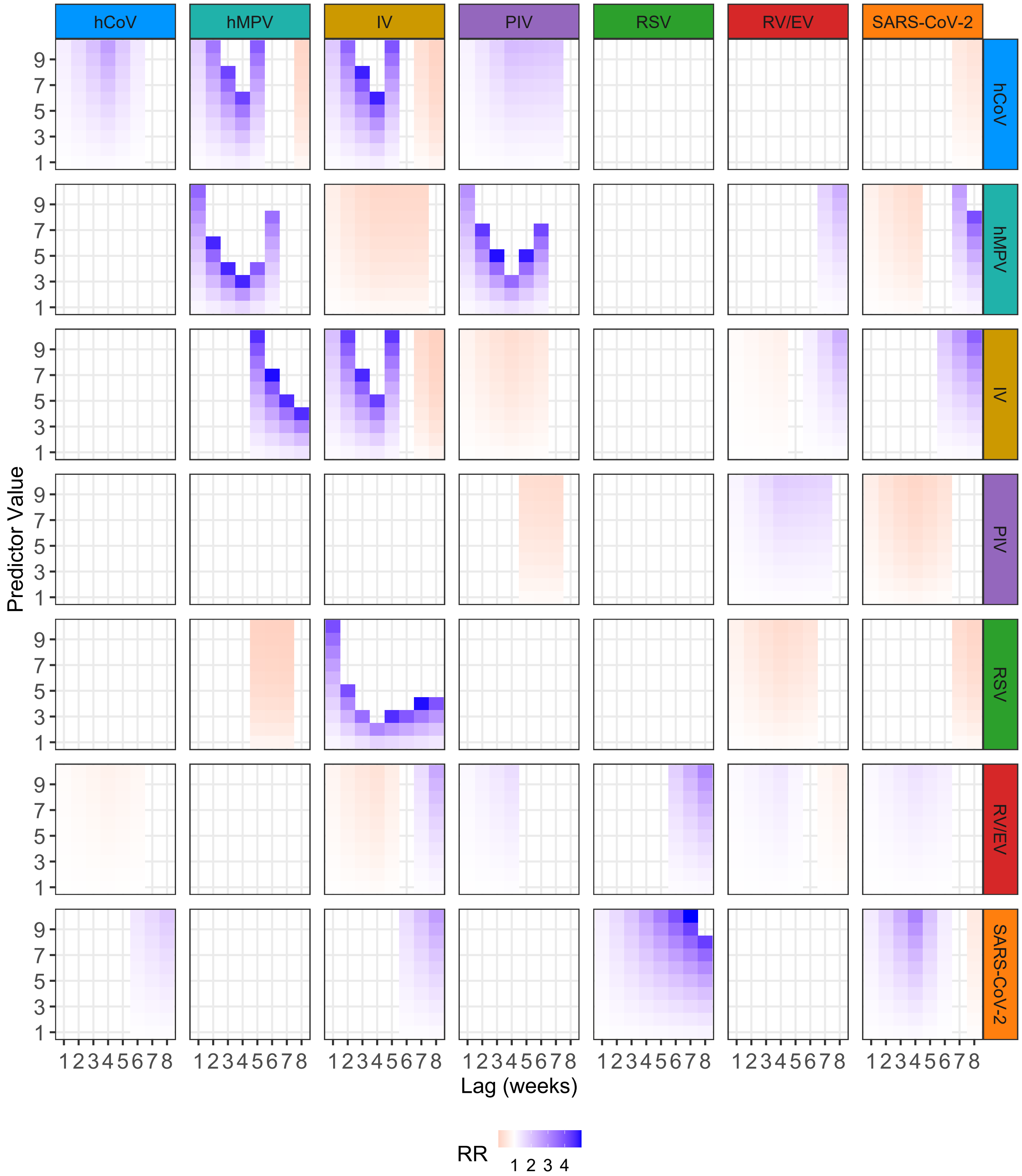


Figure G.6: Heatmap of relative risks (RRs) from the DLNM. Rows indicate the target disease and columns indicate the predictor disease. Each cell shows RR across lag (x-axis, weeks) and predictor value (y-axis, representing the observed log-transformed or standardised counts of the predictor pathogen). Blue indicates higher risk (RR > 1), and orange/red indicates lower risk (RR < 1). For example, a blue area at higher predictor values and at lag 6 implies that weeks with unusually high incidence of the predictor pathogen are associated with increased risk of the target pathogen six weeks later.

Only statistically significant effects (95% CI not including 1) are shown.

#### G.5 Sensitivity Analysis: Different specification of the functional form of the crossbasis

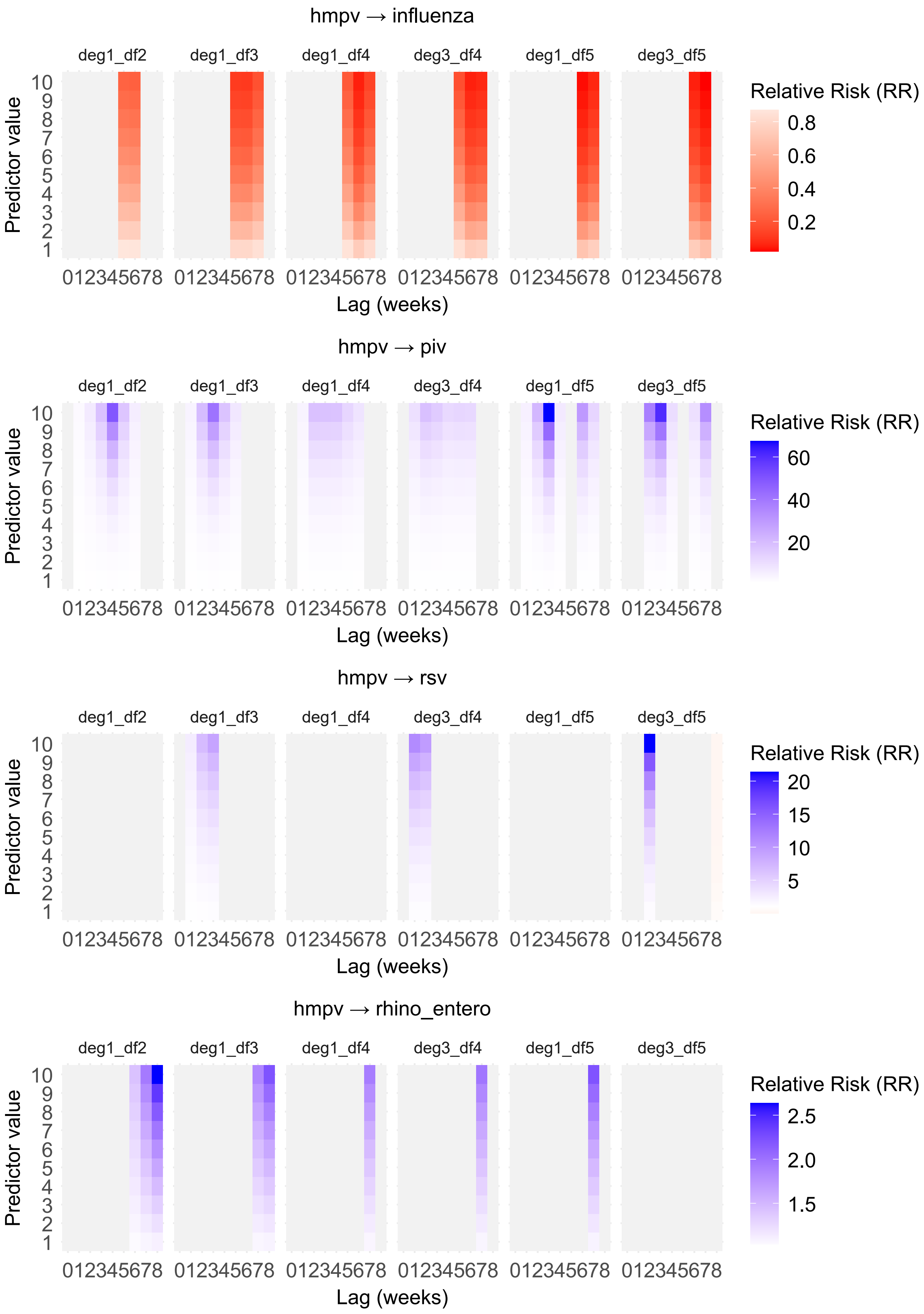


Figure G.7: Caption

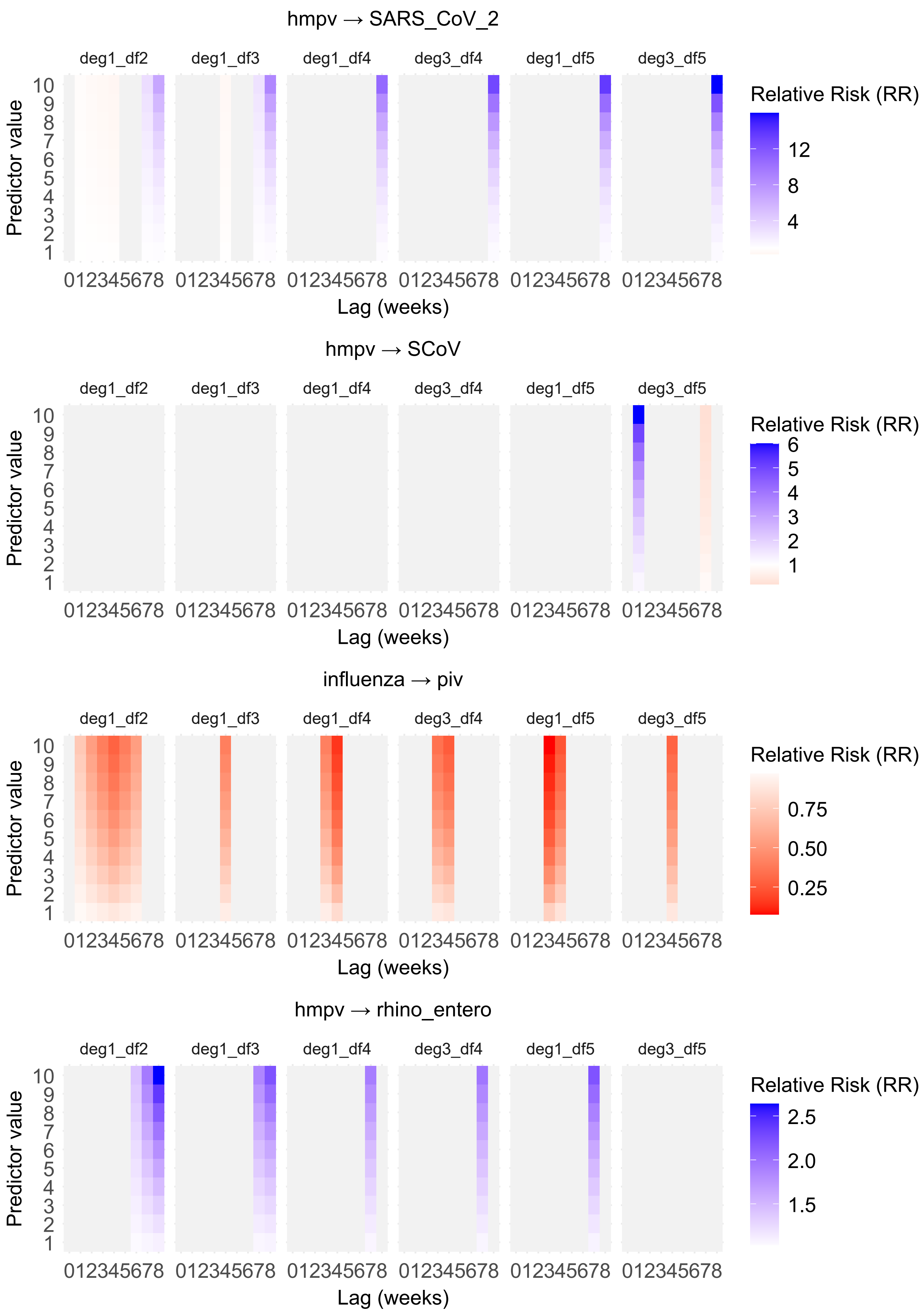


Figure G.8: Caption

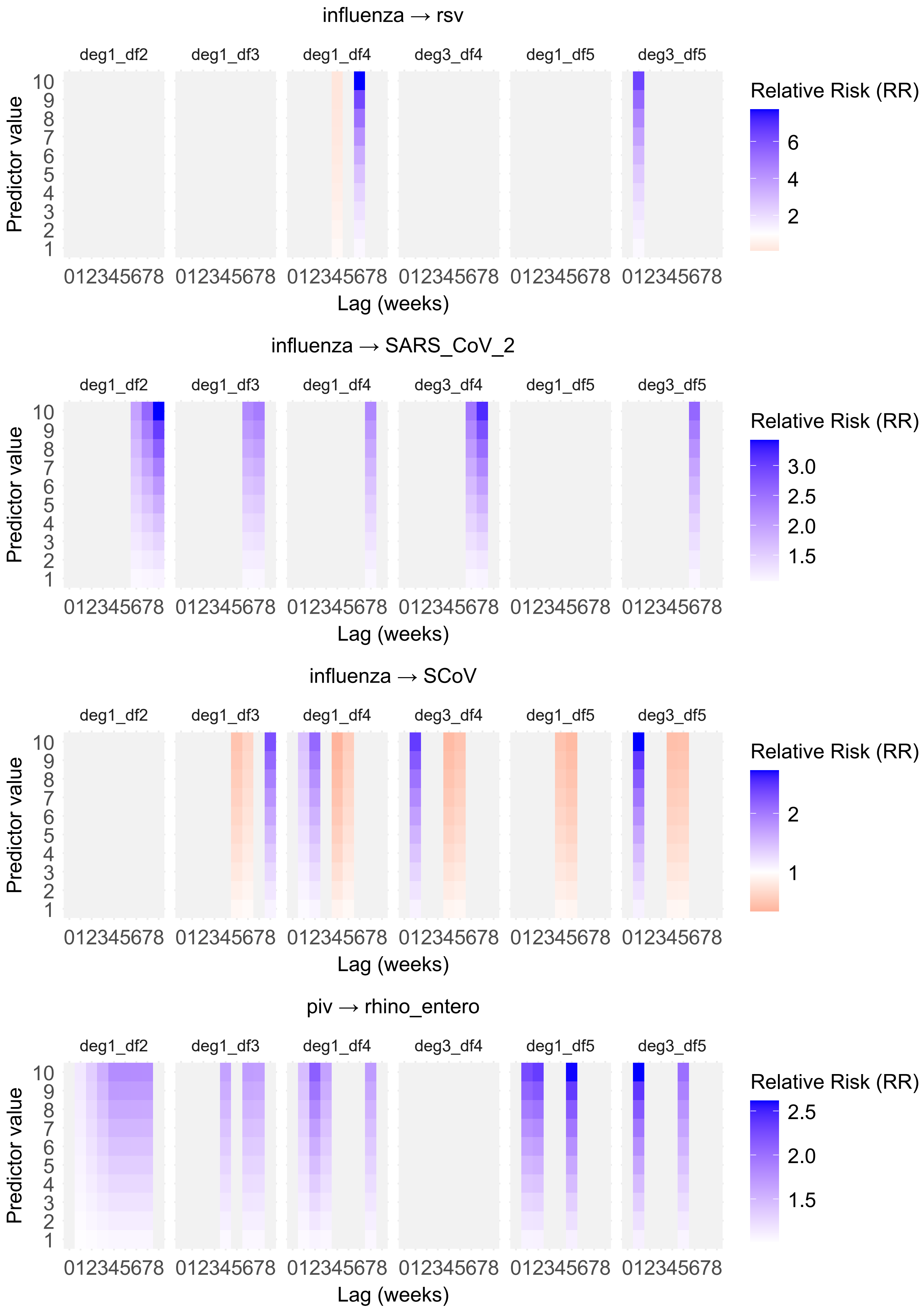


Figure G.9: Caption

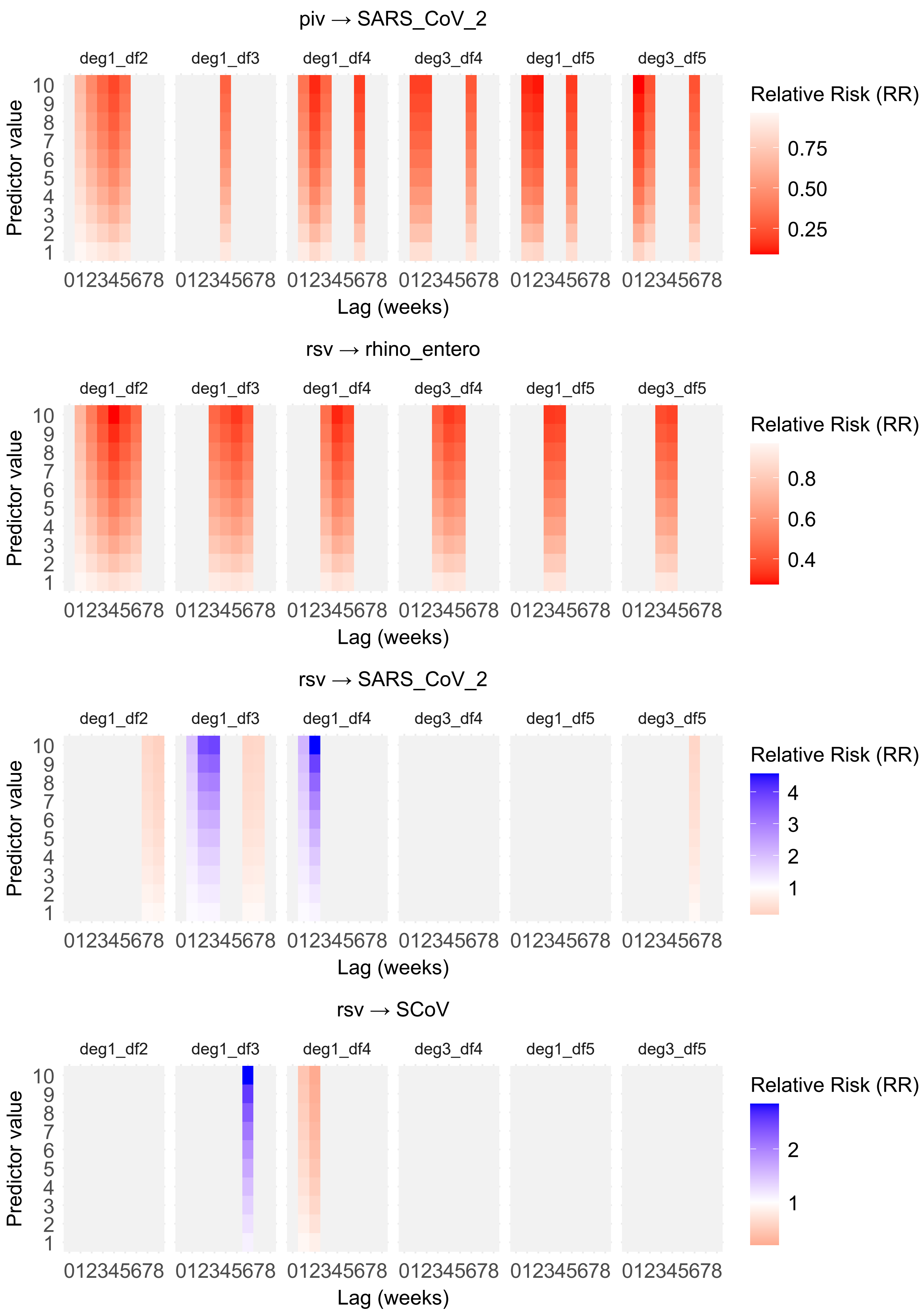


Figure G.10: Caption

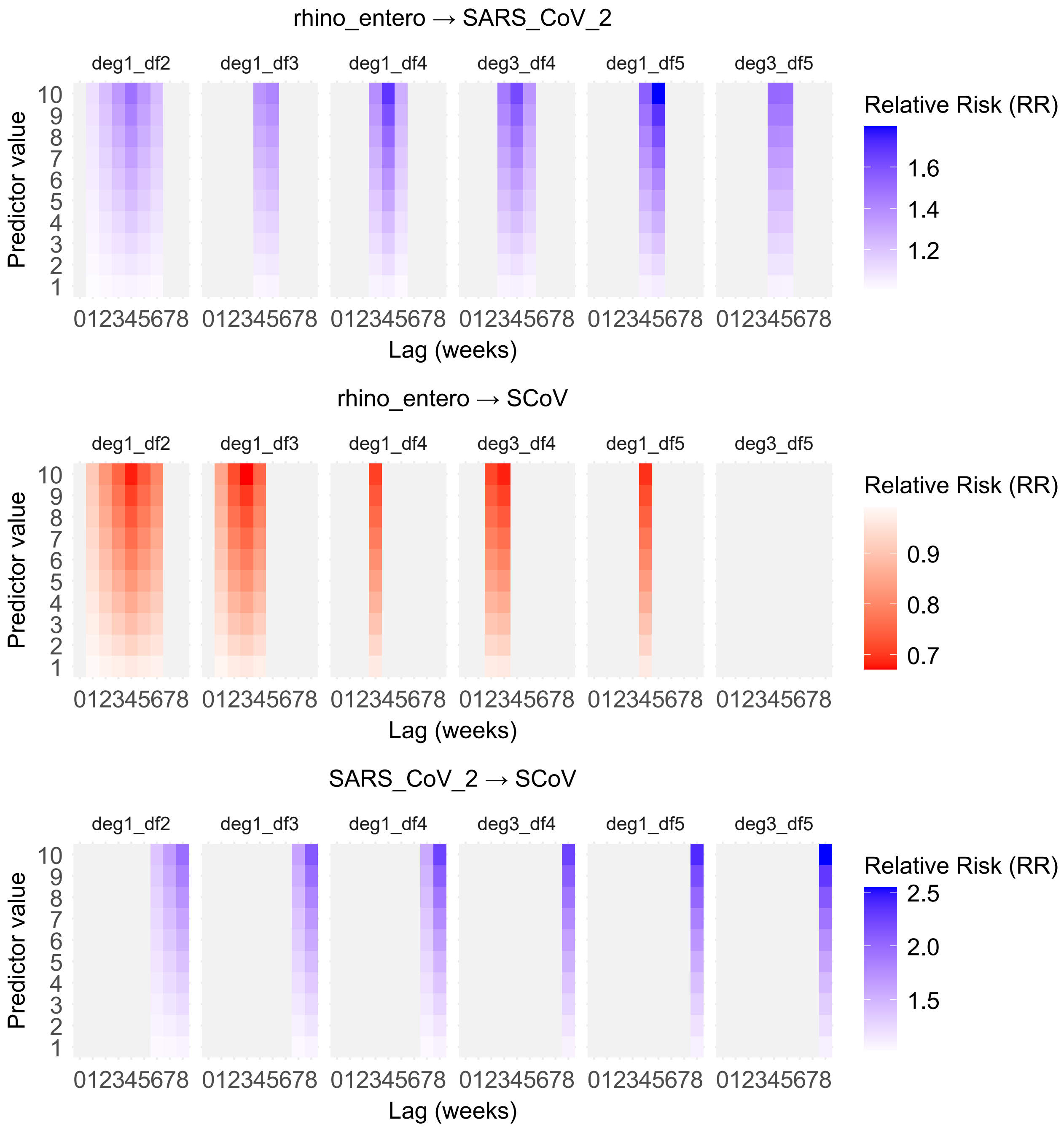


Figure G.11: Caption

### H Comparative Results

Table H.1: Summary of cross-pathogen associations across methods

| | Contemporaneous | Short-term | | | | | Long-term | Exploratory | | |
|---|---|---|---|---|---|---|---|---|---|---|
| | LGM | Regression | | | E&E | | DLNM | PCA | Granger | |
| | | GLMM | GAMLSS | VGLM | epidemic | endemic | | cosine similarity | National | Regional |
| hMPV→IV | 0.40 | | | | | | − (lags 1 − 7) | | * | 22% |
| IV→hMPV | | | | | | + | + (lags 5 − 8) | | * | 22% |
| hMPV→PIV | | + | + | + | | + | + (lags 1 − 6) | 0.636 | | 38% |
| PIV→hMPV | | | | + | | + | | | | |
| hMPV→RV/EV | | | | | | + | + (lags 7 − 8) | | | |
| RV/EV→hMPV | | | | | | + | | | | |
| hMPV→RSV | 0.21 | | | + | | + | | | | |
| RSV→hMPV | | | | + | | + | − (lags 5 − 7) | | | |
| hMPV→SARS-CoV-2 | | | | | | | −/+ (lags 1 − 4/7 − 8) | | | |
| SARS-CoV-2→hMPV | | | | + | | + | | | | |
| hMPV→hCoV | 0.41 | | | + | | + | | 0.921 | | |
| hCoV→hMPV | | | | + | + | | +/− (lags 1 − 5/8) | | | |
| IV→PIV | −0.46 | | - | - | | + | − (lags 1 − 6) | −0.625 | | 20% |
| PIV→IV | | | | | | | | | | |
| IV→RV/EV | −0.21 | - | - | - | | | −/+ (lags 1 − 4/6 − 8) | −0.579 | | |
| RV/EV→IV | | | | | | | −/+ (lags 1 − 5/7 − 8) | | | |
| IV→RSV | 0.51 | | | | | + | | | | 38% |
| RSV→IV | | | | | | + | + (lags 1 − 8) | | | 50% |
| IV→SARS-CoV-2 | | | | | | + | + (lags 6 − 8) | | | |
| SARS-CoV-2→IV | | + | + | | | | + (lags 6 − 8) | | | |
| IV→hCoV | 0.44 | | | + | | + | | 0.479 | | |
| hCoV→IV | | + | + | + | + | + | +/− (lags 1 − 6/7 − 8) | | | |
| PIV→RV/EV | 0.33 | | | | | + | + (lags 1 − 7) | | | |
| RV/EV→PIV | | | | | | + | + (lags 1 − 4) | | | |
| PIV→RSV | −0.35 | | | | | | | −0.654 | | |
| RSV→PIV | | | | | | | | | | |
| PIV→SARS-CoV-2 | −0.25 | | | | | | − (lags 1 − 6) | | | |
| SARS-CoV-2→PIV | | | | | + | | | | (*) | |
| PIV→hCoV | | | | | | + | | 0.386 | | |
| hCoV→PIV | | | | | | + | + (lags 1 − 7) | | | |
| RV/EV→RSV | | | | | | | + (lags 6 − 8) | | | 25% |
| RSV→RV/EV | | | | | | | − (lags 1 − 6) | | | |
| RV/EV→SARS-CoV-2 | 0.35 | | | + | | + | + (lags 1 − 6) | 0.736 | | |
| SARS-CoV-2→RV/EV | | | | | + | + | | | | |
| RV/EV→hCoV | | - | - | - | | + | − (lags 1 − 6) | | | |
| hCoV→RV/EV | | | | | | + | | | | |
| RSV→SARS-CoV-2 | 0.44 | | | | | + | − (lags 7 − 8) | 0.761 | | |
| SARS-CoV-2→RSV | | + | + | | | + | + (lags 1 − 8) | | | 38% |
| RSV→hCoV | | | | | | | | | | 25% |
| hCoV→RSV | | | | | | + | | | | 25% |
| SARS-CoV-2→hCoV | | + | | + | | + | + (lags 6 − 8) | | | |
| hCoV→SARS-CoV-2 | | | | | | | − (lags 7 − 8) | | | |
